# Multiethnic Prediction of Nicotine Biomarkers and Association with Nicotine Dependence

**DOI:** 10.1101/2020.12.23.20248734

**Authors:** Andrew W. Bergen, Christopher S. McMahan, Stephen McGee, Carolyn M. Ervin, Hilary A. Tindle, Loïc Le Marchand, Sharon E. Murphy, Daniel O. Stram, Yesha M. Patel, Sungshim L. Park, James W. Baurley

## Abstract

**Background:** The nicotine metabolite ratio and nicotine equivalents are measures of metabolism rate and intake. Genome-wide prediction of these nicotine biomarkers will extend biomarker studies to cohorts without measured biomarkers and enable tobacco-related behavioral and exposure research.

**Methods:** We screened genetic variants genome-wide using marginal scans and applied statistical learning algorithms on top-ranked genetic variants and age, ethnicity and sex, and cigarettes per day (CPD) (in additional modeling) to build prediction models for the urinary nicotine metabolite ratio (uNMR) and creatinine-standardized total nicotine equivalents (TNE) in 2,239 current cigarette smokers in five ethnic groups. We predicted these nicotine biomarkers using model ensembles, and evaluated external validity using behavioral outcomes in 1,864 treatment-seeking smokers in two ethnic groups.

**Results:** The genomic regions with the most selected and trained variants for measured biomarkers were chr19q13.2 (uNMR, without and with CPD) and chr15q25.1 and chr10q25.3 (TNE, without and with CPD). We observed ensemble correlations between measured and predicted biomarker values for the uNMR and TNE without (with CPD) of 0.67 (0.68), and 0.65 (0.72) in the training sample. We observed inconsistency in penalized regression models of TNE (with CPD) with fewer variants at chr15q25.1 selected and trained. In treatment-seeking smokers, predicted uNMR (without CPD) was significantly associated with CPD, and predicted TNE (without CPD) with CPD, Time-To-First-Cigarette, and Fagerström total score.

**Conclusions:** Nicotine metabolites, genome-wide data and statistical learning approaches develop novel robust predictive models for urinary nicotine biomarkers in multiple ethnic groups. Predicted biomarker associations help define genetically-influenced components of nicotine dependence.

**IMPLICATIONS:** We demonstrate development of robust models and multiethnic prediction of the urinary nicotine metabolite ratio and total nicotine equivalents using statistical and machine learning approaches. Trained variants in models for both biomarkers include top-ranked variants in multiethnic genome-wide studies of smoking behavior, nicotine metabolites and related disease. Association of the two predicted nicotine biomarkers with Fagerstr□m Test for Nicotine Dependence items support models of nicotine biomarkers as predictors of physical dependence and nicotine exposure. Predicted nicotine biomarkers may facilitate tobacco-related disease and treatment research in samples with genomic data and limited nicotine metabolite or tobacco exposure data.

## INTRODUCTION

Cigarette smoking remains the largest modifiable cause of mortality in the United States, responsible for one-third of deaths due to cancer and cardiovascular disease, and most pulmonary disease.^1^ Tobacco control programs and smoking cessation therapies have reduced smoking prevalence 67% over the last 50 years in the United States; yet, in 2018, there were 34 million adult cigarette smokers, with numerous use disparities by demographic, economic and health conditions.^1^

Nicotine (NIC) is the tobacco constituent responsible for sustained tobacco use.^2^ The nicotine metabolite ratio (NMR, the ratio of *trans*-3’-hydroxycotinine, 3HC, to cotinine, COT), is a biomarker of CYP2A6 activity. CYP2A6 is the primary catalyst of nicotine in smokers. The ratio of these two nicotine metabolites may be measured via laboratory analysis of blood, saliva or urine.^3,4^ Total nicotine equivalents (TNE) is a biomarker of nicotine consumption and is defined as the molar sum of the urinary concentration of total NIC, total COT, total 3HC, and additional metabolites depending on the particular study (“total” refers to the sum of the compound and its glucuronides).^5^ In addition to serving as a biomarker of nicotine metabolism and consumption,^6^ the NMR is associated with the efficacy of multiple tobacco cessation therapies with potential use for personalizing treatment for tobacco use disorder,^7^ while TNE is associated with smoking behaviors and toxicant exposures that may account for some race/ethnicity lung cancer risk disparities.^8,9^

Predictive genetic modeling of nicotine biomarkers promises to provide genetic signatures supporting disease, mechanistic, and treatment research. Genetic modeling of the NMR is supported by significant twin and locus specific heritability estimates.^10–12^ There are no heritability estimates of TNE. Heritability estimates of cigarettes per day (CPD), a less precise measure of consumption, are significant in twin and genome-wide approaches,^13–15^ but lower than NMR estimates.

Predictive genetic modeling of nicotine metabolism and initial applications have encompassed laboratory studies, research cohorts and cessation trials; modeling focused first on candidate gene variants and then leveraged variants from genome-wide analyses. Predictive genetic models of CYP2A6-mediated nicotine metabolism have been developed that account for approximately 38% to 62% of NMR variance^16–18^. Herein, we build and characterize prediction models of two urinary nicotine biomarkers from metabolite, demographic and genome-wide data in current smokers of multiple ethnicities.^19^ Using statistical learning techniques, we trained models which robustly predicted two urinary biomarkers in five ethnic groups. We applied these models to predict both biomarkers in treatment-seeking smokers in two ethnic groups,^20^ and explored predicted biomarker associations with nicotine dependence measures.^21^ We relate findings to prior analyses and review prospects for translation.

## MATERIALS AND METHODS

### Ethical Approval

Written informed consent was obtained from all participants. The research described herein received approvals from the Institutional Review Boards of BioRealm, the Oregon Research Institute, the University of Hawaii and the NIH Joint Addiction, Aging, and Mental Health Data Access Committee.

### Participants, Measured Biomarkers and Nicotine Dependence Measures

We utilized participant data from two multiethnic studies in this secondary data analysis: current smokers from the Multiethnic Cohort study (MEC), initially assembled in 1993 at the University of Hawaii Cancer Center and Department of Preventive Medicine, University of Southern California, to study diet and cancer; and, treatment-seeking smokers recruited by the University of Wisconsin Transdisciplinary Tobacco Use Research Center (UW-TTURC), at the Center for Tobacco Research and Intervention, established in 1992 to study nicotine dependence and deliver smoking cessation treatments. MEC study participants were not compensated for their participation; UW-TTURC participants were compensated for participation in the smoking cessation trial.

We studied a subsample of MEC current smokers who provided (2004-2006) blood and urine samples and epidemiologic data to enable research on genomics and tobacco exposures.^22^ Urinary total and free NIC, COT, and 3HC, and free nicotine N-oxide (NNO), were measured.^22^ The natural log-transformed urinary NMR (uNMR, defined as total 3HC/free COT), and the square root-transformed TNE (the creatinine-standardized molar sum of total NIC, COT, and 3HC, and free NNO) were the two nicotine biomarkers analyzed in this study.

We studied UW-TTURC smokers recruited and randomized (2000-2010) into three smoking cessation trials,^23–25^ who provided a blood sample to enable research on genetics and nicotine addiction (dbGaP phs000404.v1.p1).^20^ The dataset includes four self-administered nicotine dependence measures: the Fagerstr□m Test of Nicotine Dependence (FTND),^26^ the Tobacco Dependence Screener (TDS),^27^ the Nicotine Dependence Syndrome Scale (NDSS),^28^ and the Wisconsin Inventory of Smoking Dependence Motives (WISDM).^29^

See **Supplementary Material** for details on MEC metabolite and genomic data and UW-TTURC demographic, dependence, and genomic data.

### Variable Selection Phase

The variable selection phase makes use of a marginal scan to examine each genetic variant through a model of the form

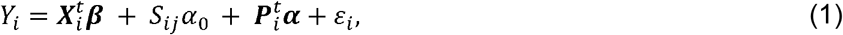

where *Y*_*i*_ is the biomarker level for the ith individual; ***X***_*i*_ is a vector of confounding variables with corresponding ***β*** regression coefficients; *S*_*i j*_ is the genetic variant in question with a_O_ as the corresponding regression coefficient; ***P***_*i*_ is a vector of principal components computed on the genotypes design matrix and ***α*** is the corresponding vector of regression coefficients; and *ε* _*i*_ is the usual error term. In our primary genome-wide analyses, we included age, sex, ethnicity and BMI, and, in additional modeling, CPD was added (“with CPD”). We include the first 50 principal components (PCs) of the genotype design matrix, which more than adequately accounts for genetic relatedness and ancestry among the study participants; i.e., the first 50 PCs explain 72% of PC variance. The model depicted in (1) was fit for each genetic variant in the MEC data with Smokescreen database annotation, and p-values for the test of *H*_0_:*α* _0_=0 vs. *H*_1_: *α*_0_ ≠ 0 were computed. This phase was completed by selecting 200 genetic variants based on the smallest p-values to move into the training phase.

### Training Phase

The training phase of our prediction process makes use of a suite of high dimensional regression and machine learning techniques. In particular, we fit first order models using the LASSO, elastic net, adaptive LASSO, and the adaptive elastic net regression methodologies,^30–33^ where predictor variables were the selected genetic variants, age, sex and ethnicity, with CPD added in an additional set of models. In this implementation, the penalty parameters were tuned to minimize the Bayesian information criterion (BIC). In each elastic net model we considered five settings (i.e., 0.20, 0.35, 0.5, 0.65, 0.8) for the penalty mixing parameter and in each adaptive method we consider five weighting schemes based on *a priori* fits. This leads to a total of 36 fitted regression models. In addition to these regression models, we also make use of three machine learning algorithms to train models. We fit: a regression tree,^34^ tuned for the minimum number of splits and maximum depth of the tree via five-fold cross validation; bagging,^35^ tuned for the number of trees; and gradient boosting machine,^36,37^ tuned for step size of each boosting step, maximum depth of tree, minimum sum of instance weight (Hessian) needed in a child, subsample ratio of the training instance and subsample ratio of columns when constructing each tree via five-fold cross validation.

### Prediction Phase

The prediction phase leverages the 39 trained models to perform out of sample prediction. In particular, we form the following predictions

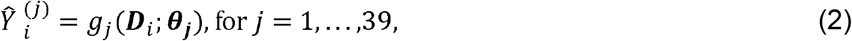

Where 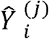 denotes the predicted nicotine biomarker level for the ith subject in the UW-TTURC data, ***D***_*i*_ denotes the demographics and genotypes available on the ith subject, *g*_*j*_(. ;.) denotes the form of the *j*th model, and ***θ*** _***j***_ denotes the set of trained parameters for the *j*th model. These predictions are then used to construct an ensemble based prediction. Briefly, ensemble methods obtain better predictive performance by aggregating over the predictions of multiple statistical/machine learning algorithms. In our application, as is the common approach, we use the following predictive aggregation

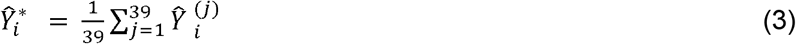

In this analysis, genotypes from selected variants were extracted from African American and white UW-TTURC participants (dbGaP phs000404.v1.p1), and cross-referenced to the Smokescreen database^38^ by chromosome and position. Dosages were transformed as needed to count Smokescreen alternate alleles. Nicotine biomarker prediction and prediction aggregation were as described using the trained models without and with CPD.

#### Variant Annotation

Variant annotation (GRCh37/hg19 assembly) was from the Ensemble Variant Effect Predictor.^39^ Trained variant-related gene associations with smoking-related phenotypes were from the Ensemble GWAS Catalog.^40^

### Measured and Predicted Biomarker Demographic Differences

We estimated significant differences in covariate-adjusted measured biomarkers in African American and white MEC participants and in predicted biomarkers in UW-TTURC participants by sex and by ethnicity.

### Predicted Biomarkers and Nicotine Dependence Measures

Predicted nicotine biomarkers uNMR and TNE were individually included in linear regression of each score of four nicotine dependence measures. Each model was adjusted for age, sex and ethnicity. Regressions were also performed to evaluate interactions with ethnicity and with sex.

## RESULTS

There were 2,239 MEC participants in five ethnic groups with biomarker and genotype data available for variable selection and model training. There were 1,864 UW-TTURC participants in two ethnic groups with genome-wide data available for model prediction, and 1,800 to 1,862 participants with nicotine dependence data.^20^ Participant age and sex distributions reflect study designs. Ethnicity distributions reflect study designs and recruitment locations, and selection of African American and white treatment-seeking smokers for prediction. CPD distributions reflect study design (current smokers) and trial recruitment criteria (treatment-seeking smokers). See **Table 1**.

**Table 1:** Samples Included in Nicotine Biomarker Modeling and Prediction

| Characteristic | MEC | UW-TTURC |
| --- | --- | --- |
| Participant N | 2,239 | 1,864 |
| Age<sup>a</sup> Mean (SD) | <sup>y</sup>63.9 (7.2) | <sup>y</sup>43.4 (11.3) |
| Female N (%) | <sup>y</sup>1199 (53.6%) | <sup>y</sup>1090 (58.5%) |
| Ethnicity N (%) |  |  |
| African American | 364 (16.3%) | 260 (14.0%) |
| Native Hawaiian | 311 (13.9%) | - |
| Japanese American | 674 (30.1%) | - |
| Latinos | 453 (20.2%) | - |
| White | 437 (19.5%) | 1604 (86.0%) |
| Cigarettes per Day | <sup>y</sup> | <sup>y</sup> |
| 1 – 10 | 1,168 (52.2%) | 99 (5.3%) |
| 11 - 20 | 870 (38.9%) | 988 (53.1%) |
| 21 - 30 | 119 (5.3%) | 533 (28.6%) |
| ≥31 | 82 (3.5%) | 242 (13.0%) |

<sup>a</sup>MEC age at biospecimen collection, UW-TTURC age at baseline interview.
<sup>a</sup> $P < .05$ , <sup>$\beta$</sup> $P < .005$ , <sup>$\gamma$</sup> $P < .001$ . Ethnicity proportions not tested.

### Measured Nicotine Biomarkers

The two covariate-adjusted (without or with CPD) biomarkers in African American and white MEC participants were significantly related to each other in a linear model (*p−values <* .001). We observed statistically significant higher levels of covariate-adjusted uNMR without CPD in female versus male participants (*P <* .001), but no significant differences between African American and white participants. There were no significant differences in covariate-adjusted uNMR with CPD by sex or ethnicity. We observed statistically significant higher levels in female participants and lower levels in African American participants of covariate-adjusted TNE without CPD, than in male or white participants, respectively (*p−values <* .001). We observed statistically significant differences in covariate-adjusted TNE with CPD by sex (*p−values <* .001), but not by ethnicity. See **Table 2** (African American and white participants) and **Supplementary Tables 1A** and **1B** (five ethnicities).

**Table 2:** Measured (MEC) and Predicted (UW-TTURC) Nicotine Biomarkers By Sex and Ethnicity, African American and White

| Biomarker | Female |  | Male |  | African American |  | White |  |
| --- | --- | --- | --- | --- | --- | --- | --- | --- |
|  | Mean | (SE) | Mean | (SE) | Mean | (SE) | Mean | (SE) |
| Measured | N = 500 |  | N = 301 |  | N = 364 |  | N = 437 |  |
| uNMR | <sup>γ</sup>1.48 | (0.03) | <sup>γ</sup>1.36 | (0.04) | 1.45 | (0.04) | 1.40 | (0.04) |
| uNMR<sub>CPD</sub> | 1.20 | (0.06) | 1.09 | (0.06) | 1.44 | (0.07) | 1.32 | (0.07) |
| TNE | <sup>γ</sup>8.03 | (0.12) | <sup>γ</sup>7.71 | (0.13) | <sup>γ</sup>7.27 | (0.12) | <sup>γ</sup>8.32 | (0.11) |
| TNE<sub>CPD</sub> | <sup>γ</sup>7.43 | (0.18) | <sup>γ</sup>6.42 | (0.18) | 6.99 | (0.20) | 7.42 | (0.20) |
| Predicted | N = 1090 |  | N = 774 |  | N = 260 |  | N = 1604 |  |
| uNMR | 1.46 | (0.01) | 1.45 | (0.01) | <sup>γ</sup>1.52 | (0.01) | <sup>γ</sup>1.45 | (0.01) |
| TNE | <sup>γ</sup>8.46 | (0.04) | <sup>γ</sup>7.91 | (0.04) | <sup>γ</sup>7.43 | (0.06) | <sup>γ</sup>8.36 | (0.03) |

\*Natural log transformed, no units. \*\*Square root transformed, nmol/mg creatinine. For measured nicotine biomarker values by sex, values are adjusted by age and ethnicity (and CPD, where indicated), and ethnicity strata values are adjusted by age and sex (and CPD, where indicated). For predicted nicotine biomarker values, age, sex and ethnicity (and CPD, where indicated) were included in the models. <sup>α</sup>*P* < .05, <sup>β</sup>*P* < .005, <sup>γ</sup>*P* < .001.

### Nicotine Biomarker Genome-wide Analysis

The number of variants in all genome-wide analyses in the MEC was N=542,732. See **Table 3** and **Supplementary Tables 2A, 2B, 3A** and **3B** for selected variant summaries and details. The genome-wide analysis of measured covariate-adjusted uNMR without CPD identified N=122 genome-wide significant (*P <* 5E*−*8) associations at chr19q13.2, and associations (*p−values <* 6.3E-7) on N=11 additional autosomes. The region of genome-wide significant association on chr19q13.2 included 158 variants, spanned 173 kilo basepairs (kbp), and included variants at the protein-coding genes *SNRPA, RAB4B, MIA, EGLN2, CYP2A6* and *CYP2A7*. The most significant marginal result genome-wide was rs56113850 (C allele, β = 0.40, *P* = 5.4E-48), in the fourth intron of *CYP2A6*. An additional 17 selected variants spanned 332 kbp at chr19q13.2, including variants at protein-coding genes *NUMBL, ADCK4, ITPKC* and *CYP2B6*.

**Table 3:** Variants Selected and Trained in Penalized Regression Models, MEC

| Chr* | uNMR |  | uNMR<sub>CPD</sub> |  | TNE |  | TNE<sub>CPD</sub> |  |
| --- | --- | --- | --- | --- | --- | --- | --- | --- |
|  | Select | Train** | Select | Train** | Select | Train** | Select | Train** |
| 1 | 4 | 3/3 | 4 | 3/4 | 25 | 12/22 | 24 | 15/22 |
| 2 | 4 | 3/4 | 5 | 3/5 | 4 | 4/4 | 4 | 4/4 |
| 3 | 5 | 4/4 | 3 | 3/3 | 5 | 5/5 | 3 | 3/3 |
| 4 | 0 | -/- | 0 | -/- | 10 | 9/10 | 13 | 12/13 |
| 5 | 3 | 3/3 | 4 | 4/4 | 8 | 6/7 | 17 | 11/14 |
| 6 | 0 | -/- | 0 | -/- | 6 | 6/9 | 9 | 9/9 |
| 7 | 1 | 1/1 | 1 | 1/1 | 5 | 5/5 | 9 | 9/9 |
| 8 | 3 | 1/1 | 3 | 1/1 | 24 | 14/23 | 25 | 17/23 |
| 9 | 0 | -/- | 0 | -/- | 4 | 4/4 | 5 | 4/5 |
| 10 | 1 | 1/1 | 1 | 1/1 | 16 | 8/13 | 16 | 10/13 |
| 11 | 1 | 1/1 | 0 | -/- | 14 | 11/14 | 20 | 10/19 |
| 12 | 0 | -/- | 0 | -/- | 3 | 3/3 | 3 | 3/3 |
| 13 | 1 | 1/1 | 1 | 1/1 | 2 | 2/2 | 6 | 6/6 |
| 14 | 0 | -/- | 1 | 1/1 | 1 | 1/1 | 1 | 1/1 |
| 15 | 1 | 1/1 | 1 | 1/1 | 34 | 9/29 | 7 | 4/7 |
| 16 | 1 | 1/1 | 1 | 1/1 | 1 | 1/1 | 3 | 3/3 |
| 17 | 0 | -/- | 0 | -/- | 3 | 3/3 | 4 | 3/4 |
| 18 | 0 | -/- | 0 | -/- | 4 | 4/4 | 4 | 4/4 |
| 19 | 175 | 43/151 | 174 | 43/148 | 20 | 6/18 | 15 | 7/12 |
| 20 | 0 | -/- | 0 | -/- | 7 | 7/7 | 7 | 6/6 |
| 21 | 0 | -/- | 0 | -/- | 1 | 1/1 | 0 | -/- |
| 22 | 0 | -/- | 0 | -/- | 3 | 3/3 | 5 | 5/5 |

\*Chr = chromosome. \*\*The number of variants trained / the number of variants available to be trained. Note: The number of variants available to be trained in the training sample excluded variants not available in the prediction sample. See **Supplementary Material** for details.

The primary genome-wide analysis of measured covariate-adjusted TNE without CPD identified variant associations (*p−values <* 2.6E*–*7) on all autosomes among the top 200 variants. The region with the most variants selected (31 variants) was chr15q25.1, spanned 203 kbp, and included variants at protein-coding genes *IREB2, HYKK, PSMA4, CHRNA5, CHRNA3* and *CHRNB4*. The most significant marginal result variant in this region was rs2036527 (A allele, β = 0.57, *P* = 1.4E-5), proximal of *CHRNA5*. The region with the top-ranked variant in the genome-wide analyses of TNE (rs56113850, C allele, β = 0.43, *P* = 2.6E-7) was chr19q13.2 (20 variants), spanned 109 kbp, and included variants at protein coding genes *CYP2A6* and *CYP2A7*.

Results of genome-wide analysis of the uNMR with CPD were nearly identical to the analysis without CPD, e.g., 87% of selected variants in both uNMR analyses were found at chr19q13.2. The genome-wide analysis of TNE with CPD exhibited reduced marginal significance (*p−values* < 3.4E-6), reduced numbers of variants in the chr15q25.1 region (four versus 31 variants selected), and different regions with most variants selected (12 chr10q25.3 versus 31 chr15q25.1 variants), than in analysis of TNE without CPD.

### Model Training, Variants, Covariates and Associated Genes

Variants selected in genome-wide analyses but not available in UW-TTURC data were not included in model training. See **Table 3** and **Supplementary Tables 2A, 2B, 3A**, and **3B** for trained variant summaries and model counts.

As expected, most trained variants in the uNMR model without CPD and associated protein-coding genes (43/63 variants and 10/19 genes) were located on chr19q13.2 from *NUMBL* to *CYP2B6*. Clinical covariates trained in 38 uNMR models included age (22 models), sex (27 models), and ethnicity (38 models). Several chr19q13.2 SNPs were trained in the two machine learning models reviewed (**Supplementary Figures**) with rs56113850 included in all models reviewed. In one machine learning method (**Supplementary Figure 1**), Japanese American ethnicity dichotomized uNMR, with chr19q13.2 variants defining the remaining tree structure.

Training variants in TNE models without CPD resulted in 124 trained variants located on all autosomes. The most trained variants were found on chromosomes 1, 8, 11, and 15. The regions with the largest number of trained variants were chr15q25.1 (eight variants), from *HYKK* to *CHRNB4*, and chr19q13.2 (six variants), from rs12459249 proximal of *CYP2A6* to the *CYP2A7-CYP2B6* intergenic region. Eighty-five of 124 trained variants were trained in all penalized regression models on all autosomes except 16 and 21. Clinical covariates trained in 38 TNE models reviewed included age (one model), sex (37 models), and ethnicity (38 models). In one machine learning model (**Supplementary Figure 2**), a chr15q25.1 variant dichotomized TNE, sex dichotomized lower values, and Latino ethnicity and a chr22q13.2 variant trichotomized higher values. Trained variants were annotated to 53 protein-coding genes distributed over all autosomes. Thirty-six of 47 annotated protein-coding genes in the GWAS catalog have associations with smoking related behaviors, diseases or traits, and five have associations with kidney function (data not shown).

In uNMR models without and with CPD, 40 of 43 trained chr19q13.2 variants and 18 of 20 trained non-chr19q13.2 variants were identical. In uNMR models with CPD, CPD was trained in 36 of 38 models reviewed, and age and sex were trained in 25 and 36 models. Across trained variants in common between uNMR models with and without CPD, there were only minor differences in the number of models trained. However, in TNE models with CPD, the number of trained variants increased and the mean, median, mode, and maximum number of models variants were trained in decreased. The number of trained variants declined at chr15q25.1 (from eight to two) and chr19q13.2 (from six to four) and increased elsewhere in the genome. In TNE models with CPD, CPD was trained in all 38 models reviewed, age was trained in six additional models and ethnicity was trained in eight fewer models. In one TNE machine learning model CPD replaced the chr15q25.1 variant, and sex replaced Latino ethnicity and the chr22q13.2 variant, as prediction criteria (data not shown).

### Training of Nicotine Biomarker Models

For each of 39 model fits, we evaluated the final form of the model via standard model diagnostic techniques, e.g., residual plots. From these diagnostics, we discovered no evidence that the assumed forms of the models were invalid. To assess the model fits, the correlation between measured and fitted nicotine biomarkers were computed for each model and biomarker in the MEC. The ensemble value of these correlations (*r*) for uNMR and TNE without CPD were 0.6695 and 0.6450, with similar correlations across all penalized regression models for each biomarker (data not shown). These values tend to indicate good fit and do not point to overfitting issues. This supports equal weighting for each contributing model in constructing our ensemble-based estimators. For measured vs fitted biomarker correlations with CPD, the ensemble values were 0.6760 and 0.7162 for uNMR and TNE. Penalized regression model correlations within each uNMR analysis (without and with CPD) were similar to each other, but penalized regression model correlations in TNE analysis with CPD dropped from 0.73 to 0.42 as penalty parameters increased (data not shown), reflecting the loss of highly correlated or confounded variables.

### Predicted Biomarkers in the UW-TTURC

Given minimal differences in model and ensemble correlations between the two analyses for the uNMR, and evidence for confounding in penalized regression TNE models with CPD, we focus further reporting on predicted biomarkers modeled without CPD. Using the ensemble-based models without CPD generated in the MEC, predictions were obtained for both nicotine biomarkers for all UW-TTURC participants (**Table 2**). Predicted uNMR and predicted TNE in participants were significantly related to each other (β(SE) = .017(.005), *P <* .001). Predicted uNMR was significantly higher in African American than white participants (*P <* .001), but there was no significant difference in predicted uNMR by sex (*P* = 0.28). Predicted TNE was significantly larger in female than male participants, and significantly smaller in African American than white participants (*p−values <* .001).

### Predicted uNMR and Nicotine Dependence

Predicted uNMR was positively associated with FTND CPD (*P* = .002), WISDM Automaticity (*P* = .049), and NDSS Tolerance (*P* = .022) (**Table 4**). In additional analyses, interactions of ethnicity and of sex with predicted uNMR (ethnicity *P* = .041, sex *P* = .024) were observed with NDSS Continuity, and of sex with predicted uNMR (*P* = .045) were observed with NDSS Stereotypy (**Supplementary Table 10**).

**Table 4:** Predicted Biomarkers (1° Model) and Nicotine Dependence, Behavior Model

| Dependence | N | uNMR |  | TNE |  |
| --- | --- | --- | --- | --- | --- |
| Measure |  | Coefficient | SE | Coefficient | SE |
| FTND |  |  |  |  |  |
| Total | 1843 | 0.129 | 0.195 | <sup>a</sup>0.099 | 0.045 |
| CPD | 1862 | <sup>β</sup>0.211 | 0.068 | <sup>a</sup>0.039 | 0.016 |
| TTFC | 1861 | -0.008 | 0.078 | <sup>a</sup>0.041 | 0.018 |
| WISDM |  |  |  |  |  |
| Automaticity | 1800 | <sup>a</sup>0.297 | 0.151 | -0.011 | 0.035 |
| Loss of Control | 1800 | -0.021 | 0.127 | 0.033 | 0.029 |
| Craving | 1800 | -0.156 | 0.120 | 0.025 | 0.028 |
| Tolerance | 1800 | 0.147 | 0.127 | <sup>a</sup>0.060 | 0.029 |
| Total PDM | 1800 | -0.003 | 1.183 | -0.065 | 0.273 |
| NDSS |  |  |  |  |  |
| Drive | 1809 | -0.062 | 0.096 | 0.005 | 0.022 |
| Priority | 1820 | 0.019 | 0.097 | -0.033 | 0.022 |
| Tolerance | 1814 | <sup>a</sup>0.239 | 0.104 | 0.032 | 0.024 |
| Continuity | 1815 | 0.033 | 0.094 | 0.015 | 0.022 |
| Stereotypy | 1813 | -0.012 | 0.096 | <sup>β</sup>0.066 | 0.022 |
| NDSS-T | 1800 | 0.008 | 0.086 | 0.022 | 0.020 |

<sup>a</sup> $P < .05$ , <sup>β</sup> $P < .005$

### Predicted TNE and Nicotine Dependence

Predicted TNE was positively associated with: FTND total score (*P* = .027), CPD (*P* = .014) and time-to-first-cigarette (TTFC) (*P* = .022); with WISDM Tolerance (*P* = .042); and NDSS Stereotypy (*P* = .003) (**Table 4**). In additional analyses, interaction of ethnicity with predicted TNE (*P* = .0036) was observed with NDSS Stereotypy (**Supplementary Table 10**).

## DISCUSSION

### Genome-wide analysis, selection and training of variants

Our analyses describe the first genome-wide modeling and prediction of the uNMR using statistical and machine learning approaches, and the first genome-wide modeling and prediction of TNE, as far as we are aware. These analyses demonstrate internal or analytic validity in current smokers and external validity in treatment-seeking smokers with prior genome-wide, biomarker and nicotine dependence findings. We modeled the two nicotine biomarkers throughout the analysis workflow without and with self-reported CPD coded as in the FTND, as CPD is a significant predictor of TNE and the uNMR. As expected, inclusion of CPD had limited influence on modeling of the uNMR, and resulted in fewer chr15q25.1 SNPs selected and trained in modeling of the TNE. We concentrate our discussion on the results of modeling the two biomarkers without CPD.

As expected from prior genome-wide studies, most (87%) variants selected in the uNMR genome-wide analysis were from the chr19q13.2 *CYP2A6* region. We previously identified rs56113850 in the MEC as the top-ranked variant for uNMR in all ethnic groups tested,^19^ and as a *cis* expression Quantitative Trait Locus (*cis* eQTL) in liver and lung for *CYP2A6*.^41^ rs56113850 has been identified as top-ranked in genome-wide studies of the NMR in smokers of European descent.^11,12^ Nearly a third (31%) of trained variants in uNMR models were located in non-chr19q13.2 genomic regions. Four of six non-chr19q13.2 protein-coding genes with variants trained in uNMR models (*DAB1, CPNE4, CAMKMT* and *RMBS3*) have prior associations with smoking-related behaviors and disease in the GWAS catalog (data not shown). This adds to the non-chr19q13.2 genes with variants trained in models of nicotine metabolism.^17^

Variant selection and training in TNE modeling was more polygenic than for the uNMR, consistent with our understanding of nicotine pharmacology,^6^ and dominant nicotine-related loci characterized in genome-wide studies.^42^ We previously identified the top-ranked trained variant for TNE (rs2036527 in chr15q25.1) as top-ranked in genome-wide studies of CPD and of lung cancer in African Americans.^43,44^ This variant is the top-ranked variant in genome-wide studies of blood-based COT and of COT+3HC levels in European ancestry smokers and a *cis* eQTL for *CHRNA5* and other chr15q25.1 genes.^12^ In our analysis, rs2036527 was trained in seven penalized regression models of TNE; among chr15q25.1 trained variants, only rs55676755 was trained in all penalized regression models. Association of rs55676755 with pulmonary disease and functional measures in multiethnic genome-wide studies^45^ supports the hypothesis that rs55676755 is a predictor of smoking-related toxicant exposures in multiple ethnicities.

We and others previously identified the proximal TNE model trained variant in the chr19q13.2 region (rs12459249) as the top-ranked variant in genome-wide analyses of the laboratory-based NMR in three ethnicities,^41^ and the blood-based NMR in African American smokers.^46^ Among six trained chr19q13.2 region variants in TNE modeling, rs12459249 was trained in six penalized regression models, while rs56113850 and rs73038469 in this region were trained in all penalized regression models. Both rs56113850 and rs73038469 are *cis* eQTLs for protein-coding and non-coding genes in multiple tissues and *cis* QTLs for methylated cytosine-guanine dinucleotides, supporting possible functional roles in gene regulation.^12^ While multiple chr19q13.2 trained variants were included in models for each biomarker, only rs56113850 was a trained variant in models of both biomarkers.

### Biomarkers, demographics and dependence

These are the first analyses to relate predicted uNMR and predicted TNE to each other, to ethnicity and sex, to major FTND items, and to WISDM and NDSS subscales. Predicted uNMR and TNE in treatment-seeking smokers were significantly associated with each other as are measured uNMR and TNE in current smokers.^19^ Significant differences for both measured and predicted TNE by ethnicity and by sex were observed, in the expected directions for creatinine-standardized TNE.^47^

Prior findings provide support for the associations with nicotine dependence measures we observed using predicted nicotine biomarkers. A systematic review found the measured NMR significantly correlated with CPD in nine of 15 studies (in three of four using the measured uNMR) examined.^48^ Predictive genetic models of the NMR have shown significant associations with CPD in ordinal and continuous coding.^16,18^ An additive wGRS of four independent chr19q13.2 variants of the blood-based NMR was significantly associated with continuous CPD in current smokers.^11^ Measured TNE (24 hour urine, molar sum of NIC, COT, 3HC, and glucuronides, unadjusted for creatinine) was significantly associated with CPD, TTFC and total FTND score in current smokers.^49^

The associations of predicted nicotine biomarkers with components of the WISMD and NDSS measures we observed are novel. Prior associations of smoking constructs provide support for the observed associations. E.g., WISDM Automaticity and Tolerance and NDSS Stereotypy and Tolerance correlations with the FTND and CPD were as large or the largest correlations of 13 WISDM and five NDSS subscales tested in treatment-seeking smokers from two UW-TTURC cessation trials.^21^ NDSS Stereotypy and Tolerance were significantly correlated with multiple physical dependence variables in daily smokers recruited for laboratory studies of smoking cessation medications.^28^

### Strengths and Limitations

Use of a multiethnic cohort for modeling nicotine biomarkers will support translation to studies of smokers of multiple ethnicities in behavioral, disease and treatment research. Further research is needed to assess the performance of multiethnic models in specific ethnic populations.

Our uNMR GWAS and model training identified multiple signals at and outside the chr19q13.2 region. Selection and training of models predicting the uNMR in larger samples may clarify the role of non-chr19q13.2 genes in nicotine metabolism. Additional model development of the NMR and the uNMR may provide clues to the reduced correlation between measured blood NMR and uNMR.^50^

Our TNE genome-wide selection identified top-ranked variants at chr15q25.1 and chr19q13.2 identified in recent genome-wide studies of smoking behavior and nicotine metabolites, and variants in both regions were included in trained models. Research in additional cohorts with measured metabolite data may elucidate how metabolite source, measurement and standardization influence model development and power.

## Conclusions

Concordances observed between our nicotine biomarker modeling and recent genome-wide studies support our goal of developing robust genome-wide prediction models for nicotine biomarkers. Meta-analysis of larger and more diverse samples with respect to participants, biomarkers and clinical data will improve the predictive power of models and enable out-of-sample model validation, but may present challenges with respect to phenotype harmonization. The associations we observed between predicted urinary biomarkers and measures of physical dependence are generally supported by prior analyses of biomarkers and previous models of predicted NMR with similar measures. Availability of smoking cessation trial data will provide an opportunity to characterize relations between genetically determined components of dependence and cessation outcomes and assess translational relevance.

## Data Availability

The Multiethnic Cohort data is available upon application to the Multiethnic Cohort (https://www.uhcancercenter.org/for-researchers/mec-data-sharing). The University of Wisconsin data is available upon application to the Database of Genotypes and Phenotypes (https://www.ncbi.nlm.nih.gov/projects/gap/cgi-bin/study.cgi?study_id=phs000404.v1.p1).

## FUNDING

This work was supported by the National Institute on Alcohol Abuse and Alcoholism (R44 AA027675 to AWB, CSM, SM, CME, SLP, and JWB) and by the National Cancer Institute (R01 CA232516 to HAT, and U01 CA164973 and P01 CA138338 to LLM, DOS, SEM, YMP and SLP). The sponsors had no role in the analysis of data, writing of the report, or in the decision to submit the paper for publication.

## DECLARATION OF INTERESTS

AWB is an employee of Oregon Research Institute and Oregon Community and Evaluation Services, and serves as a Scientific Advisor and Consultant to BioRealm, LLC. CME is a co-owner and the Principal Biostatistician for BioRealm, LLC. HAT has served as PI on NIH-supported studies for smoking cessation in which the medication was donated by the manufacturer (e.g., Pfizer, varenicline). CSM, SM, LLM, DOM, SEM, YMP and SLP have no conflicts of interest to report. JWB is an employee and an owner of BioRealm, LLC. BioRealm, LLC offers services related to the Smokescreen Genotyping Array and analysis of nicotine biomarkers.

## ACKNOWLEDGMENTS

The authors thank participants of the Multiethnic Cohort Study and of the University of Wisconsin cessation trials. The authors acknowledge the contribution of data from Genetic Architecture of Smoking and Smoking Cessation accessed through dbGAP (phs000404.v1.p1). Support for genotyping, which was performed at the Center for Inherited Disease Research (CIDR), was provided by 1 X01 HG005274-01. CIDR is fully funded through a federal contract from the National Institutes of Health to The Johns Hopkins University, contract number HHSN268200782096C. Assistance with genotype cleaning, as well as with general study coordination, was provided by the Gene Environment Association Studies (GENEVA) Coordinating Center (U01 HG004446). Funding support for collection of the University of Wisconsin Transdisciplinary Tobacco Use Research Center cessation trials was provided by P50 DA019706 and P50 CA084724.

