## Supplementary for "Multiethnic Prediction of Nicotine Biomarkers and Association with Nicotine Dependence"

**for**

**CONTENTS**

**1 Note on Ethnicity Labels 4**

**2 Measured Nicotine Biomarkers, MEC Current Smokers 4**

**3 Genomic Data Analysis 5**

3.1 Genomic Data Quality Control and Imputation 5

3.2 Genome-wide Marginal Result, Annotation and Trained Models 6

**4 Demographics, UW-TTURC Trials and Genetic Substudy 7**

4.1 Genetic Substudy (GSS) and RCTs, Differences by RCT 7

4.2 Demographics of the GSS Sample 8

**5 Nicotine Dependence Measures, GSS and GSS *vs* two RCTs 9**

5.1 The Fagerström Test for Nicotine Dependence, FTND 9

5.2 Tobacco Dependence Screener, TDS 10

5.3 Wisconsin Inventory of Smoking Dependence Motives, WISDM 11

5.4 Nicotine Dependence Syndrome Scale, NDSS 13

**6 Summary - GSS Findings 16**

6.1 GSS Demographic Findings 16

6.2 GSS Nicotine Dependence Findings 16

**7 Predicted Nicotine Biomarkers, Association with Dependence Measures 17**

**8 REFERENCES 18**

**9 TABLES 20**

**10 FIGURE LEGENDS 43**

**11 FIGURES 44**

**List of Tables**

1 Measured Nicotine Biomarkers, Current Smokers, by Ethnicity and Sex 20

2 Top 200 Variants, Urinary Nicotine Metabolite Ratio 22

3 Top 200 Variants, Total Nicotine Equivalents 28

4 Demographics of UW-TTURC Sample, by RCT 35

5 Demographics of the GSS, by Sex and by Ethnicity 36

6 FTND, CPD, and TTFC, by Sex and by Ethnicity 37

7 TDS Dependence, by Sex and by Ethnicity 38

8 WISDM Primary Dependence Motives, by Sex and by Ethnicity 39

9 NDSS Scales in the GSS Participants, by Sex and by Ethnicity 40

10 Predicted Biomarker Interactions, Association with Nicotine Dependence 41

### 1. Note on Ethnicity Labels

The Multiethnic Cohort (MEC) explicitly recruited participants of multiple (five) ethnicities using a combination of civil, health care, and research records. The MEC questionnaire elicited self-identified ethnic background of the participant and parents. The University of Wisconsin Transdisciplinary Tobacco Use Research Center (UW-TTURC) research cohort is a research study of smoking cessation and long-term outcomes, comprised of nicotine dependent research volunteers of any ethnicity recruited in two Wisconsin metropolitan areas using local media. The screening questionnaires of the UW-TTURC smoking cessation trials elicited self-identified race and self-identified Hispanic or Latino ethnicity. In the main text we use the terms "Ethnicity/ethnicity" or "Multiethnic/multiethnic" for self-identified race or ethnicity, and the labels "African American" for Black or African American, and "white" (in text) or "White" (in Tables) for White when referring to the race or ethnicity of MEC or UW-TTURC participants in these two ethnic groups. We use the MEC's ethnicity labels when describing the other three ethnicities in the MEC (e.g., in the main text and in **Supplementary Tables 1A** and **1B**). We use the labels "Black" and "White" when describing UW-TTURC cessation trial and genetic subsample participants of these two ethnicities in the **Supplementary Material**. When we cite work of other authors, we use the race, ethnicity or ancestry labels used by those authors.

### 2. Measured Nicotine Biomarkers, MEC Current Smokers

A subgroup of MEC current smokers without a cancer diagnosis provided overnight or first morning urine for metabolite analysis, blood for genetic analysis, and completed epidemiologic, smoking and concomitant medication questionnaires. To analyze samples for free and total nicotine, cotinine and *trans*-3' hydroxycotinine, diluted urine samples were plated on paired 96 well plates where one plate was incubated overnight with 500 U of *β*-

glucuronidase at 37 C. To analyze samples for free nicotine N-oxide, samples were analyzed as above without a paired plate for *β*-glucuronidase treatment. Deuterium-labeled standards for each nicotine metabolite were added to appropriate plates. Samples were loaded onto conditioned solid phase extraction 96 well elution plates, eluted with ammonium hydroxide:methanol, acidified with formic acid and dried. Re-suspended samples were subjected to liquid chromatography-tandem mass spectrometry (MS/MS) analysis, with separate injections and specific elution times for NIC and NNO, and for COT and 3HC. Specific mass transitions for each nicotine metabolite and each deuterium-labeled-standard were specified for MS/MS fragmentation, monitoring and quantification. See Murphy *et al* for further details on nicotine metabolite analysis methods, levels of quantification, and quality control metrics.[1]

Presented in **Supplementary Tables 1A** and **1B** are the means of measured nicotine biomarkers urinary NMR (natural log transformed total 3HC/free COT) and TNE (square-root transformed creatinine-standardized molar sum of nicotine, cotinine, *trans*-3'hydroxycotinine and nicotine N-oxide) without adjustment and with adjustment for CPD in participants of the five ethnicities recruited in the MEC (African American, Native Hawaiian, White, Latino and Japanese American). The individual level data from these participants were used for variable selection (**Variable Selection Phase**, main text) and for model training (**Training Phase**, main text).

### 3. Genomic Data Analysis

#### *3.1. Genomic Data Quality Control and Imputation*

*MEC Genomic Data.* For MEC, blood leukocyte DNA was genotyped using the Illumina 1M array, and quality control (QC) and imputation were performed previously; for details review Patel *et al*.[2] Samples (and variants) with call rates < 98% (and ≤ 98%), known duplicate samples, close relatives and samples with conflicting or indeterminate sex were removed. Haplotype phasing and genotype imputation of the analysis dataset of 1,131,426 variants and 2,239 samples was performed using the March 2012 1000 Genomes Project release with $r^2 \geq 0.30$ and minor allele frequency (MAF) > 1% by ethnic group.[3–5]

*UW-TTURC Genomic Data.* For UW-TTURC, blood leukocyte DNA was genotyped using the Illumina 2.5M array, and QC and imputation were performed previously; for details, see Laurie *et al* and Bierut *et al*.[6,7] Genotypes were called using BeadStudio, where variants with missing call rate < 2%, Hardy-Weinberg test *p-values* > .0001, and minor allele frequency (MAF) < 0.01 excluded. Self-identified European American and African American samples were imputed separately, using the 1000 Genomes EUR and EUR+AFR reference populations, respectively. Rare markers (MAF < 0.005) were excluded from the reference. Imputation was performed using BEAGLE.[8]

As needed for each variant analysis, annotation and genotype dosage of variants were loaded into a relational database and queried for markers in the Smokescreen Genotyping Array database by chromosome, position, and alleles.[9]

*3.2. Genome-wide Marginal Result, Annotation and Trained Models*

See **Supplementary Tables 2A**, **2B**, **3A** and **3B** for annotations of the top 200 variants selected in the uNMR and the TNE genome-wide studies, including variant rsID, location, marginal scan effect size and significance, and the associated gene annotation. The final annotation in these **Supplementary Tables** (Count) is the number of times the variant was trained in penalized regression models for the corresponding nicotine biomarker. See **Supplementary Tables 2** and **3 ReadMe** for details on annotation definitions.

### 4. Demographics, UW-TTURC Trials and Genetic Substudy

The UW-TTURC dataset we studied is comprised of cigarette smokers who: were recruited into three randomized clinical smoking cessation trials (RCTs),[10–12] agreed to be in a Genetic Substudy (GSS), provided a blood sample for DNA extraction and genotyping,[7] who self-identified as Black or White race, and with complete imputation data. Thus the GSS participants are interested in treatment and in contributing to research. Each RCT was designed to compare different sets of treatment interventions, but had similar inclusion criteria (aged ≥ 18 years, smoking ≥ 10 cigarettes per day, motivated to quit smoking, able to read and write English, willing to complete study assessments) and exclusion criteria (carbon monoxide < 10 ppm, medical and psychiatric comorbidities, risk of pregnancy).

#### *4.1. Genetic Substudy (GSS) and RCTs, Differences by RCT*

We summarize observations and results of tests of demographic differences between the GSS subsamples drawn from the three RCTs, and between the GSS subsamples and the RCTs from which they were recruited (**Supplementary Table 4**).

*RCT Site and Participation in the GSS.* The “ED SR” RCT was performed in Madison,[10] the “Depend” RCT in Milwaukee,[11] and the “TTURC2” RCT in both cities.[12] The participants of the GSS are drawn unequally from the RCTs, i.e., TTURC2 contributes the majority of participants. The number and proportion of participants who participated in the GSS by RCT significantly increases between the RCTs (all $P < .001$).

*Age, Gender, and Ethnicity in the GSS and RCTs.* The age of GSS participants differs significantly by RCT source, i.e., the age of the RCT participants significantly increased from ED SR to Depend to TTURC2 (all $P < .001$), however age does not differ between the GSS sample and the source RCT. Sex proportions did not differ between RCT contributions to the GSS (all $P > .056$), or between each RCT sample within the GSS and each RCT (all $P > .12$). Ethnicity (Black and White) proportions differ between the RCT contributions to the GSS (all $P < .001$) and between RCTs (all $P < .005$), but ethnicity proportions do not differ between RCTs and their contributions to the GSS (all $P > .24$).

*Recruitment into the GSS.* We conclude that recruitment protocols into the GSS became significantly more successful from ED SR to Depend to TTURC2, and that age, sex and ethnicity did not influence recruitment into the GSS. We observed differences in age and ethnicity between the RCTs, but sex proportions did not differ. Ethnicity differences may be related to the city of recruitment.

*Comparison with ED SR and Depend RCTs.* Previously, Piper *et al* observed differences in age, sex, and ethnicity between participants recruited in the ED SR and Depend RCTs.[13] We do not observe differences in sex between the GSS participants recruited from these two RCTs because the ED SR female contribution to the GSS was higher than the female proportion in the ED SR RCT. Differences in age and ethnicity in the ED SR and Depend contributions to the GSS are similar to those previously observed in the complete RCTs.[13]

*4.2. Demographics of the GSS Sample*

The mean (SD) age of the GSS (N=1,864) is 43.41 (11.33) years, N=1,090 (58.5%) are female, and N=260 (14.0%) are Black (**Supplementary Table 5**). The mean duration of smoking is nearly three decades, reflecting the recruitment of treatment-seeking smokers. Male participants are older than female participants ($P = .036$) and Black participants are older than White participants ($P = .0012$). There are proportionally more (fewer) female (male) Black participants than female (male) White participants ($P = .0046$). Age of smoking initiation and endorsement of Hispanic ethnicity do not differ by sex or ethnicity

(Black and White). Female participants have smoked about one year less than male participants ($P < .022$), however, women were younger than men by a similar amount at baseline interview. Years smoked does not differ by ethnicity.

### 5. Nicotine Dependence Measures, GSS and GSS *vs* two RCTs

There are four nicotine dependence measures available for analysis: the Fagerström Test of Nicotine Dependence (FTND),[14] the Tobacco Dependence Screener (TDS),[15] the Nicotine Dependence Syndrome Scale (NDSS),[16] and the Wisconsin Inventory of Smoking Dependence Motives (WISDM).[17] Below the development of each measure is briefly reviewed with notes on heritability. Then values of each measure in the GSS sample are compared by sex and ethnicity, and then to the N=1,071 sample from the complete RCTs (ED SR and Depend) analyzed by Piper *et al*.[13]

#### *5.1. The Fagerström Test for Nicotine Dependence, FTND*

*Development of the FTND.* The FTND was modified from the FTQ, designed to provide a self-report measure of nicotine dependence.[14] The FTND consists of six questions including two (Cigarettes per day, CPD) and the time to first cigarette in the morning, TTFC) with four possible responses, and four dichotomous questions. In studies of European ancestry smokers, FTND CDP and FTND TTFC have significant heritability estimates in twin analyses,[18] while single nucleotide polymorphism (SNP) heritability estimates are significant for CPD codings,[19,20] FTND TTFC,[21,22] and FTND total score.[21,23] Analyses of the phenotypic factor structure of FTND in young adult female twins finds two factors with cross-loadings of CPD and TTFC on both factors;[18] analyses of adult substance use disorder comorbid research subjects finds two phenotype factors with distinct loadings of CPD and TTFC on each factor.[24]

*FTND in the GSS.* In the GSS dataset (**Supplementary Table 6**), FTND total scores are lower in female participants than male participants (*P* < .001), but do not differ by ethnicity. The CPD distribution is shifted lower in female compared to male participants, and in Black compared to White participants (both *P* < .001). While there are no differences in TTFC distributions by sex, the TTFC distribution is shifted higher in Black participants compared to White participants (*P* < .001).

*FTND in the GSS and ED SR and Depend RCTs.* FTND total scores overall and in each sex and ethnicity stratum in the GSS are similar to those in the N=1,071 sample and in each sex and ethnicity stratum analyzed by Piper *et al* (all *P* > .495, data not shown).[13]

*5.2. The Tobacco Dependence Screener, TDS*

*Development of the TDS.* The TDS was developed as a self-administered screener for nicotine dependence.[15] It consists of 10 dichotomous items from the tobacco use section of the Composite International Diagnostic Interview version 1.1,[25] which assesses *ICD-10* and *DSM-III-R* tobacco and nicotine dependence symptoms. The developers validated the screener in three samples of Japanese smokers (University related volunteers, out-patients, and employees of a company) by assessment of internal (Cronbach's *α*) and external (CIDI diagnoses of *ICD-10*, *DSM-III-R* and *DSM-IV* nicotine dependence, and FTQ, CPD, and CO measures) correlations. Analysis of *DSM-IV* nicotine dependence criteria in young adult European ancestry female twins indicate that all criteria except "Gave up activities to smoke" are heritable.[18] SNP heritability for a DSM-IV Nicotine Dependence factor extracted from a European ancestry sample of smokers was significant.[22]

*TDS in the GSS.* In the GSS dataset, female participants endorsed a higher level of TDS dependence then male participants (*P* < .001), and Black participants endorsed a lower level of TDS dependence than White participants (*P* = .042) (**Supplementary Table 7**). In the

N=1,071 sample of ED SR and Depend RCT participants analyzed by Piper *et al* (which share 562 participants (52.5% of 1,071) with the sample analyzed here), female participants endorsed higher TDS nicotine dependence than male participants, and Black participants endorsed lower TDS nicotine dependence than White participants (both $P < .05$, data not shown).[13]

*TDS in the GSS and in the ED SR and Depend RCTs.* TDS nicotine dependence scores in the GSS sample are increased overall and in each sex and ethnicity stratum over the corresponding strata in the ED SR and Depend RCT participants in Piper *et al* (all $P < .05$, data not shown),[13] suggesting that those participating in the GSS from ED SR and Depend, and/or those participating in the GSS from TTURC2, have higher levels of TDS-defined nicotine dependence than the full sample of ED SR and Depend RCT participants.

*5.3. Wisconsin Inventory of Smoking Dependence Motives, WISDM*

*Initial Development of the WISDM.* The WISDM was developed as a nicotine dependence measure with the explicit assumption that dependence is based upon theoretical mechanisms that underlie multiple aspects of smoking behavior.[17] Development began with the psychological community developing questions to measure 13 separate motives for drug use based on a literature review. A derivation sample (N=775) of adult daily and non-daily smokers (60% female, 84% White) recruited from cessation trial participants, university students and local residents answered 285 questions in a large group format and provided a breath sample for CO measurement. Using a portion of the adult smoker sample, items were selected by high internal consistency to generate a subset of questions for the 13 motives. Validation in the remaining smokers, and then by sex, daily versus non-daily smoking and ethnicity demonstrated excellent consistency overall. Confirmatory factor analysis identified stronger support for a multiple (13) factor model than for a single factor model. Multiple regression analyses with CPD, carbon monoxide level and *DSM-IV* dependence indicate the 13 WISDM subscales account for the majority (53% to 60%) of the variance in these concurrent validity items.

*Refinement of the WISDM.* With participants from the derivation study and three cessation trials,[10,11,26] further analyses discovered and characterized four WISDM motives (Automaticity, Craving, Loss of Control and Tolerance) that constitute a common dependence factor,[13] later called Primary Dependence Motives (PDMs).[27] The first analysis used latent profile analyses (LPA) to identify a group of smoking motives that collectively are highly predictive of dependence, including withdrawal and relapse. Initial LPA analysis results indicated that five class solutions were preferred, with four classes consisting of grades of nicotine dependence severity and one class more heavily weighted on the four PDMs, and with lower weighting on the remaining nine classes. The PDM weighted class was named Automatic-Atypical, and had a prevalence of 17% in the smokers from the four samples analyzed. Exploratory Factor Analysis (EFA) together with correlation and eigenplot analyses were performed to provide another analytic perspective. A two factor solution was favored with the four PDMs loading heavily on factor one, eight of the remaining nine loading on factor two, and Cue Exposure/Associative Processes loading moderately on both factors. Intercorrelations of the four heavily loaded subscales in factor one were greater than with the other nine subscales. A third analysis approach, Factor Mixture Analysis (FMA), using two factors was used to evaluate heterogeneity among the multiple classes identified in LPA analysis. These analyses again saw the four PDMs loading heavily on the first factor, and three factors on which the remaining nine subscales were loaded heavily, where the three factors were called Low, Medium and High. Fit criteria were better in the FMA than in the LPA analyses. In regression analyses, factors one and two were found to be significantly related to other measures of nicotine dependence such as the FTND, CPD and carbon monoxide where factor one associations were larger than factor two associations. Finally, factor one was associated with lower education, with alcohol problems, and relapse at 1 week and six months, while factor two was associated with gender (Females > Males).

Further characterization of the WISDM was performed in the analysis of three clinical trials of smoking cessation,[10,11,26] where the four Primary Dependence Motives (Automaticity, Loss of Control, Craving and Tolerance, which are the four subscales of the WISDM available in the dbGaP accession) were found to be more highly correlated to the FTND, CPD, carbon monoxide, withdrawal and relapse than other WISDM subscales.[13]

*WISDM Primary Dependence Motives in the GSS.* In the GSS dataset, female participants endorse higher WISDM Automaticity, Loss of Control and PDM total scores than do male participants (all $P < .05$) (**Supplementary Table 8**). Black participants endorse lower WISDM Loss of Control and higher Tolerance scores than do White participants (both $P < .05$).

*WISDM PDM Scores in the GSS and ED SR and Depend RCTs.* In the N=1,071 sample of ED SR and Depend RCT participants analyzed by Piper *et al*, Black participants endorsed lower Loss of Control and higher Tolerance scores than White participants (both $P < .05$, data not shown), as seen in the GSS dataset.[13] There were no differences in the four PDM scores overall or by sex or ethnicity between the GSS sample of N=1,864 and the N=1,071 sample analyzed by Piper *et al* (all $P > .07$, data not shown).[13]

*5.4. Nicotine Dependence Syndrome Scale, NDSS*

*Development of the NDSS.* The NDSS is a multiple factor measure of nicotine dependence developed and validated using samples of treatment-seeking smokers.[16] Initial item development was based on the Edwards and Gross dependence construct,[28] which has informed the development of diagnostic dependence criteria.[29] Treatment-seeking smokers enrolled in a research smoking cessation clinic with high motivation and efficacy to quit (N=317, minimum 10 CPD/day, mean(SD) 26(10) CPD, age 23(10) years, 57% female),[30–33] were asked to discuss and comment upon the initial list of items and their nicotine addiction. The list was further refined by addiction investigators and focus groups of smokers. A sample of treatment-seeking smokers were administered 23 provisional items along with other measures of dependence (e.g., Fagerström Tolerance Questionnaire (FTQ), withdrawal symptoms and the Horn/Russell Reasons for Smoking Scale) and smoking outcomes.[34,35] Principal component, orthogonal factor analysis and selection of items were applied to the sample data resulting in five factors labeled Drive, Priority, Tolerance, Continuity and Stereotypy. The first principal component (accounting for 29% of total item variance) was retained as an omnibus measure (NDSS-T). NDSS-T and four of five factors exhibited substantial partial correlation coefficients (PCC) with the FTQ and smoking behaviors and outcomes, e.g., PCCs ranging from 0.24-0.54 with the FTQ, 0.16-0.48 with smoking rate, and 0.16-0.44 with the Horn-Russell pharmacological reason for smoking. Many of associations persisted after correction for the FTQ or smoking rate demonstrating incremental utility.

*Refinement of the NDSS.* Further development of the NDSS utilized a sample of treatment-seeking smokers (N=802, mean(SD) 24(8) CPD, age 39(11) years, 57% female, 66% White, 31% Black), recruited to test acute effects of smoking cessation medication.[16] Additional items were added to improve the number and reliability of Continuity and Stereotypy factor items, and participants self-administered measures of addiction (e.g., FTQ), difficulty abstaining, and past severity of withdrawal. As before, principal components and orthogonal factor analysis extracted five factors, accounting for 57% of item variance. The factors Drive, Priority, Tolerance were similar to the initial study, the new Continuity and Stereotypy items loaded onto their respective factors, and Stereotypy reliability increased. As before, the first principal component (NDSS-T) and four of five factors exhibited significant association with the FTQ and smoking rate; in addition, the incremental utility and variance explained increased for difficulty abstaining and past severity of withdrawal. Analysis of variance by race identified significant differences between race (Continuity and Drive > Whites, and Stereotypy > Blacks) without differences in total score (NDSS-T).

*Test-Retest Reliability of the NDSS.* A third sample of chronic heavy smokers selected for high dependence (*N*=91, mean(SD) 36(9) CPD, age 35(9) years, 41% female, 81% White) was utilized to assess test-retest reliability after NDSS self-administration at baseline and on day 14 of a randomized clinical trial of bupropion treatment and acute abstinence.[36] No significant differences in scores overall or by factor were observed with test-retest correlations of 0.71-0.83 (NDSS-T correlation of 0.81).

*The NDSS in the GSS.* In the GSS sample, female participants endorsed higher NDSS Drive ($P < .001$), lower Continuity ($P = .03$) and lower Stereotypy ($P < .001$) scales than male participants (**Supplementary Table 9**). Black participants endorsed higher NDSS Priority and Stereotypy scores and a lower Continuity score than White participants (all $P < .001$).

*The NDSS in the ED SR and Depend RCTs.* In the N=1,071 sample of ED SR and Depend RCT participants analyzed by Piper *et al*, female participants endorsed higher NDSS Drive and Priority (both $P < .05$) scores, and lower Continuity and Stereotypy (both $P < .001$) scores than male participants; there were no NDSS score differences observed between

Black and White participants in the ED SR and Depend RCT participants.[13]

*The NDSS in the GSS and in the ED SR and Depend RCTs.* There were differences between GSS participants and ED SR and Depend RCT participants[13] in multiple NDSS scores and sex and ethnicity strata. GSS dataset participants endorsed a higher NDSS Drive ($P < .01$) score, and lower Priority, Tolerance, and Stereotypy (all $P < .001$), and Total NDSS ($P < .05$) scores than did ED SR and Depend RCT participants. Female GSS participants endorsed a higher NDSS Drive ($P < .01$) and lower Priority, Tolerance, Stereotypy (all $P < .001$) and Total ($P < .05$) scores than female participants of the ED SR and Depend RCTs. Male GSS participants endorsed a higher NDSS Drive ($P < .05$) and lower Priority and Stereotypy (both $P < .001$) scores than male participants of the ED SR and Depend RCTs. There were no differences in NDSS scores between Black GSS and Black ED SR and Dependence RCT participants. White GSS participants endorsed higher Drive ($P < .01$), and lower Priority, Tolerance, Stereotypy (all $P < .001$) and Total ($P < .01$) scores than White ED SR and Depend RCT participants.

### 6. Summary - GSS Findings

#### *6.1. Demographic Findings*

Age varies by sex (female participants younger, $P < .05$) and ethnicity (Black participants older, $P < .01$). Sex and ethnicity are associated (proportionally more female Black participants than female White participants, $P < .005$).

#### *6.2. Nicotine Dependence Findings*

*FTND.* In analysis of FTND scores as continuous scores and subscores as ordinal categories, FTND scores and subscores vary by sex and ethnicity. Female participants endorse lower total FTND scores and CPD distributions than male participants, and Black participants endorse lower CPD and higher TTFC distributions than White participants (all $P < .001$).

*TDS.* In analysis of TDS scores, female participants endorse higher TDS scores than male participants ($P < .001$), and Black participants endorse lower TDS scores than White participants ($P = .042$).

*WISDM.* In analysis of WISDM subscale and PDM scores, female participants endorse higher Automaticity, Loss of Control and PDM scores than do male participants (all $P < .05$), and Black participants endorse lower Loss of Control and higher Tolerance scores than do White participants (both $P < .05$).

*NDSS.* In analysis of NDSS scales, female participants endorsed higher Drive and lower Continuity and Stereotypy scales than do male participants (all $P < .05$), and Black participants endorse higher Priority and Stereotypy scores and lower Continuity scores than White participants (all $P < .001$).

### 7. Predicted Nicotine Biomarkers, Association with Dependence Measures

Linear regression of predicted nicotine biomarkers with the nicotine dependence measures in the UW-TTURC sample, adjusted for age, ethnicity and sex was performed and discussed in the main text (Table 4 main text). Secondary data analyses of predicted nicotine biomarkers were performed to examine potential interactions with ethnicity and sex. Significant findings were highlighted in the main text and all results are included in **Supplementary Table 10**. See **Supplementary Table 10 ReadMe** for variable definitions.

### TABLES

**Supplementary Table 1A:** Measured Nicotine Biomarkers (1° Analysis), Current Smokers, by Ethnicity and Sex

| Ethnicity | | African American | | Native Hawaiian | | White | | Latino | | Japanese American | |
|---|---|---|---|---|---|---|---|---|---|---|---|
| | | N | Mean (SE) | N | Mean (SE) | N | Mean (SE) | N | Mean (SE) | N | Mean (SE) |
| NMR* | All | 364 | 1.45 (0.04) | 311 | 1.00 (0.05) | 437 | 1.40 (0.04) | 453 | 1.52 (0.04) | 674 | 0.51 (0.03) |
| | Female | 253 | 1.48 (0.04) | 197 | 1.03 (0.05) | 247 | 1.44 (0.04) | 216 | 1.55 (0.04) | 286 | 0.55 (0.04) |
| | Male | 111 | 1.42 (0.05) | 114 | 0.97 (0.05) | 190 | 1.37 (0.04) | 237 | 1.48 (0.04) | 388 | 0.48 (0.03) |
| TNE** | All | 364 | 7.27 (0.12) | 311 | 7.36 (0.14) | 437 | 8.32 (0.11) | 453 | 6.83 (0.11) | 674 | 7.16 (0.09) |
| | Female | 253 | 7.59 (0.13) | 197 | 7.68 (0.14) | 247 | 8.62 (0.12) | 216 | 7.16 (0.13) | 286 | 7.48 (0.11) |
| | Male | 111 | 6.95 (0.14) | 114 | 7.03 (0.15) | 190 | 7.97 (0.13) | 237 | 6.51 (0.12) | 388 | 6.83 (0.10) |

*Natural log transformed, no units. **Square root transformed, nmol/mg creatinine. "All" ethnicity values are adjusted by age and sex, and ethnicity-sex strata values are adjusted by age.

**Supplementary Table 1B:** Measured Nicotine Biomarkers (2° Analysis), Current Smokers, by Ethnicity and Sex

| Ethnicity | | African American | | Native Hawaiian | | White | | Latino | | Japanese American | |
|---|---|---|---|---|---|---|---|---|---|---|---|
| | | N | Mean (SE) | N | Mean (SE) | N | Mean (SE) | N | Mean (SE) | N | Mean (SE) |
| NMR* | All | 364 | 1.44 (0.07) | 311 | 0.95 (0.07) | 437 | 1.32 (0.07) | 453 | 1.53 (0.07) | 674 | 0.48 (0.06) |
| | Female | 253 | 1.49 (0.07) | 197 | 1.00 (0.07) | 247 | 1.38 (0.07) | 216 | 1.58 (0.08) | 286 | 0.54 (0.07) |
| | Male | 111 | 1.38 (0.07) | 114 | 0.89 (0.08) | 190 | 1.27 (0.07) | 237 | 1.47 (0.07) | 388 | 0.43 (0.07) |
| TNE** | All | 364 | 6.99 (0.20) | 311 | 6.72 (0.20) | 437 | 7.42 (0.20) | 453 | 6.81 (0.19) | 674 | 6.70 (0.18) |
| | Female | 253 | 7.49 (0.20) | 197 | 7.23 (0.21) | 247 | 7.02 (0.20) | 216 | 7.32 (0.20) | 286 | 7.21 (0.19) |
| | Male | 111 | 6.48 (0.21) | 114 | 6.21 (0.21) | 190 | 6.91 (0.20) | 237 | 6.30 (0.20) | 388 | 6.19 (0.19) |

*Natural log transformed, no units. **Square root transformed, nmol/mg creatinine. "All" ethnicity values are adjusted by age, sex and CPD, and ethnicity-sex strata values are adjusted by age and CPD.

**Supplementary Table 2A. Top 200 Variants, Urinary Nicotine Metabolite Ratio, Without CPD**

| Index | SNP | CHR | POS | beta | -log10p | Allele | Functi | Symbol | Biotyp | Exon | Intron | Count |
|---|---|---|---|---|---|---|---|---|---|---|---|---|
| 1 | rs68051884 | 1 | 58324761 | -0.1668 | 4.7110 | G | intron | DAB1 | PC | - | 2/16 | 1 |
| 2 | rs10489669 | 1 | 58345238 | -0.1864 | 5.6863 | C | intron | DAB1 | PC | - | 2/16 | 36 |
| 3 | rs4912174 | 1 | 58350767 | -0.1830 | 5.3534 | G | intron | DAB1 | PC | - | 1/16 | 2 |
| 4 | rs188711835 | 1 | 229122962 | -0.1392 | 4.6167 | T | regulat | - | PFR | - | - | NA |
| 5 | rs4953085 | 2 | 44554724 | 0.1106 | 4.5806 | G | intron | PREPL | PC | - | 9/13 | 0 |
| 6 | rs9309116 | 2 | 44557919 | 0.1065 | 4.7919 | T | intron | PREPL | PC | - | 8/13 | 26 |
| 7 | rs1085487 | 2 | 44617633 | 0.1173 | 4.8128 | G | intron | CAMKMT | PC | - | 3/10 | 11 |
| 8 | rs1085489 | 2 | 44618589 | 0.1172 | 4.7649 | T | intron | CAMKMT | PC | - | 3/10 | 36 |
| 9 | rs9824170 | 3 | 29498007 | -0.1126 | 5.1041 | T | intron | RBMS3 | PC | - | 2/12 | 36 |
| 10 | rs12635474 | 3 | 131490919 | -0.2036 | 4.5065 | A | intron | CPNE4 | PC | - | 2/15 | NA |
| 11 | rs73222317 | 3 | 131516788 | -0.1452 | 5.0819 | A | intron | CPNE4 | PC | - | 2/15 | 34 |
| 12 | rs17295561 | 3 | 131527785 | -0.1492 | 4.4597 | A | intron | CPNE4 | PC | - | 2/15 | 6 |
| 13 | rs73222340 | 3 | 131534405 | -0.1434 | 4.9853 | C | intron | CPNE4 | PC | - | 2/15 | 6 |
| 14 | rs246842 | 5 | 131401801 | -0.1138 | 4.6226 | T | downst | IL3 | PC | - | - | 36 |
| 15 | rs3896275 | 5 | 159381526 | -0.1299 | 5.0948 | T | intron | ADRA1B | PC | - | 1/1 | 36 |
| 16 | rs2909728 | 5 | 173777372 | 0.1126 | 5.5163 | C | intronN | RP11-267A15.1 | lincRN | - | 1/3 | 36 |
| 17 | rs146821805 | 7 | 107813617 | -0.1697 | 4.4503 | A | intron | NRCAM | PC | - | 25/27 | 36 |
| 18 | rs28573305 | 8 | 59074226 | -0.1141 | 4.7699 | T | intronN | FAM110B | PT | - | 3/4 | 26 |
| 19 | rs2512413 | 8 | 98583425 | 0.2298 | 5.4646 | T | interge | - | - | - | - | NA |
| 20 | rs2512415 | 8 | 98589253 | 0.2306 | 5.4840 | G | interge | - | - | - | - | NA |
| 21 | rs73622422 | 10 | 7031340 | 0.1336 | 4.8664 | A | interge | - | - | - | - | 36 |
| 22 | rs6483899 | 11 | 23125029 | -0.1014 | 4.5938 | T | interge | - | - | - | - | 26 |
| 23 | rs67605123 | 13 | 71486708 | -0.1145 | 4.4980 | A | interge | - | - | - | - | 23 |
| 24 | rs971313 | 15 | 55027398 | -0.1574 | 5.1102 | C | interge | - | - | - | - | 36 |
| 25 | rs4889411 | 16 | 81883434 | 0.1274 | 5.2591 | A | intron | PLCG2 | PC | - | 2/32 | 36 |
| 26 | rs10411347 | 19 | 41166954 | -0.1194 | 5.8017 | T | interge | - | - | - | - | 12 |
| 27 | rs2604860 | 19 | 41167721 | -0.1206 | 5.8764 | G | downst | NUMBL | PC | - | - | 9 |
| 28 | rs2561554 | 19 | 41168196 | -0.1266 | 5.3759 | T | downst | NUMBL | PC | - | - | 0 |
| 29 | rs2561553 | 19 | 41169518 | -0.1242 | 5.3746 | A | downst | NUMBL | PC | - | - | 0 |
| 30 | rs2250994 | 19 | 41176403 | -0.1067 | 4.5321 | G | intron | NUMBL | PC | - | 8/9 | 0 |
| 31 | rs2561547 | 19 | 41180687 | -0.1207 | 5.6449 | A | intron | NUMBL | PC | - | 7/9 | 2 |
| 32 | rs2604879 | 19 | 41195279 | -0.1400 | 6.6278 | A | downst | ADCK4 | PC | - | - | NA |
| 33 | rs28493229 | 19 | 41224204 | -0.1848 | 6.4441 | A | upstrea | ADCK4 | PC | - | - | 6 |
| 34 | rs890934 | 19 | 41227968 | 0.1207 | 5.9207 | T | intron | ITPKC | PC | - | 1/6 | 0 |
| 35 | rs3745213 | 19 | 41248009 | -0.1804 | 6.3467 | T | downst | ITPKC | PC | - | - | 3 |
| 36 | rs1869710 | 19 | 41256375 | -0.1653 | 4.8998 | T | upstrea | SNRPA | PC | - | - | 0 |
| 37 | rs1455434 | 19 | 41257046 | -0.1832 | 5.3374 | A | 5'UTR | SNRPA | PC | 1/6 | - | 36 |
| 38 | rs2607416 | 19 | 41260831 | -0.1691 | 5.5767 | T | intron | SNRPA | PC | - | 1/5 | 5 |
| 39 | rs2305797 | 19 | 41269076 | 0.1372 | 6.8856 | C | intron | SNRPA | PC | - | 4/5 | 0 |
| 40 | rs2279011 | 19 | 41269288 | 0.1569 | 9.6306 | T | intron | SNRPA | PC | - | 4/5 | 0 |
| 41 | rs17713068 | 19 | 41270055 | -0.2014 | 7.6350 | G | intron | SNRPA | PC | - | 5/5 | 5 |
| 42 | rs2233152 | 19 | 41281016 | -0.2004 | 7.5355 | A | upstrea | MIA | PC | - | - | 0 |
| 43 | rs2287691 | 19 | 41286163 | -0.1591 | 6.3406 | G | downst | MIA | PC | - | - | 15 |
| 44 | rs17726258 | 19 | 41287605 | -0.1700 | 4.4637 | C | downst | MIA | PC | - | - | 0 |
| 45 | rs12973666 | 19 | 41289397 | 0.1582 | 9.8918 | C | intron | RAB4B | PC | - | 3/7 | 0 |
| 46 | rs2287692 | 19 | 41289756 | -0.2072 | 7.7858 | A | intron | RAB4B | PC | - | 4/7 | 10 |
| 47 | rs2604894 | 19 | 41292404 | 0.1093 | 4.7057 | G | intron | RAB4B | PC | - | 5/7 | 22 |
| 48 | rs7245595 | 19 | 41295859 | 0.1284 | 6.1925 | C | intron | RAB4B | PC | - | 7/7 | 0 |
| 49 | rs10403040 | 19 | 41296046 | -0.1384 | 5.1490 | A | intron | RAB4B | PC | - | 7/7 | 0 |
| 50 | rs28660501 | 19 | 41300277 | -0.1307 | 4.6997 | T | upstrea | EGLN2 | PC | - | - | 0 |
| 51 | rs7937 | 19 | 41302706 | 0.1037 | 4.7306 | A | upstrea | EGLN2 | PC | - | - | 5 |
| 52 | rs3733829 | 19 | 41310571 | 0.1338 | 6.5565 | G | intron | EGLN2 | PC | - | 2/5 | 0 |
| 53 | rs3736329 | 19 | 41313202 | 0.1356 | 6.6691 | A | intron | EGLN2 | PC | - | 4/5 | 0 |
| 54 | rs4802091 | 19 | 41315318 | 0.1361 | 6.7220 | C | downst | EGLN2 | PC | - | - | 0 |
| 55 | rs4803369 | 19 | 41315980 | 0.1349 | 6.6452 | A | downst | EGLN2 | PC | - | - | 0 |
| 56 | rs41530251 | 19 | 41316355 | 0.1296 | 5.9406 | C | downst | EGLN2 | PC | - | - | 0 |
| 57 | rs35061187 | 19 | 41316898 | 0.1571 | 8.5616 | T | downst | EGLN2 | PC | - | - | 0 |
| 58 | rs12052092 | 19 | 41318899 | 0.1330 | 6.3636 | A | downst | EGLN2 | PC | - | - | 0 |
| 59 | rs73048928 | 19 | 41323793 | 0.1454 | 7.2049 | A | downst | CYP2F2P | Unitary | - | - | 0 |
| 60 | rs10412779 | 19 | 41324131 | 0.1520 | 5.1181 | G | downst | CYP2F2P | Unitary | - | - | 0 |
| 61 | rs2545769 | 19 | 41324179 | -0.2065 | 9.0385 | A | downst | CYP2F2P | Unitary | - | - | NA |
| 62 | rs4803373 | 19 | 41326426 | 0.1222 | 5.1629 | G | intronN | CYP2F2P | Unitary | - | 5/5 | 0 |
| 63 | rs34842714 | 19 | 41328196 | 0.1609 | 7.6718 | T | NCTex | CYP2F2P | Unitary | 4/6 | - | 0 |
| 64 | rs112673025 | 19 | 41328823 | -0.2276 | 9.2411 | A | intronN | CYP2F2P | Unitary | - | 3/5 | 1 |
| 65 | rs7507400 | 19 | 41330179 | -0.1582 | 5.5579 | T | intronN | CYP2F2P | Unitary | - | 3/5 | NA |
| 66 | rs145256949 | 19 | 41332357 | 0.2661 | 13.5538 | G | intronN | CYP2F2P | Unitary | - | 1/5 | NA |
| 67 | rs11878604 | 19 | 41333284 | -0.3176 | 25.0528 | C | upstrea | CYP2F2P | Unitary | - | - | NA |

| | | | | | | | | | | | | |
|---|---|---|---|---|---|---|---|---|---|---|---|---|
| 68 | rs11881918 | 19 | 41334199 | -0.1744 | 6.1270 | A | upstrea | CYP2F2P | Unitary | - | - | 0 |
| 69 | rs7255471 | 19 | 41335066 | 0.2391 | 11.7166 | T | upstrea | CYP2F2P | Unitary | - | - | 0 |
| 70 | rs55978439 | 19 | 41336556 | 0.2018 | 6.7319 | A | upstrea | CYP2F2P | Unitary | - | - | NA |
| 71 | rs11879413 | 19 | 41337923 | -0.1781 | 6.5467 | T | intronN | - | lincRN | - | 1/1 | 0 |
| 72 | rs3865453 | 19 | 41338556 | -0.1783 | 6.5115 | T | intronN | - | lincRN | - | 1/1 | 0 |
| 73 | rs66889044 | 19 | 41338712 | 0.2043 | 10.0222 | T | intronN | - | lincRN | - | 1/1 | 36 |
| 74 | rs2258314 | 19 | 41338835 | -0.1697 | 5.2735 | T | intronN | - | lincRN | - | 1/1 | 0 |
| 75 | rs2258380 | 19 | 41338988 | 0.2039 | 6.8244 | G | intronN | - | lincRN | - | 1/1 | 0 |
| 76 | rs7247231 | 19 | 41339837 | -0.1643 | 5.1901 | A | intronN | - | lincRN | - | 1/1 | 0 |
| 77 | rs7246188 | 19 | 41339842 | -0.1696 | 5.4518 | G | intronN | - | lincRN | - | 1/1 | 6 |
| 78 | rs12459249 | 19 | 41339896 | 0.2104 | 15.9008 | C | intronN | - | lincRN | - | 1/1 | 0 |
| 79 | rs11083569 | 19 | 41340321 | 0.1918 | 12.0113 | G | intronN | - | lincRN | - | 1/1 | NA |
| 80 | rs10853742 | 19 | 41340573 | 0.2140 | 16.3835 | A | intronN | - | lincRN | - | 1/1 | 0 |
| 81 | rs12327581 | 19 | 41340579 | 0.2324 | 4.9592 | C | intronN | - | lincRN | - | 1/1 | 11 |
| 82 | rs28450491 | 19 | 41340937 | 0.2290 | 4.8071 | C | intronN | - | lincRN | - | 1/1 | 1 |
| 83 | rs11667314 | 19 | 41340983 | 0.2107 | 15.8840 | C | intronN | - | lincRN | - | 1/1 | 0 |
| 84 | rs12461964 | 19 | 41341229 | 0.1784 | 11.3959 | G | intronN | - | lincRN | - | 1/1 | 0 |
| 85 | rs12986371 | 19 | 41343698 | 0.1379 | 6.2184 | A | upstrea | CTC-490E21.10 | lincRN | - | - | 0 |
| 86 | rs76112798 | 19 | 41343700 | -0.1999 | 7.1993 | A | upstrea | CTC-490E21.10 | lincRN | - | - | 3 |
| 87 | rs35755165 | 19 | 41345989 | 0.2012 | 13.4934 | C | downst | CYP2A6 | PC | - | - | 0 |
| 88 | rs2316205 | 19 | 41346768 | 0.1911 | 12.3878 | C | downst | CYP2A6 | PC | - | - | 5 |
| 89 | rs60446182 | 19 | 41347998 | 0.2009 | 13.3932 | G | downst | CYP2A6 | PC | - | - | 0 |
| 90 | rs8192733 | 19 | 41349550 | 0.1481 | 7.2303 | C | 3'UTR | CYP2A6 | PC | 9/9 | - | 32 |
| 91 | rs7248240 | 19 | 41349640 | -0.1267 | 5.0313 | C | 3'UTR | CYP2A6 | PC | 9/9 | - | 33 |
| 92 | rs56113850 | 19 | 41353107 | 0.3979 | 47.2641 | C | intron | CYP2A6 | PC | - | 4/8 | 36 |
| 93 | rs56267346 | 19 | 41353338 | -0.2860 | 20.8119 | G | intron | CYP2A6 | PC | - | 4/8 | NA |
| 94 | rs2388868 | 19 | 41353849 | 0.1630 | 7.6254 | T | intron | CYP2A6 | PC | - | 4/8 | 0 |
| 95 | rs7250713 | 19 | 41355195 | 0.1257 | 5.2837 | C | intron | CYP2A6 | PC | - | 2/8 | 9 |
| 96 | rs28399433 | 19 | 41356379 | -0.3158 | 16.2826 | C | upstrea | CYP2A6 | PC | - | - | NA |
| 97 | rs61663607 | 19 | 41357076 | -0.2105 | 7.8568 | C | upstrea | CYP2A6 | PC | - | - | 0 |
| 98 | rs150298687 | 19 | 41357344 | 0.2117 | 12.6338 | C | upstrea | CYP2A6 | PC | - | - | 0 |
| 99 | rs57837628 | 19 | 41357910 | 0.3469 | 33.1463 | G | upstrea | CYP2A6 | PC | - | - | 0 |
| 100 | rs137939449 | 19 | 41359307 | -0.2355 | 9.1766 | G | upstrea | CYP2A6 | PC | - | - | 1 |
| 101 | rs3875149 | 19 | 41359996 | 0.2029 | 12.2998 | G | upstrea | CYP2A6 | PC | - | - | 0 |
| 102 | rs113288603 | 19 | 41362293 | -0.2392 | 9.8844 | T | intronN | CTC-490E21.12 | NMD | - | 1/3 | 32 |
| 103 | rs1496402 | 19 | 41366134 | 0.1609 | 7.9147 | A | intronN | CTC-490E21.12 | NMD | - | 1/3 | 33 |
| 104 | rs8102900 | 19 | 41366928 | 0.1944 | 11.6188 | A | intronN | CTC-490E21.12 | NMD | - | 1/3 | 0 |
| 105 | rs113029345 | 19 | 41370176 | 0.3862 | 37.7769 | C | intronN | - | NMD | - | 1/3 | 36 |
| 106 | rs12461383 | 19 | 41370338 | 0.3486 | 31.9991 | G | intronN | - | NMD | - | 1/3 | 0 |
| 107 | rs6508949 | 19 | 41371110 | -0.3074 | 21.8877 | A | intronN | - | NMD | - | 1/3 | 21 |
| 108 | rs7247903 | 19 | 41372475 | -0.3679 | 22.2197 | G | intronN | - | NMD | - | 1/3 | NA |
| 109 | rs59586387 | 19 | 41375030 | -0.3473 | 18.4526 | G | intronN | CTC-490E21.12 | NMD | - | 1/3 | NA |
| 110 | rs2892624 | 19 | 41375147 | -0.1422 | 5.5023 | T | intronN | CTC-490E21.12 | NMD | - | 1/3 | NA |
| 111 | rs11666974 | 19 | 41376000 | -0.1929 | 7.3668 | A | intronN | CTC-490E21.12 | NMD | - | 1/3 | NA |
| 112 | rs12151196 | 19 | 41376777 | 0.2668 | 15.1147 | G | downst | CYP2A7 | PC | - | - | 0 |
| 113 | rs7255037 | 19 | 41377338 | -0.2062 | 10.0813 | A | downst | CYP2A7 | PC | - | - | NA |
| 114 | rs56097499 | 19 | 41381334 | 0.2909 | 17.4870 | A | downst | CYP2A7 | PC | - | - | 0 |
| 115 | rs11083575 | 19 | 41381840 | 0.1382 | 5.9225 | C | intron | CYP2A7 | PC | - | 7/7 | 0 |
| 116 | rs11083576 | 19 | 41381868 | 0.1727 | 9.1312 | T | intron | CYP2A7 | PC | - | 7/7 | 0 |
| 117 | rs11083577 | 19 | 41382180 | 0.1710 | 10.0842 | C | intron | CYP2A7 | PC | - | 7/7 | 0 |
| 118 | rs3852869 | 19 | 41382665 | 0.1641 | 8.7002 | A | intron | CYP2A7 | PC | - | 6/7 | 0 |
| 119 | rs2261144 | 19 | 41383153 | 0.2857 | 22.0014 | G | missen | CYP2A7 | PC | 6/8 | - | 0 |
| 120 | rs2302988 | 19 | 41383344 | 0.2022 | 12.9597 | C | intron | CYP2A7 | PC | - | 5/7 | 0 |
| 121 | rs16958956 | 19 | 41383378 | 0.1891 | 10.7277 | A | intron | CYP2A7 | PC | - | 5/7 | 4 |
| 122 | rs3869579 | 19 | 41383799 | 0.2117 | 15.9110 | A | missen | CYP2A7 | PC | 5/8 | - | 0 |
| 123 | rs3822479 | 19 | 41383989 | 0.2601 | 16.5732 | T | intron | CYP2A7 | PC | - | 4/7 | 0 |
| 124 | rs2909906 | 19 | 41384084 | 0.2026 | 13.0855 | C | intron | CYP2A7 | PC | - | 4/7 | 0 |
| 125 | rs10404300 | 19 | 41384385 | 0.1954 | 12.2561 | C | intron | CYP2A7 | PC | - | 4/7 | 0 |
| 126 | rs4079367 | 19 | 41384637 | 0.2028 | 13.6247 | G | intron | CYP2A7 | PC | - | 4/7 | 0 |
| 127 | rs4079366 | 19 | 41384675 | 0.1863 | 10.3591 | A | missen | CYP2A7 | PC | 4/8 | - | 11 |
| 128 | rs4803391 | 19 | 41385434 | 0.1972 | 13.7022 | G | intron | CYP2A7 | PC | - | 3/7 | 0 |
| 129 | rs2545781 | 19 | 41385768 | 0.1932 | 11.6286 | A | intron | CYP2A7 | PC | - | 3/7 | 0 |
| 130 | rs12982314 | 19 | 41385774 | 0.1894 | 11.0994 | G | intron | CYP2A7 | PC | - | 3/7 | 0 |
| 131 | rs4142867 | 19 | 41386136 | 0.2036 | 14.4818 | C | missen | CYP2A7 | PC | 3/8 | - | 0 |
| 132 | rs3815704 | 19 | 41386209 | 0.1879 | 11.2289 | G | intron | CYP2A7 | PC | - | 2/7 | 0 |
| 133 | rs3815705 | 19 | 41386282 | 0.2606 | 19.9469 | A | intron | CYP2A7 | PC | - | 2/7 | 0 |
| 134 | rs56081734 | 19 | 41386420 | 0.2023 | 14.2691 | C | missen | CYP2A7 | PC | 2/8 | - | 0 |
| 135 | rs3815709 | 19 | 41386487 | 0.1806 | 10.0442 | C | missen | CYP2A7 | PC | 2/8 | - | NA |
| 136 | rs3815710 | 19 | 41386494 | 0.1808 | 9.9813 | C | missen | CYP2A7 | PC | 2/8 | - | NA |

| | | | | | | | | | | | | |
|---|---|---|---|---|---|---|---|---|---|---|---|---|
| 137 | rs58798281 | 19 | 41386527 | 0.1950 | 11.8543 | A | missen | CYP2A7 | PC | 2/8 | - | 0 |
| 138 | rs74219554 | 19 | 41386547 | 0.1945 | 12.3040 | C | intron | CYP2A7 | PC | - | 1/7 | 0 |
| 139 | rs61482096 | 19 | 41386622 | 0.1742 | 10.7435 | G | intron | CYP2A7 | PC | - | 1/7 | 0 |
| 140 | rs11669113 | 19 | 41386891 | 0.1850 | 11.5494 | G | intron | CYP2A7 | PC | - | 1/7 | 0 |
| 141 | rs10424834 | 19 | 41387241 | 0.1864 | 11.8486 | A | intron | CYP2A7 | PC | - | 1/7 | 0 |
| 142 | rs10425037 | 19 | 41387300 | 0.1831 | 11.1081 | A | intron | CYP2A7 | PC | - | 1/7 | 0 |
| 143 | rs10425169 | 19 | 41387647 | 0.1876 | 10.5336 | G | intron | CYP2A7 | PC | - | 1/7 | NA |
| 144 | rs10425185 | 19 | 41387669 | 0.1568 | 7.6664 | G | intron | CYP2A7 | PC | - | 1/7 | 0 |
| 145 | rs4803393 | 19 | 41388575 | 0.1794 | 10.8752 | G | 5'UTR | CYP2A7 | PC | 1/8 | - | 0 |
| 146 | rs3797218 | 19 | 41388707 | 0.2056 | 14.4535 | C | upstrea | CYP2A7 | PC | - | - | 0 |
| 147 | rs28602288 | 19 | 41388740 | 0.2129 | 15.6018 | T | upstrea | CYP2A7 | PC | - | - | 0 |
| 148 | rs28427254 | 19 | 41388777 | 0.1998 | 13.1773 | G | upstrea | CYP2A7 | PC | - | - | 0 |
| 149 | rs11083581 | 19 | 41388949 | 0.2164 | 14.9079 | T | upstrea | CYP2A7 | PC | - | - | 36 |
| 150 | rs4802094 | 19 | 41389509 | 0.1438 | 6.5862 | G | upstrea | CYP2A7 | PC | - | - | 0 |
| 151 | rs7253657 | 19 | 41389514 | 0.1550 | 7.7281 | C | upstrea | CYP2A7 | PC | - | - | 0 |
| 152 | rs4802095 | 19 | 41389625 | 0.1699 | 9.9865 | T | upstrea | CYP2A7 | PC | - | - | 0 |
| 153 | rs144070863 | 19 | 41390051 | 0.1963 | 11.1282 | G | upstrea | CYP2A7 | PC | - | - | NA |
| 154 | rs148634519 | 19 | 41390153 | 0.1546 | 6.9424 | G | upstrea | CYP2A7 | PC | - | - | NA |
| 155 | rs10423695 | 19 | 41392141 | 0.2070 | 13.5747 | C | upstrea | CYP2G1P | PT | - | - | 5 |
| 156 | rs3875159 | 19 | 41392154 | 0.2008 | 12.9903 | C | upstrea | CYP2G1P | PT | - | - | 0 |
| 157 | rs10423165 | 19 | 41392466 | 0.1910 | 12.6841 | C | upstrea | CYP2G1P | PT | - | - | 0 |
| 158 | rs3909341 | 19 | 41393326 | 0.1995 | 13.7970 | A | upstrea | CYP2G1P | PT | - | - | 0 |
| 159 | rs5007415 | 19 | 41393760 | 0.2090 | 15.2886 | A | upstrea | CYP2G1P | PT | - | - | 0 |
| 160 | rs67421541 | 19 | 41394420 | 0.1988 | 13.7022 | T | upstrea | CYP2G1P | PT | - | - | 0 |
| 161 | rs10418318 | 19 | 41395036 | 0.2026 | 14.2116 | A | upstrea | CYP2G1P | PT | - | - | 0 |
| 162 | rs28472879 | 19 | 41395755 | 0.2087 | 15.1610 | A | upstrea | CYP2G1P | PT | - | - | 0 |
| 163 | rs4803397 | 19 | 41396865 | 0.1752 | 9.5647 | A | NCTex | CYP2G1P | PT | 1/5 | - | 0 |
| 164 | rs8103444 | 19 | 41397661 | 0.1910 | 11.6803 | A | NCTex | CYP2G1P | PT | 1/5 | - | 0 |
| 165 | rs78374326 | 19 | 41398651 | 0.2550 | 18.6542 | C | intronN | CYP2G1P | PT | - | 1/4 | NA |
| 166 | rs6508953 | 19 | 41402579 | 0.1572 | 8.6179 | C | intronN | CYP2G1P | PT | - | 2/4 | 0 |
| 167 | rs7252852 | 19 | 41403325 | 0.2095 | 15.4066 | C | intronN | CYP2G1P | PT | - | 2/4 | 0 |
| 168 | rs10419393 | 19 | 41404196 | 0.2112 | 15.6391 | C | NCTex | CYP2G1P | PT | 3/5 | - | 0 |
| 169 | rs4803400 | 19 | 41405962 | 0.2048 | 14.7604 | C | NCTex | CYP2G1P | PT | 5/5 | - | 0 |
| 170 | rs7254188 | 19 | 41407343 | 0.1999 | 14.1048 | A | downst | CYP2G1P | PT | - | - | 0 |
| 171 | rs7258590 | 19 | 41408581 | 0.2169 | 15.9197 | T | downst | CYP2G1P | PT | - | - | 0 |
| 172 | rs4803402 | 19 | 41412185 | 0.2191 | 13.5535 | A | downst | CTC-490E21.13 | UPPG | - | - | 7 |
| 173 | rs72480748 | 19 | 41414481 | 0.2881 | 23.9701 | A | NCTex | CTC-490E21.13 | UPPG | 5/5 | - | 0 |
| 174 | rs3844442 | 19 | 41415112 | 0.2092 | 15.5529 | A | intronN | CTC-490E21.13 | UPPG | - | 4/4 | 2 |
| 175 | rs3852872 | 19 | 41416143 | 0.2100 | 12.6548 | T | intronN | CTC-490E21.13 | UPPG | - | 2/4 | 0 |
| 176 | rs3852873 | 19 | 41416260 | 0.2194 | 13.4643 | A | intronN | CTC-490E21.13 | UPPG | - | 2/4 | 0 |
| 177 | rs10419589 | 19 | 41416750 | 0.2098 | 15.5135 | C | NCTex | CTC-490E21.13 | UPPG | 1/5 | - | 2 |
| 178 | rs10425738 | 19 | 41417727 | 0.2476 | 19.0880 | A | upstrea | CTC-490E21.13 | UPPG | - | - | NA |
| 179 | rs73034462 | 19 | 41418134 | 0.2399 | 18.8658 | A | upstrea | CTC-490E21.13 | UPPG | - | - | 0 |
| 180 | rs76935404 | 19 | 41419294 | 0.2635 | 22.2004 | T | upstrea | CTC-490E21.13 | UPPG | - | - | 34 |
| 181 | rs10420231 | 19 | 41420030 | 0.2015 | 11.8502 | A | upstrea | CTC-490E21.13 | UPPG | - | - | NA |
| 182 | rs8108939 | 19 | 41425149 | 0.2128 | 16.0890 | G | upstrea | CYP2B7P | RI | - | - | 36 |
| 183 | rs4001943 | 19 | 41425900 | 0.2306 | 16.3938 | C | upstrea | CYP2B7P | TUPG | - | - | 2 |
| 184 | rs10424844 | 19 | 41425959 | 0.1740 | 11.4110 | T | upstrea | CYP2B7P | TUPG | - | - | 0 |
| 185 | rs12459565 | 19 | 41427539 | 0.2192 | 13.4447 | A | upstrea | CYP2B7P | TUPG | - | - | 0 |
| 186 | rs28417358 | 19 | 41428105 | 0.2308 | 17.6375 | A | upstrea | CYP2B7P | TUPG | - | - | 0 |
| 187 | rs11083589 | 19 | 41428416 | 0.2099 | 14.2376 | G | upstrea | CYP2B7P | TUPG | - | - | NA |
| 188 | rs57274441 | 19 | 41431422 | 0.2360 | 17.6373 | G | intronN | CYP2B7P | TUPG | - | 1/8 | NA |
| 189 | rs3844443 | 19 | 41431935 | 0.2131 | 13.0003 | C | intronN | CYP2B7P | TUPG | - | 1/8 | 0 |
| 190 | rs12151139 | 19 | 41433543 | 0.2567 | 21.3516 | T | intronN | CYP2B7P | TUPG | - | 1/8 | 1 |
| 191 | rs4609955 | 19 | 41433613 | 0.2409 | 19.0239 | C | intronN | CYP2B7P | TUPG | - | 1/8 | 0 |
| 192 | rs3843043 | 19 | 41433931 | 0.2131 | 13.0003 | T | intronN | CYP2B7P | TUPG | - | 1/8 | NA |
| 193 | rs3844445 | 19 | 41434106 | 0.2188 | 14.5714 | G | intronN | CYP2B7P | TUPG | - | 1/8 | 2 |
| 194 | rs7248187 | 19 | 41437426 | 0.1764 | 10.0203 | C | intronN | CYP2B7P | TUPG | - | 1/8 | 0 |
| 195 | rs7247910 | 19 | 41437440 | 0.1789 | 10.3045 | G | intronN | CYP2B7P | TUPG | - | 1/8 | 5 |
| 196 | rs6508960 | 19 | 41437717 | 0.1639 | 8.6159 | A | intronN | CYP2B7P | TUPG | - | 1/8 | 0 |
| 197 | rs73038469 | 19 | 41442597 | 0.2579 | 16.9787 | A | intronN | CYP2B7P | TUPG | - | 3/8 | 0 |
| 198 | rs112819506 | 19 | 41444725 | -0.2467 | 4.5650 | G | intronN | CYP2B7P | TUPG | - | 3/8 | 0 |
| 199 | rs56401945 | 19 | 41463690 | -0.2662 | 5.2738 | C | interge | - | - | - | - | 6 |
| 200 | rs16974790 | 19 | 41498946 | -0.2291 | 4.4749 | A | intron | CYP2B6 | PC | - | 1/8 | 4 |

**Supplementary Table 2B. Top 200 Variants, Urinary Nicotine Metabolite Ratio, With CPD**

| Index | SNP | CHR | POS | beta | -log10p | Allele | Consequence | Symbol | Biotype | Exon | Intron | Count |
|---|---|---|---|---|---|---|---|---|---|---|---|---|
| 1 | rs68051884 | 1 | 58324761 | -0.1702 | 4.9417 | G | intron | DAB1 | PC | - | 2/16 | 1 |
| 2 | rs10489669 | 1 | 58345238 | -0.1943 | 6.1987 | C | intron | DAB1 | PC | - | 2/16 | 36 |
| 3 | rs4912174 | 1 | 58350767 | -0.1891 | 5.7473 | G | intron | DAB1 | PC | - | 1/16 | 1 |
| 4 | rs188711835 | 1 | 229122962 | -0.1366 | 4.5259 | T | regulatory | - | PFR | - | - | NA |
| 5 | rs4953085 | 2 | 44554724 | 0.1115 | 4.7075 | G | intron | PREPL | PC | - | 9/13 | 0 |
| 6 | rs9309116 | 2 | 44557919 | 0.1081 | 4.9957 | T | intron | PREPL | PC | - | 8/13 | 28 |
| 7 | rs1085487 | 2 | 44617633 | 0.1200 | 5.0745 | G | intron | CAMKMT | PC | - | 3/10 | 6 |
| 8 | rs1085489 | 2 | 44618589 | 0.1206 | 5.0817 | T | intron | CAMKMT | PC | - | 3/10 | 36 |
| 9 | rs1067368 | 2 | 44654889 | 0.1107 | 4.8832 | A | intron | CAMKMT | PC | - | 3/10 | 0 |
| 10 | rs9824170 | 3 | 29498007 | -0.1146 | 5.3359 | T | intron | RBMS3 | PC | - | 2/12 | 36 |
| 11 | rs73222317 | 3 | 131516788 | -0.1429 | 5.0132 | A | intron | CPNE4 | PC | - | 2/15 | 34 |
| 12 | rs17295561 | 3 | 131527785 | -0.1473 | 4.4231 | A | intron | CPNE4 | PC | - | 2/15 | 6 |
| 13 | rs73222340 | 3 | 131534405 | -0.1416 | 4.9462 | C | intron | CPNE4 | PC | - | 2/15 | 6 |
| 14 | rs246842 | 5 | 131401801 | -0.1148 | 4.7560 | T | downstream | IL3 | PC | - | - | 28 |
| 15 | rs168681 | 5 | 131402450 | -0.1021 | 4.5029 | G | downstream | IL3 | PC | - | - | 12 |
| 16 | rs3896275 | 5 | 159381526 | -0.1214 | 4.5725 | T | intron | ADRA1B | PC | - | 1/1 | 36 |
| 17 | rs2909728 | 5 | 173777372 | 0.1115 | 5.4999 | C | intronNCT | RP11-267A15.1 | lincRNA | - | 1/3 | 36 |
| 18 | rs146821805 | 7 | 107813617 | -0.1669 | 4.3838 | A | intron | NRCAM | PC | - | 25/27 | 36 |
| 19 | rs28573305 | 8 | 59074226 | -0.1203 | 5.3060 | T | intronNCT | FAM110B | PT | - | 3/4 | 29 |
| 20 | rs2512413 | 8 | 98583425 | 0.2281 | 5.4694 | T | intergenic | - | - | - | - | NA |
| 21 | rs2512415 | 8 | 98589253 | 0.2283 | 5.4644 | G | intergenic | - | - | - | - | NA |
| 22 | rs73622422 | 10 | 7031340 | 0.1343 | 4.9834 | A | intergenic | - | - | - | - | 36 |
| 23 | rs67605123 | 13 | 71486708 | -0.1139 | 4.5186 | A | intergenic | - | - | - | - | 26 |
| 24 | rs4243579 | 14 | 52161021 | -0.1028 | 4.5326 | C | intron | FRMD6 | PC | - | 2/13 | 30 |
| 25 | rs971313 | 15 | 55027398 | -0.1504 | 4.7693 | C | intergenic | - | - | - | - | 36 |
| 26 | rs4889411 | 16 | 81883434 | 0.1234 | 5.0362 | A | intron | PLCG2 | PC | - | 2/1 | 36 |
| 27 | rs10411347 | 19 | 41166954 | -0.1139 | 5.3968 | T | intergenic | - | - | - | - | 9 |
| 28 | rs2604860 | 19 | 41167721 | -0.1147 | 5.4444 | G | downstream | NUMBL | PC | - | - | 10 |
| 29 | rs2561554 | 19 | 41168196 | -0.1251 | 5.3369 | T | downstream | NUMBL | PC | - | - | 0 |
| 30 | rs2561553 | 19 | 41169518 | -0.1224 | 5.3095 | A | downstream | NUMBL | PC | - | - | 0 |
| 31 | rs2561547 | 19 | 41180687 | -0.1147 | 5.2173 | A | intron | NUMBL | PC | - | 7/9 | 1 |
| 32 | rs2604879 | 19 | 41195279 | -0.1348 | 6.2629 | A | downstream | ADCK4 | PC | - | - | NA |
| 33 | rs28493229 | 19 | 41224204 | -0.1778 | 6.0771 | A | upstream | ADCK4 | PC | - | - | 4 |
| 34 | rs890934 | 19 | 41227968 | 0.1138 | 5.3871 | T | intron | ITPKC | PC | - | 1/6 | 0 |
| 35 | rs3745213 | 19 | 41248009 | -0.1741 | 6.0267 | T | downstream | ITPKC | PC | - | - | 0 |
| 36 | rs1869710 | 19 | 41256375 | -0.1619 | 4.7866 | T | upstream | SNRPA | PC | - | - | 0 |
| 37 | rs1455434 | 19 | 41257046 | -0.1796 | 5.2224 | A | 5'UTR | SNRPA | PC | 1/6 | - | 36 |
| 38 | rs2607416 | 19 | 41260831 | -0.1622 | 5.2428 | T | intron | SNRPA | PC | - | 1/5 | 4 |
| 39 | rs2305797 | 19 | 41269076 | 0.1297 | 6.2793 | C | intron | SNRPA | PC | - | 4/5 | 0 |
| 40 | rs2279011 | 19 | 41269288 | 0.1506 | 9.0138 | T | intron | SNRPA | PC | - | 4/5 | 0 |
| 41 | rs17713068 | 19 | 41270055 | -0.1936 | 7.1892 | G | intron | SNRPA | PC | - | 5/5 | 4 |
| 42 | rs2233152 | 19 | 41281016 | -0.1927 | 7.1015 | A | upstream | MIA | PC | - | - | 0 |
| 43 | rs2287691 | 19 | 41286163 | -0.1526 | 5.9555 | G | downstream | MIA | PC | - | - | 7 |
| 44 | rs12973666 | 19 | 41289397 | 0.1526 | 9.3421 | C | intron | RAB4B | PC | - | 3/7 | 0 |
| 45 | rs2287692 | 19 | 41289756 | -0.1991 | 7.3267 | A | intron | RAB4B | PC | - | 4/7 | 14 |
| 46 | rs2604894 | 19 | 41292404 | 0.1046 | 4.3962 | G | intron | RAB4B | PC | - | 5/7 | 21 |
| 47 | rs7245595 | 19 | 41295859 | 0.1211 | 5.6246 | C | intron | RAB4B | PC | - | 7/7 | 1 |
| 48 | rs10403040 | 19 | 41296046 | -0.1318 | 4.7825 | A | intron | RAB4B | PC | - | 7/7 | 0 |
| 49 | rs28660501 | 19 | 41300277 | -0.1248 | 4.3908 | T | upstream | EGLN2 | PC | - | - | 0 |
| 50 | rs7937 | 19 | 41302706 | 0.0985 | 4.3583 | A | upstream | EGLN2 | PC | - | - | 4 |
| 51 | rs3733829 | 19 | 41310571 | 0.1293 | 6.2365 | G | intron | EGLN2 | PC | - | 2/5 | 0 |
| 52 | rs3736329 | 19 | 41313202 | 0.1310 | 6.3363 | A | intron | EGLN2 | PC | - | 4/5 | 0 |
| 53 | rs4802091 | 19 | 41315318 | 0.1315 | 6.3895 | C | downstream | EGLN2 | PC | - | - | 0 |
| 54 | rs4803369 | 19 | 41315980 | 0.1303 | 6.3063 | A | downstream | EGLN2 | PC | - | - | 0 |
| 55 | rs41530251 | 19 | 41316355 | 0.1249 | 5.6284 | C | downstream | EGLN2 | PC | - | - | 0 |
| 56 | rs35061187 | 19 | 41316898 | 0.1519 | 8.1457 | T | downstream | EGLN2 | PC | - | - | 0 |
| 57 | rs12052092 | 19 | 41318899 | 0.1280 | 6.0067 | A | downstream | EGLN2 | PC | - | - | 0 |
| 58 | rs73048928 | 19 | 41323793 | 0.1407 | 6.8683 | A | downstream | CYP2F2P | UnitaryPG | - | - | 0 |
| 59 | rs10412779 | 19 | 41324131 | 0.1565 | 5.4629 | G | downstream | CYP2F2P | UnitaryPG | - | - | 0 |
| 60 | rs2545769 | 19 | 41324179 | -0.2098 | 9.4495 | A | downstream | CYP2F2P | UnitaryPG | - | - | NA |
| 61 | rs4803373 | 19 | 41326426 | 0.1200 | 5.0590 | G | intronNCT | CYP2F2P | UnitaryPG | - | 5/5 | 0 |
| 62 | rs34842714 | 19 | 41328196 | 0.1563 | 7.3704 | T | NCTexon | CYP2F2P | UnitaryPG | 4/6 | - | 0 |
| 63 | rs112673025 | 19 | 41328823 | -0.2231 | 9.0393 | A | intronNCT | CYP2F2P | UnitaryPG | - | 3/5 | 1 |
| 64 | rs7507400 | 19 | 41330179 | -0.1647 | 6.0630 | T | intronNCT | CYP2F2P | UnitaryPG | - | 3/5 | NA |
| 65 | rs145256949 | 19 | 41332357 | 0.2620 | 13.3434 | G | intronNCT | CYP2F2P | UnitaryPG | - | 1/5 | NA |
| 66 | rs11878604 | 19 | 41333284 | -0.3093 | 24.0543 | C | upstream | CYP2F2P | UnitaryPG | - | - | NA |
| 67 | rs11881918 | 19 | 41334199 | -0.1722 | 6.0611 | A | upstream | CYP2F2P | UnitaryPG | - | - | 0 |

| | | | | | | | | | | | | |
|---|---|---|---|---|---|---|---|---|---|---|---|---|
| 68 | rs7255471 | 19 | 41335066 | 0.2354 | 11.5386 | T | upstream | CYP2F2P | UnitaryPG | - | - | 0 |
| 69 | rs55978439 | 19 | 41336556 | 0.1976 | 6.5704 | A | upstream | CYP2F2P | UnitaryPG | - | - | NA |
| 70 | rs11879413 | 19 | 41337923 | -0.1762 | 6.4987 | T | intronNCT | - | lincRNA | - | 1/1 | 0 |
| 71 | rs3865453 | 19 | 41338556 | -0.1763 | 6.4609 | T | intronNCT | - | lincRNA | - | 1/1 | 0 |
| 72 | rs66889044 | 19 | 41338712 | 0.2000 | 9.7668 | T | intronNCT | - | lincRNA | - | 1/1 | 36 |
| 73 | rs2258314 | 19 | 41338835 | -0.1693 | 5.3175 | T | intronNCT | - | lincRNA | - | 1/1 | 0 |
| 74 | rs2258380 | 19 | 41338988 | 0.1993 | 6.6381 | G | intronNCT | - | lincRNA | - | 1/1 | 0 |
| 75 | rs7247231 | 19 | 41339837 | -0.1640 | 5.2441 | A | intronNCT | - | lincRNA | - | 1/1 | 0 |
| 76 | rs7246188 | 19 | 41339842 | -0.1684 | 5.4539 | G | intronNCT | - | lincRNA | - | 1/1 | 5 |
| 77 | rs12459249 | 19 | 41339896 | 0.2044 | 15.2171 | C | intronNCT | - | lincRNA | - | 1/1 | 0 |
| 78 | rs11083569 | 19 | 41340321 | 0.1870 | 11.5882 | G | intronNCT | - | lincRNA | - | 1/1 | NA |
| 79 | rs10853742 | 19 | 41340573 | 0.2081 | 15.7112 | A | intronNCT | - | lincRNA | - | 1/1 | 0 |
| 80 | rs12327581 | 19 | 41340579 | 0.2323 | 5.0297 | C | intronNCT | - | lincRNA | - | 1/1 | 15 |
| 81 | rs28450491 | 19 | 41340937 | 0.2289 | 4.8750 | C | intronNCT | - | lincRNA | - | 1/1 | 1 |
| 82 | rs11667314 | 19 | 41340983 | 0.2046 | 15.2022 | C | intronNCT | - | lincRNA | - | 1/1 | 0 |
| 83 | rs12461964 | 19 | 41341229 | 0.1733 | 10.9315 | G | intronNCT | - | lincRNA | - | 1/1 | 0 |
| 84 | rs12986371 | 19 | 41343698 | 0.1318 | 5.8068 | A | upstream | CTC-490E21.10 | lincRNA | - | - | 0 |
| 85 | rs76112798 | 19 | 41343700 | -0.1998 | 7.2862 | A | upstream | CTC-490E21.10 | lincRNA | - | - | 3 |
| 86 | rs35755165 | 19 | 41345989 | 0.1956 | 12.9562 | C | downstream | CYP2A6 | PC | - | - | 0 |
| 87 | rs2316205 | 19 | 41346768 | 0.1860 | 11.9134 | C | downstream | CYP2A6 | PC | - | - | 5 |
| 88 | rs60446182 | 19 | 41347998 | 0.1962 | 12.9788 | G | downstream | CYP2A6 | PC | - | - | 0 |
| 89 | rs8192733 | 19 | 41349550 | 0.1468 | 7.2193 | C | 3'UTR | CYP2A6 | PC | 9/9 | - | 30 |
| 90 | rs7248240 | 19 | 41349640 | -0.1218 | 4.7501 | C | 3'UTR | CYP2A6 | PC | 9/9 | - | 34 |
| 91 | rs56113850 | 19 | 41353107 | 0.3897 | 45.8266 | C | intron | CYP2A6 | PC | - | 4/8 | 36 |
| 92 | rs56267346 | 19 | 41353338 | -0.2828 | 20.6607 | G | intron | CYP2A6 | PC | - | 4/8 | NA |
| 93 | rs2388868 | 19 | 41353849 | 0.1528 | 6.8501 | T | intron | CYP2A6 | PC | - | 4/8 | 0 |
| 94 | rs7250713 | 19 | 41355195 | 0.1199 | 4.9186 | C | intron | CYP2A6 | PC | - | 2/8 | 14 |
| 95 | rs28399433 | 19 | 41356379 | -0.3114 | 16.0832 | C | upstream | CYP2A6 | PC | - | - | NA |
| 96 | rs61663607 | 19 | 41357076 | -0.2002 | 7.2552 | C | upstream | CYP2A6 | PC | - | - | 0 |
| 97 | rs150298687 | 19 | 41357344 | 0.2060 | 12.1609 | C | upstream | CYP2A6 | PC | - | - | 0 |
| 98 | rs57837628 | 19 | 41357910 | 0.3388 | 32.0043 | G | upstream | CYP2A6 | PC | - | - | 0 |
| 99 | rs137939449 | 19 | 41359307 | -0.2258 | 8.5950 | G | upstream | CYP2A6 | PC | - | - | 1 |
| 100 | rs3875149 | 19 | 41359996 | 0.1969 | 11.7802 | G | upstream | CYP2A6 | PC | - | - | 0 |
| 101 | rs113288603 | 19 | 41362293 | -0.2289 | 9.2214 | T | intronNMD | CTC-490E21.12 | NMD | - | 1/3 | 35 |
| 102 | rs1496402 | 19 | 41366134 | 0.1553 | 7.5130 | A | intronNMD | CTC-490E21.12 | NMD | - | 1/3 | 33 |
| 103 | rs8102900 | 19 | 41366928 | 0.1882 | 11.0833 | A | intronNMD | CTC-490E21.12 | NMD | - | 1/3 | 0 |
| 104 | rs113029345 | 19 | 41370176 | 0.3786 | 36.7468 | C | intronNMD | - | NMD | - | 1/3 | 36 |
| 105 | rs12461383 | 19 | 41370338 | 0.3419 | 31.1728 | G | intronNMD | - | NMD | - | 1/3 | 0 |
| 106 | rs6508949 | 19 | 41371110 | -0.2999 | 21.1319 | A | intronNMD | - | NMD | - | 1/3 | 14 |
| 107 | rs7247903 | 19 | 41372475 | -0.3555 | 21.0294 | G | intronNMD | - | NMD | - | 1/3 | NA |
| 108 | rs59586387 | 19 | 41375030 | -0.3369 | 17.6249 | G | intronNMD | CTC-490E21.12 | NMD | - | 1/3 | NA |
| 109 | rs2892624 | 19 | 41375147 | -0.1406 | 5.4701 | T | intronNMD | CTC-490E21.12 | NMD | - | 1/3 | NA |
| 110 | rs11666974 | 19 | 41376000 | -0.1834 | 6.7975 | A | intronNMD | CTC-490E21.12 | NMD | - | 1/3 | NA |
| 111 | rs12151196 | 19 | 41376777 | 0.2567 | 14.2025 | G | downstream | CYP2A7 | PC | - | - | 1 |
| 112 | rs7255037 | 19 | 41377338 | -0.1980 | 9.4600 | A | downstream | CYP2A7 | PC | - | - | NA |
| 113 | rs56097499 | 19 | 41381334 | 0.2793 | 16.3356 | A | downstream | CYP2A7 | PC | - | - | 0 |
| 114 | rs11083575 | 19 | 41381840 | 0.1327 | 5.5764 | C | intron | CYP2A7 | PC | - | 7/7 | 0 |
| 115 | rs11083576 | 19 | 41381868 | 0.1675 | 8.7454 | T | intron | CYP2A7 | PC | - | 7/7 | 0 |
| 116 | rs11083577 | 19 | 41382180 | 0.1650 | 9.5534 | C | intron | CYP2A7 | PC | - | 7/7 | 0 |
| 117 | rs3852869 | 19 | 41382665 | 0.1629 | 8.7069 | A | intron | CYP2A7 | PC | - | 6/7 | 0 |
| 118 | rs2261144 | 19 | 41383153 | 0.2739 | 20.4644 | G | missense | CYP2A7 | PC | 6/8 | - | 0 |
| 119 | rs2302988 | 19 | 41383344 | 0.1999 | 12.8685 | C | intron | CYP2A7 | PC | - | 5/7 | 0 |
| 120 | rs16958956 | 19 | 41383378 | 0.1871 | 10.6827 | A | intron | CYP2A7 | PC | - | 5/7 | 4 |
| 121 | rs3869579 | 19 | 41383799 | 0.2048 | 15.1278 | A | missense | CYP2A7 | PC | 5/8 | - | 0 |
| 122 | rs3822479 | 19 | 41383989 | 0.2465 | 15.0783 | T | intron | CYP2A7 | PC | - | 4/7 | 0 |
| 123 | rs2909906 | 19 | 41384084 | 0.1983 | 12.7381 | C | intron | CYP2A7 | PC | - | 4/7 | 0 |
| 124 | rs10404300 | 19 | 41384385 | 0.1913 | 11.9497 | C | intron | CYP2A7 | PC | - | 4/7 | 0 |
| 125 | rs4079367 | 19 | 41384637 | 0.1965 | 12.9994 | G | intron | CYP2A7 | PC | - | 4/7 | 0 |
| 126 | rs4079366 | 19 | 41384675 | 0.1834 | 10.2069 | A | missense | CYP2A7 | PC | 4/8 | - | 7 |
| 127 | rs4803391 | 19 | 41385434 | 0.1904 | 12.9846 | G | intron | CYP2A7 | PC | - | 3/7 | 0 |
| 128 | rs2545781 | 19 | 41385768 | 0.1881 | 11.2076 | A | intron | CYP2A7 | PC | - | 3/7 | 0 |
| 129 | rs12982314 | 19 | 41385774 | 0.1854 | 10.8252 | G | intron | CYP2A7 | PC | - | 3/7 | 0 |
| 130 | rs4142867 | 19 | 41386136 | 0.1967 | 13.7409 | C | missense | CYP2A7 | PC | 3/8 | - | 0 |
| 131 | rs3815704 | 19 | 41386209 | 0.1789 | 10.3648 | G | intron | CYP2A7 | PC | - | 2/7 | 0 |
| 132 | rs3815705 | 19 | 41386282 | 0.2513 | 18.8198 | A | intron | CYP2A7 | PC | - | 2/7 | 0 |
| 133 | rs56081734 | 19 | 41386420 | 0.1954 | 13.5343 | C | missense | CYP2A7 | PC | 2/8 | - | 0 |
| 134 | rs3815709 | 19 | 41386487 | 0.1736 | 9.4545 | C | missense | CYP2A7 | PC | 2/8 | - | NA |
| 135 | rs3815710 | 19 | 41386494 | 0.1732 | 9.3286 | C | missense | CYP2A7 | PC | 2/8 | - | NA |
| 136 | rs58798281 | 19 | 41386527 | 0.1874 | 11.1454 | A | missense | CYP2A7 | PC | 2/8 | - | 0 |

| | | | | | | | | | | | | |
|---|---|---|---|---|---|---|---|---|---|---|---|---|
| 137 | rs74219554 | 19 | 41386547 | 0.1868 | 11.5448 | C | intron | CYP2A7 | PC | - | 1/7 | 0 |
| 138 | rs61482096 | 19 | 41386622 | 0.1679 | 10.1650 | G | intron | CYP2A7 | PC | - | 1/7 | 0 |
| 139 | rs11669113 | 19 | 41386891 | 0.1776 | 10.8346 | G | intron | CYP2A7 | PC | - | 1/7 | 0 |
| 140 | rs10424834 | 19 | 41387241 | 0.1784 | 11.0390 | A | intron | CYP2A7 | PC | - | 1/7 | 0 |
| 141 | rs10425037 | 19 | 41387300 | 0.1748 | 10.3056 | A | intron | CYP2A7 | PC | - | 1/7 | 0 |
| 142 | rs10425169 | 19 | 41387647 | 0.1785 | 9.7160 | G | intron | CYP2A7 | PC | - | 1/7 | NA |
| 143 | rs10425185 | 19 | 41387669 | 0.1557 | 7.6810 | G | intron | CYP2A7 | PC | - | 1/7 | 0 |
| 144 | rs4803393 | 19 | 41388575 | 0.1732 | 10.3229 | G | 5'UTR | CYP2A7 | PC | 1/8 | - | 0 |
| 145 | rs3797218 | 19 | 41388707 | 0.1987 | 13.7188 | C | upstream | CYP2A7 | PC | - | - | 0 |
| 146 | rs28602288 | 19 | 41388740 | 0.2104 | 15.4845 | T | upstream | CYP2A7 | PC | - | - | 0 |
| 147 | rs28427254 | 19 | 41388777 | 0.1939 | 12.6142 | G | upstream | CYP2A7 | PC | - | - | 0 |
| 148 | rs11083581 | 19 | 41388949 | 0.2146 | 14.8915 | T | upstream | CYP2A7 | PC | - | - | 36 |
| 149 | rs4802094 | 19 | 41389509 | 0.1371 | 6.1199 | G | upstream | CYP2A7 | PC | - | - | 0 |
| 150 | rs7253657 | 19 | 41389514 | 0.1516 | 7.5158 | C | upstream | CYP2A7 | PC | - | - | 0 |
| 151 | rs4802095 | 19 | 41389625 | 0.1647 | 9.5546 | T | upstream | CYP2A7 | PC | - | - | 0 |
| 152 | rs144070863 | 19 | 41390051 | 0.1923 | 10.8596 | G | upstream | CYP2A7 | PC | - | - | NA |
| 153 | rs148634519 | 19 | 41390153 | 0.1489 | 6.5661 | G | upstream | CYP2A7 | PC | - | - | NA |
| 154 | rs10423695 | 19 | 41392141 | 0.2004 | 12.9250 | C | upstream | CYP2G1P | PT | - | - | 5 |
| 155 | rs3875159 | 19 | 41392154 | 0.1958 | 12.5641 | C | upstream | CYP2G1P | PT | - | - | 0 |
| 156 | rs10423165 | 19 | 41392466 | 0.1853 | 12.1298 | C | upstream | CYP2G1P | PT | - | - | 0 |
| 157 | rs3909341 | 19 | 41393326 | 0.1936 | 13.2133 | A | upstream | CYP2G1P | PT | - | - | 0 |
| 158 | rs5007415 | 19 | 41393760 | 0.2031 | 14.6586 | A | upstream | CYP2G1P | PT | - | - | 0 |
| 159 | rs67421541 | 19 | 41394420 | 0.1931 | 13.1344 | T | upstream | CYP2G1P | PT | - | - | 0 |
| 160 | rs10418318 | 19 | 41395036 | 0.1969 | 13.6408 | A | upstream | CYP2G1P | PT | - | - | 0 |
| 161 | rs28472879 | 19 | 41395755 | 0.2029 | 14.5483 | A | upstream | CYP2G1P | PT | - | - | 0 |
| 162 | rs4803397 | 19 | 41396865 | 0.1742 | 9.6108 | A | NCTexon | CYP2G1P | PT | 1/5 | - | 0 |
| 163 | rs8103444 | 19 | 41397661 | 0.1895 | 11.6836 | A | NCTexon | CYP2G1P | PT | 1/5 | - | 0 |
| 164 | rs78374326 | 19 | 41398651 | 0.2457 | 17.5471 | C | intronNCT | CYP2G1P | PT | - | 1/4 | NA |
| 165 | rs6508953 | 19 | 41402579 | 0.1529 | 8.3068 | C | intronNCT | CYP2G1P | PT | - | 2/4 | 0 |
| 166 | rs7252852 | 19 | 41403325 | 0.2034 | 14.7570 | C | intronNCT | CYP2G1P | PT | - | 2/4 | 0 |
| 167 | rs10419393 | 19 | 41404196 | 0.2051 | 14.9791 | C | NCTexon | CYP2G1P | PT | 3/5 | - | 0 |
| 168 | rs4803400 | 19 | 41405962 | 0.1989 | 14.1535 | C | NCTexon | CYP2G1P | PT | 5/5 | - | 0 |
| 169 | rs7254188 | 19 | 41407343 | 0.1943 | 13.5384 | A | downstream | CYP2G1P | PT | - | - | 0 |
| 170 | rs7258590 | 19 | 41408581 | 0.2093 | 15.0416 | T | downstream | CYP2G1P | PT | - | - | 0 |
| 171 | rs4803402 | 19 | 41412185 | 0.2121 | 12.9114 | A | downstream | CTC-490E21.13 | UPPG | - | - | 6 |
| 172 | rs72480748 | 19 | 41414481 | 0.2779 | 22.5549 | A | NCTexon | CTC-490E21.13 | UPPG | 5/5 | - | 0 |
| 173 | rs3844442 | 19 | 41415112 | 0.2083 | 15.6682 | A | intronNCT | CTC-490E21.13 | UPPG | - | 4/4 | 3 |
| 174 | rs3852872 | 19 | 41416143 | 0.2085 | 12.6794 | T | intronNCT | CTC-490E21.13 | UPPG | - | 2/4 | 0 |
| 175 | rs3852873 | 19 | 41416260 | 0.2182 | 13.5158 | A | intronNCT | CTC-490E21.13 | UPPG | - | 2/4 | 0 |
| 176 | rs10419589 | 19 | 41416750 | 0.2073 | 15.3887 | C | NCTexon | CTC-490E21.13 | UPPG | 1/5 | - | 2 |
| 177 | rs10425738 | 19 | 41417727 | 0.2465 | 19.2204 | A | upstream | CTC-490E21.13 | UPPG | - | - | NA |
| 178 | rs73034462 | 19 | 41418134 | 0.2327 | 18.0183 | A | upstream | CTC-490E21.13 | UPPG | - | - | 0 |
| 179 | rs76935404 | 19 | 41419294 | 0.2556 | 21.1760 | T | upstream | CTC-490E21.13 | UPPG | - | - | 35 |
| 180 | rs10420231 | 19 | 41420030 | 0.1963 | 11.4360 | A | upstream | CTC-490E21.13 | UPPG | - | - | NA |
| 181 | rs8108939 | 19 | 41425149 | 0.2107 | 16.0161 | G | upstream | CYP2B7P | RI | - | - | 36 |
| 182 | rs4001943 | 19 | 41425900 | 0.2297 | 16.5281 | C | upstream | CYP2B7P | TUPG | - | - | 0 |
| 183 | rs10424844 | 19 | 41425959 | 0.1743 | 11.6110 | T | upstream | CYP2B7P | TUPG | - | - | 0 |
| 184 | rs12459565 | 19 | 41427539 | 0.2181 | 13.5166 | A | upstream | CYP2B7P | TUPG | - | - | 0 |
| 185 | rs28417358 | 19 | 41428105 | 0.2236 | 16.8030 | A | upstream | CYP2B7P | TUPG | - | - | 0 |
| 186 | rs11083589 | 19 | 41428416 | 0.2082 | 14.2242 | G | upstream | CYP2B7P | TUPG | - | - | NA |
| 187 | rs57274441 | 19 | 41431422 | 0.2286 | 16.7867 | G | intronNCT | CYP2B7P | TUPG | - | 1/8 | NA |
| 188 | rs3844443 | 19 | 41431935 | 0.2117 | 13.0324 | C | intronNCT | CYP2B7P | TUPG | - | 1/8 | 0 |
| 189 | rs12151139 | 19 | 41433543 | 0.2492 | 20.4082 | T | intronNCT | CYP2B7P | TUPG | - | 1/8 | 0 |
| 190 | rs4609955 | 19 | 41433613 | 0.2335 | 18.1488 | C | intronNCT | CYP2B7P | TUPG | - | 1/8 | 0 |
| 191 | rs3843043 | 19 | 41433931 | 0.2117 | 13.0324 | T | intronNCT | CYP2B7P | TUPG | - | 1/8 | NA |
| 192 | rs3844445 | 19 | 41434106 | 0.2179 | 14.6739 | G | intronNCT | CYP2B7P | TUPG | - | 1/8 | 1 |
| 193 | rs7248187 | 19 | 41437426 | 0.1768 | 10.2153 | C | intronNCT | CYP2B7P | TUPG | - | 1/8 | 0 |
| 194 | rs7247910 | 19 | 41437440 | 0.1798 | 10.5637 | G | intronNCT | CYP2B7P | TUPG | - | 1/8 | 4 |
| 195 | rs6508960 | 19 | 41437717 | 0.1646 | 8.8190 | A | intronNCT | CYP2B7P | TUPG | - | 1/8 | 0 |
| 196 | rs73038469 | 19 | 41442597 | 0.2493 | 16.1147 | A | intronNCT | CYP2B7P | TUPG | - | 3/8 | 0 |
| 197 | rs112819506 | 19 | 41444725 | -0.2502 | 4.7483 | G | intronNCT | CYP2B7P | TUPG | - | 3/8 | 4 |
| 198 | rs10407500 | 19 | 41446076 | -0.2832 | 4.3147 | C | intronNCT | CYP2B7P | TUPG | - | 4/8 | NA |
| 199 | rs56401945 | 19 | 41463690 | -0.2697 | 5.4766 | C | intergenic | - | - | - | - | 13 |
| 200 | rs16974790 | 19 | 41498946 | -0.2268 | 4.4593 | A | intron | CYP2B6 | PC | - | 1/8 | 1 |

**Supplememtary Table 3A. Top 200 Variants, Genome-wide Analysis of Total Nicotine Equivalents, Without CPD**

| Index | SNP | CHR | POS | beta | -log10p | Allele | Function | Symbol | Biotype | Exon | Intron | Count |
|---|---|---|---|---|---|---|---|---|---|---|---|---|
| 1 | rs7531583 | 1 | 1706160 | -0.3050 | 3.9311 | G | intron | NADK | PC | - | 1/11 | 36 |
| 2 | rs12045736 | 1 | 12654035 | -0.2560 | 3.4207 | T | intron | DHRS3 | PC | - | 1/5 | 23 |
| 3 | rs6686971 | 1 | 85152809 | -0.3933 | 4.1323 | T | intron | SSX2IP | PC | - | 1/13 | 36 |
| 4 | rs12037175 | 1 | 85170927 | -0.3782 | 3.8525 | A | regulatory | - | PFR | - | - | 0 |
| 5 | rs17631306 | 1 | 111072322 | -0.4280 | 3.4608 | A | intergenic | - | - | - | - | 36 |
| 6 | rs11118921 | 1 | 222160609 | 0.3359 | 3.8737 | A | downstream | RP11-400N13.2 | lincRNA | - | - | 36 |
| 7 | rs7552453 | 1 | 239856834 | -0.3168 | 4.7730 | T | intron | CHRM3 | PC | - | 3/4 | NA |
| 8 | rs2278644 | 1 | 239867792 | -0.3075 | 4.5491 | C | intron | CHRM3 | PC | - | 3/4 | 0 |
| 9 | rs10802794 | 1 | 239870621 | -0.3122 | 4.6986 | T | intron | CHRM3 | PC | - | 3/4 | 0 |
| 10 | rs10802795 | 1 | 239870775 | -0.3038 | 4.5210 | C | intron | CHRM3 | PC | - | 3/4 | 18 |
| 11 | rs6684622 | 1 | 239877537 | -0.3099 | 4.7385 | C | intron | CHRM3 | PC | - | 3/4 | 2 |
| 12 | rs6663632 | 1 | 239877721 | -0.3037 | 4.5529 | A | intron | CHRM3 | PC | - | 3/4 | 6 |
| 13 | rs1431719 | 1 | 239881203 | -0.3072 | 4.5480 | G | intron | CHRM3 | PC | - | 3/4 | 0 |
| 14 | rs11583349 | 1 | 239901107 | -0.3174 | 4.8238 | T | intron | CHRM3 | PC | - | 3/4 | 0 |
| 15 | rs7513757 | 1 | 239901317 | -0.3089 | 4.6397 | A | intron | CHRM3 | PC | - | 3/4 | NA |
| 16 | rs1416789 | 1 | 239901645 | -0.3268 | 5.1701 | G | intron | CHRM3 | PC | - | 3/4 | 1 |
| 17 | rs10925964 | 1 | 239902514 | -0.2795 | 3.7828 | A | intron | CHRM3 | PC | - | 3/4 | 0 |
| 18 | rs10802801 | 1 | 239902841 | -0.2756 | 3.6606 | A | intron | CHRM3 | PC | - | 3/4 | 0 |
| 19 | chr1:239904987:I | 1 | 239904987 | -0.2614 | 3.5144 | - | not defined | - | - | - | - | NA |
| 20 | rs12060884 | 1 | 239905830 | -0.2760 | 3.8894 | G | intron | CHRM3 | PC | - | 3/4 | 0 |
| 21 | rs1544170 | 1 | 239908236 | -0.3405 | 5.5369 | A | intron | CHRM3 | PC | - | 3/4 | 0 |
| 22 | rs11585281 | 1 | 239909651 | -0.2663 | 3.5321 | T | intron | CHRM3 | PC | - | 3/4 | 0 |
| 23 | rs7537514 | 1 | 239910572 | -0.2630 | 3.4525 | G | intron | CHRM3 | PC | - | 3/4 | 0 |
| 24 | rs934344 | 1 | 239910999 | -0.2587 | 3.4410 | A | intron | CHRM3 | PC | - | 3/4 | 0 |
| 25 | rs12126146 | 1 | 239917787 | 0.5383 | 4.5355 | G | intron | CHRM3 | PC | - | 3/4 | 36 |
| 26 | rs10167265 | 2 | 42087252 | 0.2843 | 3.4518 | A | upstream | Y_RNA | miscRNA | - | - | 36 |
| 27 | rs4672114 | 2 | 56614299 | 0.2989 | 3.7392 | C | downstream | CCDC85A | PC | - | - | 36 |
| 28 | rs11680204 | 2 | 168574906 | 0.4079 | 3.6089 | T | upstream | CTAGE14P | PPG | - | - | 36 |
| 29 | rs72973408 | 2 | 236047364 | -0.4122 | 3.7914 | A | regulatory | - | CTCF | - | - | 36 |
| 30 | rs1472476 | 3 | 7052542 | 0.3185 | 3.4308 | A | intron | GRM7 | PC | - | 1/9 | 36 |
| 31 | rs1391950 | 3 | 7058417 | 0.2594 | 3.4230 | A | intron | GRM7 | PC | - | 1/9 | 36 |
| 32 | rs2620558 | 3 | 22111050 | -0.4592 | 4.8685 | G | intronNCT | ZNF385D | PT | - | 1/1 | 36 |
| 33 | rs16899 | 3 | 87811945 | -0.3147 | 3.4685 | A | intronNCT | RP11-451B8.1 | lincRNA | - | 1/1 | 36 |
| 34 | rs2125109 | 3 | 182277971 | -0.3147 | 3.9775 | T | intergenic | - | - | - | - | 36 |
| 35 | rs17442778 | 4 | 41064147 | 0.4478 | 4.9059 | A | intron | APBB2 | PC | - | 4/17 | 36 |
| 36 | rs13131251 | 4 | 62942299 | -0.3436 | 5.0877 | C | downstream | LPHN3 | PC | - | - | 28 |
| 37 | rs10026213 | 4 | 62942911 | -0.3405 | 4.8719 | C | downstream | LPHN3 | PC | - | - | 6 |
| 38 | rs12715707 | 4 | 62948759 | -0.3044 | 3.6229 | T | intronNCT | RP11-84A1.3 | antisense | - | 6/7 | 6 |
| 39 | rs9999827 | 4 | 62954273 | -0.3021 | 4.2789 | G | intronNCT | RP11-84A1.3 | antisense | - | 6/7 | 24 |
| 40 | rs34240473 | 4 | 128635022 | 0.3004 | 3.8749 | T | intron | INTU | PC | - | 14/15 | 36 |
| 41 | rs1216365 | 4 | 129569808 | -0.2903 | 4.0722 | T | regulatory | - | enhancer | - | - | 36 |
| 42 | rs9685999 | 4 | 156963413 | 0.2814 | 3.5841 | A | intergenic | - | - | - | - | 36 |
| 43 | rs10520270 | 4 | 175046985 | -0.4875 | 4.2092 | A | intronNCT | RP11-148L24.1 | lincRNA | - | 1/4 | 0 |
| 44 | rs1992019 | 4 | 175051486 | -0.5110 | 4.4984 | A | downstream | RP11-248N22.1 | lincRNA | - | - | 36 |
| 45 | rs425620 | 5 | 9413362 | -0.3708 | 4.2367 | A | intron | SEMA5A | PC | - | 2/22 | 36 |
| 46 | rs2081922 | 5 | 24457943 | -0.4410 | 3.8994 | G | intergenic | - | - | - | - | 36 |
| 47 | rs62345637 | 5 | 26216260 | 0.3076 | 4.3693 | A | intergenic | - | - | - | - | 36 |
| 48 | rs6450194 | 5 | 53702552 | -0.2976 | 4.1551 | A | intronNCT | LINC01033 | lincRNA | - | 4/5 | 36 |
| 49 | rs6887887 | 5 | 142535913 | 0.5209 | 4.0966 | A | intron | ARHGAP26 | PC | - | 20/22 | 36 |
| 50 | rs78817974 | 5 | 160881054 | -0.3555 | 4.0436 | T | intron | GABRB2 | PC | - | 5/10 | NA |
| 51 | rs6869521 | 5 | 160889272 | -0.3501 | 3.6628 | C | intron | GABRB2 | PC | - | 4/10 | 0 |
| 52 | rs62381570 | 5 | 160892153 | -0.3328 | 3.5751 | C | intron | GABRB2 | PC | - | 4/10 | 36 |
| 53 | rs4713925 | 6 | 11798414 | 0.3118 | 4.7262 | T | intron | ADTRP | PC | - | 2/5 | 36 |
| 54 | rs196701 | 6 | 80147187 | -0.3887 | 3.6808 | A | upstream | DBIP1 | PPG | - | - | 36 |
| 55 | rs9400512 | 6 | 112211802 | 0.2743 | 3.4607 | T | intergenic | - | - | - | - | 36 |
| 56 | rs728017 | 6 | 124292594 | 0.3170 | 3.6985 | G | intron | NKAIN2 | PC | - | 1/3 | 36 |
| 57 | rs1057793 | 6 | 148835416 | 0.3115 | 3.5635 | T | intron | SASH1 | PC | - | 8/19 | 36 |
| 58 | rs17729786 | 6 | 149304906 | -0.4915 | 3.4155 | T | intron | UST | PC | - | 5/7 | 36 |
| 59 | rs7798735 | 7 | 10146577 | 0.3279 | 3.5347 | A | intergenic | - | - | - | - | 36 |
| 60 | rs17763518 | 7 | 14230968 | -0.4417 | 3.6978 | C | intron | DGKB | PC | - | 22/24 | 36 |
| 61 | rs17171441 | 7 | 38722499 | 0.3496 | 3.5588 | C | downstream | FAM183B | PC | - | - | 36 |
| 62 | rs968908 | 7 | 89341720 | 0.2779 | 3.7748 | C | intergenic | - | - | - | - | 36 |
| 63 | rs74782537 | 7 | 151531336 | 0.4084 | 3.6391 | C | intron | PRKAG2 | PC | - | 1/15 | 36 |
| 64 | rs6558708 | 8 | 2926193 | 0.4177 | 3.5653 | A | intron | CSMD1 | PC | - | 36/55 | 36 |
| 65 | rs68112061 | 8 | 2931097 | 0.4824 | 3.4320 | C | intron | CSMD1 | PC | - | 36/55 | 0 |
| 66 | rs11774005 | 8 | 2966358 | 0.4847 | 3.8118 | A | intron | CSMD1 | PC | - | 31/55 | 36 |
| 67 | rs17390567 | 8 | 2968364 | 0.4987 | 3.5075 | C | intron | CSMD1 | PC | - | 30/55 | 0 |

| | | | | | | | | | | | | |
|---|---|---|---|---|---|---|---|---|---|---|---|---|
| 68 | rs9650503 | 8 | 3066067 | 0.2574 | 3.5014 | G | intron | CSMD1 | PC | - | 18/55 | 27 |
| 69 | rs7835399 | 8 | 3115052 | 0.3740 | 4.8336 | A | intron | CSMD1 | PC | - | 14/55 | 36 |
| 70 | rs13248596 | 8 | 3120483 | 0.3633 | 4.3345 | G | intron | CSMD1 | PC | - | 14/55 | 0 |
| 71 | rs2897414 | 8 | 3122111 | 0.2886 | 3.6308 | G | intron | CSMD1 | PC | - | 14/55 | 17 |
| 72 | rs13254027 | 8 | 3125924 | 0.3325 | 3.7247 | A | intron | CSMD1 | PC | - | 14/55 | 0 |
| 73 | rs12541801 | 8 | 39611992 | -0.2807 | 3.9202 | A | intron | ADAM2 | PC | - | 16/20 | 36 |
| 74 | rs12548821 | 8 | 39615296 | -0.2707 | 3.6538 | A | intron | ADAM2 | PC | - | 15/20 | 0 |
| 75 | rs4873713 | 8 | 54001049 | 0.2780 | 3.7236 | C | intergenic | - | - | - | - | 0 |
| 76 | rs2376432 | 8 | 54003760 | 0.2792 | 3.7340 | A | intergenic | - | - | - | - | 0 |
| 77 | rs4873722 | 8 | 54007044 | 0.2779 | 3.7210 | A | intergenic | - | - | - | - | 0 |
| 78 | rs4873729 | 8 | 54029036 | 0.2723 | 3.5399 | A | intergenic | - | - | - | - | 0 |
| 79 | rs2553916 | 8 | 54038872 | 0.2782 | 3.6382 | C | intergenic | - | - | - | - | 27 |
| 80 | rs2717637 | 8 | 54039378 | 0.2842 | 3.8374 | A | intergenic | - | - | - | - | 7 |
| 81 | rs34439026 | 8 | 74328861 | 0.4096 | 3.6058 | T | upstream | STAU2-AS1 | lincRNA | - | - | NA |
| 82 | rs34643738 | 8 | 112466724 | 0.5892 | 3.8403 | C | intronNCT | RP11-1101K5.1 | lincRNA | - | 2/4 | 36 |
| 83 | rs2125553 | 8 | 113900367 | -0.4389 | 4.5069 | C | intron | CSMD3 | PC | - | 10/70 | 36 |
| 84 | rs17715679 | 8 | 116565162 | 0.4204 | 3.6188 | T | intron | TRPS1 | PC | - | 4/5 | 36 |
| 85 | rs56282194 | 8 | 133574214 | -0.2882 | 3.4208 | A | upstream | HPYR1 | lincRNA | - | - | 36 |
| 86 | rs13439493 | 8 | 135108404 | 0.3995 | 3.4595 | T | intergenic | - | - | - | - | 36 |
| 87 | rs2960109 | 8 | 138444604 | -0.2779 | 3.4353 | C | intergenic | - | - | - | - | 17 |
| 88 | rs28406373 | 9 | 7618765 | -0.3041 | 3.4636 | A | intergenic | - | - | - | - | 36 |
| 89 | rs2417654 | 9 | 108643176 | -0.3493 | 3.4297 | T | intergenic | - | - | - | - | 36 |
| 90 | rs10982256 | 9 | 117260834 | 0.2743 | 3.4679 | T | intron | DFNB31 | PC | - | 1/11 | 36 |
| 91 | rs10985450 | 9 | 124685381 | 0.3129 | 3.6075 | C | intron | TTLL11 | PC | - | 6/8 | 36 |
| 92 | rs11013700 | 10 | 18626566 | 0.3460 | 4.4230 | T | intron | CACNB2 | PC | - | 2/13 | 36 |
| 93 | rs977754 | 10 | 44817419 | -0.4296 | 4.3083 | T | intron | CXCL12 | PC | - | 3/3 | NA |
| 94 | rs1720367 | 10 | 50908644 | -0.3254 | 3.7036 | A | intron | C10orf53 | PC | - | 2/2 | NA |
| 95 | rs34016608 | 10 | 78911244 | 0.3407 | 3.6102 | C | intron | KCNMA1 | PC | - | 5/26 | 36 |
| 96 | rs12241006 | 10 | 79030744 | -0.4174 | 4.5664 | A | intron | KCNMA1 | PC | - | 2/26 | 6 |
| 97 | rs181836 | 10 | 79124420 | -0.3055 | 3.4586 | G | intron | KCNMA1 | PC | - | 2/26 | 17 |
| 98 | rs7919945 | 10 | 87724145 | -0.4792 | 4.0105 | A | intron | GRID1 | PC | - | 4/15 | 36 |
| 99 | rs12773375 | 10 | 108134101 | -0.5928 | 3.8468 | A | intergenic | - | - | - | - | 18 |
| 100 | rs58634906 | 10 | 118986090 | -0.4506 | 4.8131 | A | intergenic | - | - | - | - | 0 |
| 101 | rs7393602 | 10 | 118987781 | -0.4361 | 4.6525 | C | intergenic | - | - | - | - | 6 |
| 102 | rs12414919 | 10 | 118991768 | -0.5127 | 4.4915 | A | intergenic | - | - | - | - | 36 |
| 103 | rs2283135 | 10 | 118999932 | -0.4134 | 3.7184 | G | upstream | SLC18A2 | PC | - | - | 0 |
| 104 | rs12412905 | 10 | 119000560 | -0.4156 | 3.7509 | T | upstream | SLC18A2 | PC | - | - | NA |
| 105 | rs363387 | 10 | 119003564 | -0.4077 | 3.5896 | A | synonymous | SLC18A2 | PC | 3/16 | - | 0 |
| 106 | rs2283136 | 10 | 119006945 | -0.3996 | 3.5839 | G | intron | SLC18A2 | PC | - | 3/15 | 0 |
| 107 | rs2532805 | 10 | 119009966 | 0.3744 | 3.4547 | A | intron | SLC18A2 | PC | - | 3/15 | 0 |
| 108 | rs1396860 | 11 | 6283452 | 0.4029 | 3.6911 | C | intron | CCKBR | PC | - | 1/4 | 0 |
| 109 | rs3793993 | 11 | 6285553 | 0.3903 | 3.5271 | C | intron | CCKBR | PC | - | 1/4 | 0 |
| 110 | rs1112716 | 11 | 6286156 | 0.4037 | 3.4293 | A | intron | CCKBR | PC | - | 1/4 | 6 |
| 111 | rs2929184 | 11 | 6289118 | 0.3823 | 3.5190 | G | intron | CCKBR | PC | - | 1/4 | 36 |
| 112 | rs11026012 | 11 | 21330530 | -0.3144 | 3.4540 | G | intron | NELL1 | PC | - | 15/20 | 6 |
| 113 | rs1535717 | 11 | 34501166 | -0.3702 | 3.4805 | A | 3'UTR | ELF5 | PC | 7/7 | - | 36 |
| 114 | rs11036034 | 11 | 40735958 | -0.2674 | 3.6580 | A | intron | LRRC4C | PC | - | 2/6 | 36 |
| 115 | rs28568946 | 11 | 70419429 | -0.3359 | 3.6677 | C | intron | SHANK2 | PC | - | 7/14 | 36 |
| 116 | rs10765595 | 11 | 88263649 | -0.4581 | 4.1653 | G | intron | GRM5 | PC | - | 7/7 | 36 |
| 117 | rs972938 | 11 | 88594590 | -0.3477 | 3.4228 | C | intron | GRM5 | PC | - | 1/7 | 0 |
| 118 | rs10831536 | 11 | 88597438 | -0.3665 | 3.7570 | A | intron | GRM5 | PC | - | 1/7 | 6 |
| 119 | rs10831537 | 11 | 88597440 | -0.3916 | 4.1225 | C | intron | GRM5 | PC | - | 1/7 | 6 |
| 120 | rs10765745 | 11 | 95252080 | 0.3093 | 4.0350 | A | intergenic | - | - | - | - | 36 |
| 121 | rs12805779 | 11 | 116523063 | 0.5126 | 3.7440 | T | intronNCT | AP000770.1 | lincRNA | - | 1/2 | 36 |
| 122 | rs1275582 | 12 | 76255406 | -0.3140 | 3.8429 | T | intronNCT | RP11-114H23.1 | lincRNA | - | 1/4 | 36 |
| 123 | rs4760385 | 12 | 92665526 | -0.2757 | 3.5602 | G | intergenic | - | - | - | - | 36 |
| 124 | rs4767528 | 12 | 117719054 | -0.3462 | 3.9129 | A | intron | NOS1 | PC | - | 7/28 | 36 |
| 125 | rs9603543 | 13 | 40036131 | 0.2662 | 3.5945 | C | intron | LHFP | PC | - | 2/3 | 36 |
| 126 | rs61035869 | 13 | 101979673 | -0.3950 | 3.9964 | A | intron | NALCN | PC | - | 7/43 | 36 |
| 127 | rs7155706 | 14 | 58447752 | -0.2917 | 3.6096 | A | intronNCT | SLC35F4 | PT | - | 1/1 | 36 |
| 128 | rs937051 | 15 | 29405523 | -0.3392 | 3.5672 | G | intron | APBA2 | PC | - | 11/12 | 17 |
| 129 | rs586642 | 15 | 71432007 | -0.5550 | 3.5514 | A | upstream | THSD4 | PC | - | - | 36 |
| 130 | rs951985 | 15 | 78720923 | 0.5046 | 3.6518 | G | regulatory | - | PFR | - | - | NA |
| 131 | chr15:78801393:I | 15 | 78801393 | 0.5561 | 4.1937 | - | not defined | - | - | - | - | NA |
| 132 | rs11852372 | 15 | 78801394 | 0.5393 | 4.0683 | C | intron | HYKK | PC | - | 1/3 | NA |
| 133 | rs8034191 | 15 | 78806023 | 0.5435 | 4.3326 | C | intron | HYKK | PC | - | 2/3 | 17 |
| 134 | rs8031948 | 15 | 78816057 | 0.5078 | 3.8824 | T | intron | HYKK | PC | - | 3/4 | 0 |
| 135 | rs58365910 | 15 | 78849034 | 0.5416 | 4.5894 | C | intergenic | - | - | - | - | 0 |
| 136 | rs72740955 | 15 | 78849779 | 0.5718 | 4.2388 | T | intergenic | - | - | - | - | 0 |

| | | | | | | | | | | | | |
|---|---|---|---|---|---|---|---|---|---|---|---|---|
| 137 | rs2036527 | 15 | 78851615 | 0.5686 | 4.8710 | A | intergenic | - | - | - | - | 7 |
| 138 | rs55781567 | 15 | 78857986 | 0.4499 | 3.4403 | G | 5'UTR | CHRNA5 | PC | 1/6 | - | 0 |
| 139 | rs11633958 | 15 | 78862064 | 0.5692 | 3.7233 | A | intron | CHRNA5 | PC | - | 1/5 | 0 |
| 140 | rs7172118 | 15 | 78862453 | 0.5595 | 3.9349 | A | intron | CHRNA5 | PC | - | 1/5 | 0 |
| 141 | rs17486195 | 15 | 78865197 | 0.5323 | 3.6832 | G | intron | CHRNA5 | PC | - | 1/5 | 0 |
| 142 | rs140330585 | 15 | 78866445 | 0.5166 | 3.5079 | A | intron | CHRNA5 | PC | - | 1/5 | 0 |
| 143 | rs17486278 | 15 | 78867482 | 0.3950 | 4.3513 | C | intron | CHRNA5 | PC | - | 1/5 | 6 |
| 144 | rs72740964 | 15 | 78868636 | 0.5665 | 3.7651 | A | intron | CHRNA5 | PC | - | 1/5 | 0 |
| 145 | rs7180002 | 15 | 78873993 | 0.5571 | 3.9229 | T | intron | CHRNA5 | PC | - | 2/5 | 0 |
| 146 | rs56390833 | 15 | 78877381 | 0.5596 | 3.9495 | A | intron | CHRNA5 | PC | - | 2/5 | 0 |
| 147 | rs951266 | 15 | 78878541 | 0.5630 | 3.9847 | A | intron | CHRNA5 | PC | - | 2/5 | 0 |
| 148 | rs16969968 | 15 | 78882925 | 0.5791 | 3.8265 | A | missense | CHRNA5 | PC | 5/6 | - | 0 |
| 149 | rs8192482 | 15 | 78886198 | 0.5840 | 3.9057 | T | 3'UTR | CHRNA5 | PC | 6/6 | - | 0 |
| 150 | rs4887067 | 15 | 78886947 | 0.5840 | 3.9061 | A | 3'UTR | CHRNA5 | PC | 6/6 | - | 0 |
| 151 | rs1051730 | 15 | 78894339 | 0.5724 | 4.2294 | A | synonymous | CHRNA3 | PC | 5/6 | - | 8 |
| 152 | rs12914385 | 15 | 78898723 | 0.3960 | 4.1171 | A | intron | CHRNA3 | PC | - | 4/5 | 17 |
| 153 | rs55676755 | 15 | 78898932 | 0.5862 | 4.7666 | G | intron | CHRNA3 | PC | - | 4/5 | 36 |
| 154 | rs56077333 | 15 | 78899003 | 0.3944 | 4.0496 | A | intron | CHRNA3 | PC | - | 4/5 | 0 |
| 155 | rs147144681 | 15 | 78900908 | 0.5475 | 4.0104 | T | intron | CHRNA3 | PC | - | 4/5 | NA |
| 156 | rs114205691 | 15 | 78901113 | 0.3952 | 4.1874 | A | intron | CHRNA3 | PC | - | 4/5 | 0 |
| 157 | rs146009840 | 15 | 78906177 | 0.5587 | 3.8847 | T | intron | CHRNA3 | PC | - | 4/5 | 0 |
| 158 | rs4243084 | 15 | 78911672 | 0.3486 | 3.4520 | C | downstream | CHRNB4 | PC | - | - | 6 |
| 159 | rs55958997 | 15 | 78915872 | 0.3806 | 3.5458 | A | downstream | CHRNB4 | PC | - | - | NA |
| 160 | rs17487223 | 15 | 78923987 | 0.5216 | 3.5639 | T | intron | CHRNB4 | PC | - | 2/5 | 6 |
| 161 | rs2679090 | 15 | 87954255 | 0.2798 | 3.4428 | T | intergenic | - | - | - | - | 36 |
| 162 | rs12447354 | 16 | 2946055 | -0.2830 | 3.8454 | A | intron | FLYWCH2 | PC | - | 2/3 | 27 |
| 163 | rs9898328 | 17 | 40032356 | 0.4522 | 5.0642 | A | intron | ACLY | PC | - | 22/28 | 36 |
| 164 | rs891765 | 17 | 65474156 | 0.3175 | 4.1030 | G | intron | PITPNC1 | PC | - | 1/9 | 36 |
| 165 | rs2949923 | 17 | 65475228 | 0.2955 | 4.2622 | G | intron | PITPNC1 | PC | - | 1/9 | 19 |
| 166 | rs1004413 | 18 | 3042297 | -0.2684 | 3.4788 | G | regulatory | - | PFR | - | - | 23 |
| 167 | rs12185370 | 18 | 4582373 | 0.3313 | 3.4158 | A | intergenic | - | - | - | - | 36 |
| 168 | rs11081290 | 18 | 6979570 | -0.3818 | 4.0789 | A | intron | LAMA1 | PC | - | 42/62 | 36 |
| 169 | rs55787463 | 18 | 76383097 | 0.3292 | 3.4839 | C | intergenic | - | - | - | - | 36 |
| 170 | rs11878604 | 19 | 41333284 | -0.3824 | 4.5339 | C | upstream | CYP2F2P | UnitaryPG | - | - | NA |
| 171 | rs12459249 | 19 | 41339896 | 0.3109 | 4.3157 | C | intronNCT | - | lincRNA | - | 1/1 | 6 |
| 172 | rs10853742 | 19 | 41340573 | 0.3025 | 4.1026 | A | intronNCT | - | lincRNA | - | 1/1 | 0 |
| 173 | rs11667314 | 19 | 41340983 | 0.3049 | 4.1584 | C | intronNCT | - | lincRNA | - | 1/1 | 0 |
| 174 | rs12461964 | 19 | 41341229 | 0.2976 | 3.9333 | G | intronNCT | - | lincRNA | - | 1/1 | 0 |
| 175 | rs35755165 | 19 | 41345989 | 0.2940 | 3.6471 | C | downstream | CYP2A6 | PC | - | - | 0 |
| 176 | rs2316205 | 19 | 41346768 | 0.2845 | 3.4839 | C | downstream | CYP2A6 | PC | - | - | 0 |
| 177 | rs60446182 | 19 | 41347998 | 0.3067 | 3.9081 | G | downstream | CYP2A6 | PC | - | - | 0 |
| 178 | rs56113850 | 19 | 41353107 | 0.4305 | 6.5938 | C | intron | CYP2A6 | PC | - | 4/8 | 36 |
| 179 | rs150298687 | 19 | 41357344 | 0.3086 | 3.4200 | C | upstream | CYP2A6 | PC | - | - | 6 |
| 180 | rs57837628 | 19 | 41357910 | 0.3880 | 5.1070 | G | upstream | CYP2A6 | PC | - | - | 0 |
| 181 | rs113029345 | 19 | 41370176 | 0.3870 | 4.7138 | C | intronNMD | - | NMD | - | 1/3 | 0 |
| 182 | rs12461383 | 19 | 41370338 | 0.3953 | 5.0713 | G | intronNMD | - | NMD | - | 1/3 | 0 |
| 183 | rs7247903 | 19 | 41372475 | -0.4049 | 3.4858 | G | intronNMD | - | NMD | - | 1/3 | NA |
| 184 | rs56097499 | 19 | 41381334 | 0.4184 | 4.4893 | A | downstream | CYP2A7 | PC | - | - | 9 |
| 185 | rs2261144 | 19 | 41383153 | 0.3485 | 4.1328 | G | missense | CYP2A7 | PC | 6/8 | - | 0 |
| 186 | rs3822479 | 19 | 41383989 | 0.3617 | 4.0304 | T | intron | CYP2A7 | PC | - | 4/7 | 0 |
| 187 | rs3815705 | 19 | 41386282 | 0.3290 | 4.0203 | A | intron | CYP2A7 | PC | - | 2/7 | 18 |
| 188 | rs72480748 | 19 | 41414481 | 0.3306 | 4.0085 | A | NCTexon | CTC-490E21.13 | UPPG | 5/5 | - | 0 |
| 189 | rs73038469 | 19 | 41442597 | 0.3253 | 3.4780 | A | intronNCT | CYP2B7P | TUPG | - | 3/8 | 36 |
| 190 | rs1002929 | 20 | 5209217 | 0.3105 | 4.6974 | A | intergenic | - | - | - | - | 36 |
| 191 | rs6074121 | 20 | 10290703 | -0.2743 | 3.5040 | A | downstream | SNAP25 | PC | - | - | 1 |
| 192 | rs7268664 | 20 | 10295257 | -0.2890 | 3.8876 | T | intronNCT | SNAP25-AS1 | antisense | - | 1/3 | 36 |
| 193 | rs13433103 | 20 | 11251165 | 0.2980 | 3.6307 | G | upstream | RP4-734C18.1 | lincRNA | - | - | 36 |
| 194 | rs871913 | 20 | 16144345 | 0.4365 | 3.4784 | A | intergenic | - | - | - | - | 36 |
| 195 | rs6068022 | 20 | 50565828 | 0.2590 | 3.5183 | G | regulatory | - | enhancer | - | - | 36 |
| 196 | rs12481177 | 20 | 57835389 | 0.3912 | 4.2816 | A | downstream | ZNF831 | PC | - | - | 36 |
| 197 | rs2833270 | 21 | 32487354 | -0.4135 | 3.6820 | G | downstream | TIAM1 | PC | - | - | 30 |
| 198 | rs736898 | 22 | 22711786 | 0.3373 | 3.6245 | T | downstream | IGLV5-48 | IGVG | - | - | 36 |
| 199 | rs132952 | 22 | 38549373 | -0.3181 | 3.8154 | C | intron | PLA2G6 | PC | - | 2/16 | 36 |
| 200 | rs4822253 | 22 | 43427776 | 0.4068 | 3.5485 | A | intergenic | - | - | - | - | 36 |

**Supplemenmtary Table 3B. Top 200 Variants, Genome-wide Analysis of Total Nicotine Equivalents, With CPD**

| Index | SNP | CHR | POS | beta | -log10p | Allele | Function | Symbol | Biotype | Exon | Intron | Count |
|---|---|---|---|---|---|---|---|---|---|---|---|---|
| 1 | rs1023252 | 1 | 11899033 | -0.1164 | 3.3180 | A | intron | CLCN6 | PC | - | 21/21 | 19 |
| 2 | rs4652959 | 1 | 38015372 | 0.2850 | 3.5262 | G | intron | SNIP1 | PC | - | 2/3 | 27 |
| 3 | rs857093 | 1 | 57232805 | 0.1774 | 3.8749 | C | intron | FYB2 | PC | - | 5/19 | 25 |
| 4 | rs2811883 | 1 | 59230228 | 0.1249 | 3.6097 | A | intronNCT | AL136985.3 | lncRNA | - | 1/3 | 30 |
| 5 | rs11208512 | 1 | 65170405 | 0.0126 | 3.3698 | A | intergenic | - | - | - | - | 20 |
| 6 | rs10493405 | 1 | 66920877 | 0.1450 | 3.6926 | A | intergenic | - | - | - | - | 30 |
| 7 | rs17631306 | 1 | 111072322 | 0.1840 | 3.6323 | A | intergenic | - | - | - | - | 24 |
| 8 | rs6674761 | 1 | 146942884 | -0.0666 | 3.3182 | T | intronNCT | LINC00624 | lncRNA | - | 3/5 | 24 |
| 9 | rs3811454 | 1 | 154164139 | 0.1508 | 3.6389 | C | intron | TPM3 | PC | - | 1/8 | 30 |
| 10 | rs79609663 | 1 | 201753696 | 0.1208 | 3.8019 | A | intron | NAV1 | PC | - | 5/26 | 24 |
| 11 | rs11118921 | 1 | 222160609 | -0.2912 | 4.5726 | A | downstream | RP11-400N13.2 | lincRNA | - | - | 24 |
| 12 | rs7552453 | 1 | 239856834 | 0.2494 | 3.7409 | T | intron | CHRM3 | PC | - | 3/4 | NA |
| 13 | rs2278644 | 1 | 239867792 | 0.1480 | 3.7442 | C | intron | CHRM3 | PC | - | 3/4 | 0 |
| 14 | rs10802794 | 1 | 239870621 | 0.1945 | 4.1389 | T | intron | CHRM3 | PC | - | 3/4 | 0 |
| 15 | rs10802795 | 1 | 239870775 | -0.2676 | 4.3075 | C | intron | CHRM3 | PC | - | 3/4 | 24 |
| 16 | rs6684622 | 1 | 239877537 | 0.1934 | 4.1744 | C | intron | CHRM3 | PC | - | 3/4 | 0 |
| 17 | rs6663632 | 1 | 239877721 | 0.1777 | 4.0098 | A | intron | CHRM3 | PC | - | 3/4 | 0 |
| 18 | rs1431719 | 1 | 239881203 | 0.1622 | 3.6848 | G | intron | CHRM3 | PC | - | 3/4 | 1 |
| 19 | rs11583349 | 1 | 239901107 | 0.1339 | 3.8206 | T | intron | CHRM3 | PC | - | 3/4 | 0 |
| 20 | rs7513757 | 1 | 239901317 | -0.2194 | 3.4494 | A | intron | CHRM3 | PC | - | 3/4 | NA |
| 21 | rs1416789 | 1 | 239901645 | 0.2292 | 4.4998 | G | intron | CHRM3 | PC | - | 3/4 | 0 |
| 22 | rs1544170 | 1 | 239908236 | 0.1672 | 4.6227 | A | intron | CHRM3 | PC | - | 3/4 | 0 |
| 23 | rs1915884 | 1 | 241383781 | 0.1145 | 3.5571 | A | intron | RGS7 | PC | - | 1/14 | 24 |
| 24 | rs74783457 | 1 | 241623551 | 0.1029 | 3.4493 | C | intronNCT | AL359764.2 | lncRNA | - | 3/3 | 30 |
| 25 | rs6708742 | 2 | 1788251 | 0.0888 | 3.4825 | C | downstream | MYT1L | PC | - | - | 30 |
| 26 | rs11903584 | 2 | 75672583 | 0.3239 | 5.2095 | G | intergenic | - | - | - | - | 30 |
| 27 | rs7570009 | 2 | 98403683 | 0.1113 | 3.6486 | T | intron | TMEM131 | PC | - | 31/40 | 30 |
| 28 | rs1604689 | 2 | 214303523 | -0.2343 | 3.4554 | G | intron | SPAG16 | PC | - | 9/15 | 19 |
| 29 | rs6806144 | 3 | 1323078 | 0.1149 | 3.7410 | A | intron | CNTN6 | PC | - | 5/22 | 24 |
| 30 | rs2620558 | 3 | 22111050 | 0.1787 | 3.9345 | G | intronNCT | ZNF385D | PT | - | 1/1 | 24 |
| 31 | rs73824112 | 3 | 31383686 | 0.2633 | 4.1181 | A | intergenic | - | - | - | - | 30 |
| 32 | rs17442778 | 4 | 41064147 | 0.2743 | 3.8339 | A | intron | APBB2 | PC | - | 4/17 | 31 |
| 33 | rs1522095 | 4 | 59222703 | 0.1416 | 3.6655 | G | intergenic | - | - | - | - | 24 |
| 34 | rs13131251 | 4 | 62942299 | 0.2623 | 4.2147 | C | downstream | LPHN3 | PC | - | - | 12 |
| 35 | rs10026213 | 4 | 62942911 | 0.2708 | 4.1275 | C | downstream | LPHN3 | PC | - | - | 3 |
| 36 | rs9999827 | 4 | 62954273 | 0.1752 | 3.8782 | G | intronNCT | RP11-84A1.3 | antisense | - | 6/7 | 24 |
| 37 | rs17627471 | 4 | 112906965 | 0.1044 | 3.6554 | C | intronNCT | AC004704.1 | lncRNA | - | 1/2 | 24 |
| 38 | rs1216365 | 4 | 129569808 | -0.0484 | 3.3536 | T | regulatory | - | enhancer | - | - | 24 |
| 39 | rs6852267 | 4 | 140692082 | -0.0680 | 3.4049 | G | intron | MAML3 | PC | - | 1/3 | 24 |
| 40 | rs9685999 | 4 | 156963413 | 0.1532 | 3.6126 | A | intergenic | - | - | - | - | 24 |
| 41 | rs72683600 | 4 | 156966121 | -0.0805 | 3.4357 | G | intergenic | - | - | - | - | 0 |
| 42 | rs4691129 | 4 | 165609979 | 0.1457 | 3.6375 | T | intergenic | - | - | - | - | 24 |
| 43 | rs1992019 | 4 | 175051486 | 0.0130 | 3.3267 | A | downstream | RP11-248N22.1 | lincRNA | - | - | 26 |
| 44 | rs28892343 | 4 | 180311659 | -0.1195 | 3.6112 | T | intronNCT | AC020551.1 | lncRNA | - | 1/2 | 24 |
| 45 | rs6347 | 5 | 1411412 | -0.1060 | 3.3446 | C | synonymous | SLC6A3 | PC | 9/15 | - | 24 |
| 46 | rs6555217 | 5 | 3685242 | 0.1547 | 3.7193 | A | intergenic | - | - | - | - | 31 |
| 47 | rs60178658 | 5 | 11694217 | 0.1044 | 3.6362 | C | intron | CTNND2 | PC | - | 2/21 | NA |
| 48 | rs73746522 | 5 | 24448102 | 0.1007 | 3.3519 | T | downstream | Metazoa_SRP | miscRNA | - | - | 24 |
| 49 | rs2081922 | 5 | 24457943 | -0.1194 | 3.3563 | G | intergenic | - | - | - | - | 6 |
| 50 | rs6450194 | 5 | 53702552 | -0.3214 | 4.9504 | A | intronNCT | LINC01033 | lincRNA | - | 4/5 | 24 |
| 51 | rs10939908 | 5 | 60879560 | 0.1403 | 3.4367 | C | intergenic | - | - | - | - | 24 |
| 52 | rs10070074 | 5 | 90022407 | 0.0884 | 3.5732 | C | intron | ADGRV1 | PC | - | 48/89 | 22 |
| 53 | rs2460162 | 5 | 90047491 | 0.1006 | 3.4314 | T | intron | ADGRV1 | PC | - | 53/89 | 0 |
| 54 | chr5:90049162:I | 5 | 90049162 | 0.1916 | 4.0799 | NA | NA | NA | NA | NA | NA | NA |
| 55 | rs10062026 | 5 | 90052289 | 0.2025 | 4.2589 | A | missense | ADGRV1 | PC | 56/90 | - | 0 |
| 56 | rs73781039 | 5 | 101065708 | 0.0896 | 3.4666 | G | intergenic | - | - | - | - | NA |
| 57 | rs35715303 | 5 | 114730406 | 0.2110 | 3.7063 | T | upstream | CTNNA1P1 | PPG | - | - | 30 |
| 58 | rs78817974 | 5 | 160881054 | 0.1483 | 4.3502 | T | intron | GABRB2 | PC | - | 5/10 | NA |
| 59 | rs6869521 | 5 | 160889272 | 0.1323 | 3.5378 | C | intron | GABRB2 | PC | - | 4/10 | 7 |
| 60 | rs62381570 | 5 | 160892153 | 0.1774 | 3.8779 | C | intron | GABRB2 | PC | - | 4/10 | 21 |
| 61 | rs62381571 | 5 | 160893196 | -0.3103 | 3.5405 | C | intron | GABRB2 | PC | - | 4/10 | 0 |
| 62 | rs61406751 | 6 | 467581 | 0.1099 | 3.5375 | C | intronNCT | AL512308.1 | lncRNA | - | 1/1 | 25 |
| 63 | rs4713925 | 6 | 11798414 | 0.2192 | 4.6587 | T | intron | ADTRP | PC | - | 2/5 | 24 |
| 64 | rs1330632 | 6 | 57148574 | -0.1539 | 3.3651 | C | intergenic | - | - | - | - | 30 |
| 65 | rs196701 | 6 | 80147187 | -0.1622 | 3.3404 | A | upstream | DBIP1 | PPG | - | - | 25 |
| 66 | rs12524558 | 6 | 105070290 | 0.0884 | 3.4932 | C | intergenic | - | - | - | - | 24 |
| 67 | rs728017 | 6 | 124292594 | 0.1453 | 3.6175 | G | intron | NKAIN2 | PC | - | 1/3 | 28 |

| | | | | | | | | | | | | |
|---|---|---|---|---|---|---|---|---|---|---|---|---|
| 68 | rs1028378 | 6 | 148834018 | 0.1421 | 3.7118 | G | intron | SASH1 | PC | - | 8/19 | 9 |
| 69 | rs1057793 | 6 | 148835416 | 0.0941 | 3.8267 | T | intron | SASH1 | PC | - | 8/19 | 24 |
| 70 | rs12526105 | 6 | 169463997 | 0.1717 | 3.8428 | A | upstream | AL109924.2 | lncRNA | - | - | 24 |
| 71 | rs12672025 | 7 | 7157756 | -0.2302 | 3.4939 | A | regulatory | - | enhancer | - | - | 24 |
| 72 | rs11540586 | 7 | 29962427 | -0.0522 | 3.3685 | C | 3'UTR | SCRN1 | PC | 8/8 | - | 18 |
| 73 | rs17171441 | 7 | 38722499 | -0.0202 | 3.3371 | C | downstream | FAM183B | PC | - | - | 24 |
| 74 | rs334529 | 7 | 47557221 | -0.2477 | 3.4173 | T | intron | TNS3 | PC | - | 2/30 | 21 |
| 75 | rs12705836 | 7 | 78547202 | 0.1631 | 4.0567 | A | intron | MAGI2 | PC | - | 2/21 | 19 |
| 76 | rs9692405 | 7 | 78657099 | 0.1551 | 3.7257 | A | intron | MAGI2 | PC | - | 1/21 | 18 |
| 77 | rs968908 | 7 | 89341720 | 0.1910 | 3.9967 | C | intergenic | - | - | - | - | 27 |
| 78 | rs4730501 | 7 | 111432940 | 0.1801 | 4.0129 | A | upstream | DOCK4 | PC | - | - | 6 |
| 79 | rs74782537 | 7 | 151531336 | -0.1060 | 3.4229 | C | intron | PRKAG2 | PC | - | =L41/15 | 25 |
| 80 | rs4260917 | 8 | 1605294 | 0.0140 | 3.3629 | G | intron | DLGAP2 | PC | - | 8/14 | 18 |
| 81 | rs68112061 | 8 | 2931097 | 0.1412 | 3.5926 | C | intron | CSMD1 | PC | - | 36/55 | 13 |
| 82 | rs4875742 | 8 | 2964206 | 0.1060 | 3.3596 | C | intron | CSMD1 | PC | - | 33/55 | 6 |
| 83 | rs11774005 | 8 | 2966358 | 0.1797 | 3.8609 | A | intron | CSMD1 | PC | - | 31/55 | 27 |
| 84 | rs11786969 | 8 | 2966388 | 0.2596 | 3.6682 | T | intron | CSMD1 | PC | - | 31/55 | 0 |
| 85 | rs67705201 | 8 | 2966633 | -0.1046 | 3.5393 | T | intron | CSMD1 | PC | - | 31/55 | 0 |
| 86 | rs898509 | 8 | 2967576 | 0.1404 | 3.3184 | A | intron | CSMD1 | PC | - | 31/55 | 0 |
| 87 | rs898510 | 8 | 2967609 | 0.2048 | 3.7157 | A | intron | CSMD1 | PC | - | 31/55 | 24 |
| 88 | rs73183533 | 8 | 2967906 | 0.1486 | 3.4300 | A | intron | CSMD1 | PC | - | 30/55 | NA |
| 89 | rs17390567 | 8 | 2968364 | 0.2624 | 3.8858 | C | intron | CSMD1 | PC | - | 30/55 | 4 |
| 90 | rs9650503 | 8 | 3066067 | 0.1588 | 3.5550 | G | intron | CSMD1 | PC | - | 18/55 | 18 |
| 91 | rs7835399 | 8 | 3115052 | 0.1025 | 3.5558 | A | intron | CSMD1 | PC | - | 14/55 | 24 |
| 92 | rs13254027 | 8 | 3125924 | 0.0918 | 3.4061 | A | intron | CSMD1 | PC | - | 14/55 | 0 |
| 93 | rs12114514 | 8 | 17357731 | 0.2544 | 3.7605 | T | intron | SLC7A2 | PC | - | 1/13 | 25 |
| 94 | rs111473411 | 8 | 26590976 | 0.1754 | 3.7784 | T | intergenic | - | - | - | - | 31 |
| 95 | rs2132449 | 8 | 26592353 | 0.1166 | 3.7631 | A | intergenic | - | - | - | - | 0 |
| 96 | rs7821479 | 8 | 26601657 | 0.1283 | 3.8086 | A | downstream | ADRA1A | PC | - | - | 0 |
| 97 | rs2553916 | 8 | 54038872 | -0.0470 | 3.3729 | C | intergenic | - | - | - | - | 1 |
| 98 | rs2717637 | 8 | 54039378 | -0.1441 | 3.5520 | A | intergenic | - | - | - | - | 24 |
| 99 | rs13258651 | 8 | 74317959 | -0.1147 | 3.3973 | A | intergenic | - | - | - | - | 24 |
| 100 | rs34439026 | 8 | 74328861 | -0.0526 | 3.6079 | T | upstream | STAU2-AS1 | lincRNA | - | - | NA |
| 101 | rs34643738 | 8 | 112466724 | 0.1702 | 3.5531 | C | intronNCT | RP11-1101K5.1 | lincRNA | - | 2/4 | 31 |
| 102 | rs2125553 | 8 | 113900367 | 0.1485 | 4.0838 | C | intron | CSMD3 | PC | - | 10/70 | 30 |
| 103 | rs56282194 | 8 | 133574214 | -0.2497 | 3.3955 | A | upstream | HPYR1 | lincRNA | - | - | 24 |
| 104 | rs13439493 | 8 | 135108404 | 0.1289 | 4.3009 | T | intergenic | - | - | - | - | 30 |
| 105 | rs12348020 | 9 | 9069548 | 0.1122 | 3.6063 | T | intron | PTPRD | PC | - | 7/42 | NA |
| 106 | rs4745687 | 9 | 80619811 | 0.0280 | 3.4579 | G | intron | GNAQ | PC | - | 1/6 | 24 |
| 107 | rs10122296 | 9 | 102334338 | 0.1221 | 3.3552 | C | intronNCT | AL359710.1 | lncRNA | - | 2/5 | 30 |
| 108 | rs2771036 | 9 | 108205262 | 0.1851 | 4.0498 | T | downstream | SLC44A1 | PC | - | - | 30 |
| 109 | rs4390017 | 9 | 124633711 | 0.2441 | 4.4056 | T | intron | TTLL11 | PC | - | 6/8 | 30 |
| 110 | rs977754 | 10 | 44817419 | 0.1338 | 4.2885 | T | intron | CXCL12 | PC | - | 3/3 | NA |
| 111 | rs1720367 | 10 | 50908644 | 0.0383 | 3.3385 | A | intron | C10orf53 | PC | - | 2/2 | NA |
| 112 | rs12241006 | 10 | 79030744 | 0.1680 | 4.0976 | A | intron | KCNMA1 | PC | - | 2/26 | 24 |
| 113 | rs7899222 | 10 | 87701735 | 0.1323 | 3.3948 | A | intron | GRID1 | PC | - | 4/15 | 24 |
| 114 | rs58634906 | 10 | 118986090 | 0.1862 | 4.4124 | A | intergenic | - | - | - | - | 7 |
| 115 | rs7393602 | 10 | 118987781 | 0.1926 | 4.0864 | C | intergenic | - | - | - | - | 6 |
| 116 | rs12414919 | 10 | 118991768 | 0.1942 | 4.2218 | A | intergenic | - | - | - | - | 24 |
| 117 | rs2072362 | 10 | 119014023 | 0.1977 | 3.6191 | T | intronNCT | SLC18A2 | RI | - | 5/14 | 6 |
| 118 | rs2072363 | 10 | 119014406 | 0.1050 | 3.6592 | G | intronNCT | SLC18A2 | RI | - | 5/14 | 0 |
| 119 | rs2072364 | 10 | 119014408 | -0.2527 | 3.4115 | C | intronNCT | SLC18A2 | RI | - | 5/14 | 0 |
| 120 | rs363420 | 10 | 119014931 | -0.1674 | 3.5839 | T | intronNCT | SLC18A2 | RI | - | 6/14/ | 0 |
| 121 | rs363245 | 10 | 119016331 | -0.3010 | 4.2371 | G | intronNCT | SLC18A2 | RI | - | 8/14 | NA |
| 122 | rs363220 | 10 | 119016690 | 0.1812 | 4.1104 | T | intronNCT | SLC18A2 | RI | - | 8/14 | 9 |
| 123 | rs2283138 | 10 | 119018679 | 0.0976 | 3.6346 | A | intronNCT | SLC18A2 | RI | - | 9/14 | 20 |
| 124 | rs929493 | 10 | 119019126 | 0.1542 | 3.7680 | T | intronNCT | SLC18A2 | RI | - | 9/14 | 17 |
| 125 | rs1860404 | 10 | 119019177 | 0.0982 | 3.5750 | C | intronNCT | SLC18A2 | RI | - | 9/14 | 6 |
| 126 | rs12575355 | 11 | 2566118 | 0.0538 | 3.4322 | G | intron | KCNQ1 | PC | - | 2/15 | 18 |
| 127 | rs7949361 | 11 | 34238019 | -0.2089 | 3.5399 | T | intron | ABTB2 | PC | - | 1/16 | 20 |
| 128 | rs208679 | 11 | 34454061 | -0.0828 | 3.3996 | G | downstream | CIR1P3 | PPG | - | - | 0 |
| 129 | rs551929 | 11 | 34487192 | -0.1587 | 3.4545 | T | intron | CAT | PC | - | 10/27 | 0 |
| 130 | rs1535720 | 11 | 34500431 | 0.1904 | 4.0198 | C | 3'UTR | ELF5 | PC | 7/7 | - | 0 |
| 131 | rs1535717 | 11 | 34501166 | 0.2274 | 4.1018 | A | 3'UTR | ELF5 | PC | 7/7 | - | 0 |
| 132 | rs12288820 | 11 | 34505926 | 0.0523 | 4.0323 | A | intron | ELF5 | PC | - | 4/6 | 0 |
| 133 | rs10836249 | 11 | 34506338 | 0.1791 | 4.0125 | A | intron | ELF5 | PC | - | 4/6 | 24 |
| 134 | rs263101 | 11 | 35962249 | 0.1586 | 4.4489 | T | upstream | LDLRAD3 | PC | - | - | 30 |
| 135 | rs2510868 | 11 | 58247718 | -0.1352 | 3.5263 | G | intergenic | - | - | - | - | 24 |
| 136 | rs2317091 | 11 | 62960760 | -0.1191 | 3.3447 | A | intron | SLC22A25 | PC | - | 4/8 | 24 |

| | | | | | | | | | | | | |
|---|---|---|---|---|---|---|---|---|---|---|---|---|
| 137 | rs10765595 | 11 | 88263649 | 0.1056 | 3.3656 | G | intron | GRM5 | PC | - | 7/7 | 20 |
| 138 | rs4753765 | 11 | 88593289 | 0.1775 | 3.5589 | T | intron | GRM5 | PC | - | 1/7 | 0 |
| 139 | rs972938 | 11 | 88594590 | 0.1674 | 3.5398 | C | intron | GRM5 | PC | - | 1/7 | 0 |
| 140 | rs982709 | 11 | 88595236 | 0.1178 | 3.7174 | A | intron | GRM5 | PC | - | 1/7 | 0 |
| 141 | rs10831536 | 11 | 88597438 | 0.1539 | 4.3517 | A | intron | GRM5 | PC | - | 1/7 | 6 |
| 142 | rs10831537 | 11 | 88597440 | 0.2411 | 4.2383 | C | intron | GRM5 | PC | - | 1/7 | 13 |
| 143 | rs2221118 | 11 | 88598040 | -0.0233 | 3.4726 | A | intron | GRM5 | PC | - | 1/7 | 0 |
| 144 | rs16919379 | 11 | 93525376 | 0.0883 | 3.5611 | T | intron | MED17 | PC | - | 3/11 | NA |
| 145 | rs73593157 | 11 | 132525442 | -0.3405 | 3.3117 | G | intron | OPCML | PC | - | 2/6 | 24 |
| 146 | rs11180602 | 12 | 76016713 | -0.0140 | 3.3268 | T | intronNCT | AC078923.1 | lncRNA | - | 4/4 | 21 |
| 147 | rs1275582 | 12 | 76255406 | -0.1761 | 3.5606 | T | intronNCT | RP11-114H23.1 | lincRNA | - | 1/4 | 24 |
| 148 | rs4767528 | 12 | 117719054 | 0.1586 | 3.6370 | A | intron | NOS1 | PC | - | 7/28 | 21 |
| 149 | rs9315675 | 13 | 39905865 | -0.1705 | 3.3432 | C | intronNMD | LHFPL6 | NMD | - | 4/13 | 18 |
| 150 | rs9300660 | 13 | 101960273 | -0.1363 | 3.3559 | G | intron | NALCN | PC | - | 7/43 | 18 |
| 151 | rs61035869 | 13 | 101979673 | -0.2466 | 4.5430 | A | intron | NALCN | PC | - | 7/43 | 24 |
| 152 | rs9558839 | 13 | 107391098 | -0.1346 | 3.4232 | G | regulatory | - | enhancer | - | - | 24 |
| 153 | rs1924349 | 13 | 109107293 | 0.1787 | 3.9751 | G | intergenic | - | - | - | - | 30 |
| 154 | rs527264 | 13 | 113736543 | 0.0824 | 3.4030 | T | intron | MCF2L | PC | - | 16/26 | 24 |
| 155 | rs7155706 | 14 | 58447752 | 0.3455 | 4.9518 | A | intronNCT | SLC35F4 | PT | - | 1/1 | 24 |
| 156 | rs937051 | 15 | 29405523 | 0.1847 | 4.0389 | G | intron | APBA2 | PC | - | 11/12 | 24 |
| 157 | rs12591914 | 15 | 61286657 | 0.2100 | 3.3693 | A | intron | RORA | PC | - | 1/10 | 24 |
| 158 | rs58365910 | 15 | 78849034 | 0.1119 | 3.4288 | C | intergenic | - | - | - | - | 0 |
| 159 | rs2036527 | 15 | 78851615 | 0.0866 | 3.4813 | A | intergenic | - | - | - | - | 20 |
| 160 | rs55676755 | 15 | 78898932 | 0.1099 | 3.3178 | G | intron | CHRNA3 | PC | - | 4/5 | 30 |
| 161 | rs10851907 | 15 | 78915864 | -0.0866 | 3.3253 | A | downstream | CHRNB4 | PC | - | - | NA |
| 162 | rs58122900 | 15 | 95246036 | 0.1151 | 4.2837 | A | regulatory | - | enhancer | - | - | NA |
| 163 | rs2531972 | 16 | 4051189 | -0.0033 | 3.3693 | A | intron | ADCY9 | PC | - | 3/10 | 24 |
| 164 | rs9933823 | 16 | 85155843 | -0.0761 | 3.3476 | A | intergenic | - | - | - | - | 24 |
| 165 | rs11641814 | 16 | 85161211 | -0.2128 | 3.5874 | C | intergenic | - | - | - | - | 12 |
| 166 | rs7214224 | 17 | 4320544 | 0.2413 | 3.4149 | G | regulatory | - | PFR | - | - | 24 |
| 167 | rs9889251 | 17 | 37148373 | -0.1186 | 3.3187 | A | downstream | AC006441.2 | PPG | - | - | 24 |
| 168 | rs9898328 | 17 | 40032356 | 0.3593 | 5.5974 | A | intron | ACLY | PC | - | 22/28 | 30 |
| 169 | rs80225105 | 17 | 40032683 | 0.3048 | 3.5012 | A | intron | ACLY | PC | - | 22/28 | 0 |
| 170 | rs1004413 | 18 | 3042297 | 0.1677 | 3.3753 | G | regulatory | - | PFR | - | - | 18 |
| 171 | rs7231904 | 18 | 4596846 | -0.1126 | 3.5383 | A | intergenic | - | - | - | - | 30 |
| 172 | rs8085681 | 18 | 9498749 | 0.1778 | 3.8930 | G | intron | RALBP1 | PC | - | 1/9 | 31 |
| 173 | rs57214198 | 18 | 22082857 | -0.2254 | 3.4948 | T | intronNCT | AC007922.4 | lncRNA | - | 2/2 | 30 |
| 174 | rs749450 | 19 | 4033027 | -0.2404 | 3.4214 | A | intron | PIAS4 | PC | - | 7/10 | NA |
| 175 | rs73029362 | 19 | 34125298 | -0.1702 | 3.3363 | A | intron | CHST8 | PC | - | 1/4 | 30 |
| 176 | rs10409200 | 19 | 39459174 | 0.1667 | 3.5670 | G | intron | FBXO17 | PC | - | 1/5 | 24 |
| 177 | rs11878604 | 19 | 41333284 | 0.2550 | 3.8278 | C | upstream | CYP2F2P | UnitaryPG | - | - | NA |
| 178 | rs12459249 | 19 | 41339896 | 0.1761 | 3.8822 | C | intronNCT | - | lincRNA | - | 1/1 | 6 |
| 179 | rs10853742 | 19 | 41340573 | 0.1641 | 3.6628 | A | intronNCT | - | lincRNA | - | 1/1 | 0 |
| 180 | rs11667314 | 19 | 41340983 | 0.1761 | 3.6877 | C | intronNCT | - | lincRNA | - | 1/1 | 0 |
| 181 | rs12461964 | 19 | 41341229 | 0.0630 | 3.5555 | G | intronNCT | - | lincRNA | - | 1/1 | 0 |
| 182 | rs60446182 | 19 | 41347998 | 0.1049 | 3.6879 | G | downstream | CYP2A6 | PC | - | - | 5 |
| 183 | rs56113850 | 19 | 41353107 | 0.3276 | 5.4667 | C | intron | CYP2A6 | PC | - | 4/8 | 30 |
| 184 | rs57837628 | 19 | 41357910 | 0.2971 | 4.2531 | G | upstream | CYP2A6 | PC | - | - | 0 |
| 185 | rs113029345 | 19 | 41370176 | 0.2254 | 4.0284 | C | intronNMD | - | NMD | - | 1/3 | 5 |
| 186 | rs12461383 | 19 | 41370338 | -0.2301 | 4.5836 | G | intronNMD | - | NMD | - | 1/3 | 0 |
| 187 | rs10410975 | 19 | 41473425 | -0.0538 | 3.4103 | A | intergenic | - | - | - | - | NA |
| 188 | rs16986309 | 19 | 55710074 | -0.2234 | 3.4012 | A | missense | PTPRH | PC | 6/18 | - | 30 |
| 189 | rs2422843 | 20 | 3081680 | -0.0877 | 3.4452 | G | regulatory | - | PFR | - | - | 21 |
| 190 | rs1002929 | 20 | 5209217 | 0.1348 | 3.6180 | A | intergenic | - | - | - | - | 25 |
| 191 | rs805726 | 20 | 5695174 | 0.0299 | 3.4958 | G | intergenic | - | - | - | - | 21 |
| 192 | rs13433103 | 20 | 11251165 | 0.1568 | 4.1499 | G | upstream | RP4-734C18.1 | lincRNA | - | - | 26 |
| 193 | rs4239715 | 20 | 16138249 | -0.1356 | 3.3378 | G | intergenic | - | - | - | - | 24 |
| 194 | rs17801258 | 20 | 38454576 | -0.0631 | 3.4079 | C | regulatory | - | enhancer | - | - | NA |
| 195 | rs12481177 | 20 | 57835389 | -0.2391 | 3.4544 | A | downstream | ZNF831 | PC | - | - | 30 |
| 196 | rs736898 | 22 | 22711786 | 0.1220 | 3.3520 | T | downstream | IGLV5-48 | IGVG | - | - | 24 |
| 197 | rs7285001 | 22 | 27896321 | 0.1570 | 3.6164 | A | intergenic | - | - | - | - | 24 |
| 198 | rs132950 | 22 | 38548972 | 0.1004 | 3.4789 | A | intron | PLA2G6 | PC | - | 2/16 | 24 |
| 199 | rs132952 | 22 | 38549373 | 0.1410 | 3.6877 | C | intron | PLA2G6 | PC | - | 2/16 | 6 |
| 200 | rs79334853 | 22 | 46714748 | -0.0659 | 3.4641 | T | intron | GTSE1 | PC | - | 7/11 | 19 |

**Supplementary Tables 2 and 3 ReadMe**

| **Label** | **Definition** |
|---|---|
| Index | Index by chromosome coordinate |
| SNP | Variant ID |
| CHR | Chromosome number |
| POS | Base pair (hg19) |
| beta | Coefficient, marginal analysis |
| -log10p | Significance, marginal analysis |
| Allele | Risk Allele |
| Function | Sequence function |
| Symbol | Gene Acronym |
| Biotype | Gene class, if any |
| Exon | Exonic location, if any |
| Intron | Intronic location, if any |
| Count | Number of penalized regression models variant trained in with range 0-36.<br>NA indicates the variant was not available in the UW-TTURC dataset,<br>and excluded from model training in the MEC as not available in UW-TTURC dataset. |
| 3'UTR | 3_prime_UTR_variant |
| 5'UTR | 5_prime_UTR_variant |
| antisense | antisense |
| CTCF | CTCF_binding_site |
| downstream | downstream_gene_variant |
| enhancer | enhancer |
| IGVG | IG_V_gene |
| intergenic | intergenic_variant |
| intron | intron_variant |
| intronNCT | intron,non_coding_transcript_variant |
| intronNMD | intron,NMD_transcript_variant |
| lincRNA | lincRNA |
| miscRNA | misc_RNA |
| missense | missense_variant |
| NCTexon | non_coding_transcript_exon_variant |
| NMD | nonsense_mediated_decay |
| not defined | - |
| PC | protein_coding |
| PFR | promoter_flanking_region |
| PPG | processed_pseudogene |
| PT | processed_transcript |
| regulatory | regulatory_region_variant |
| RI | retained_intron |
| synonymous | synonymous_variant |
| TUPG | transcribed_unprocessed_pseudogene |
| UnitaryPG | unitary_pseudogene |
| UPPG | unprocessed_pseudogene |
| upstream | upstream_gene_variant |

**Supplementary Table 4:** Demographics of UW-TTURC Sample, by RCT

| RCT | ED SR | Depend | TTURC2 | *P* |
|---|---|---|---|---|
| NCT ID | 01621009 | 01621022 | 00332644 | *no test* |
| Citation | (McCarthy et al. 2008) | (Piper et al. 2007) | (Piper et al. 2009) | *no test* |
| Site | Madison | Milwaukee | Both cities | *no test* |
| Years of Interview | 2001-2002 | 2001-2002 | 2005-2007 | *no test* |
| N (%) GSS | 186 (9.98%) | 376 (20.17%) | 1302 (69.85%) | *no test* |
| % of RCT | 40.2% of 463 | 61.8% of 608 | 86.6% of 1504 | *P* < .001** |
| Age | | | | |
| GSS Age | 37.77 (11.19) | 41.54 (10.65) | 44.75 (11.23) | $P < .001^{\alpha}$ |
| RCT Age* | 39.4 (11.3) | 41.8 (11.3) | 44.7 (11.1) | $P < .001^{\alpha}$ |
| | | | | *n.s.* GSSvRCT |
| Sex | | | | |
| GSS Sex | 97 (52.2%) | 218 (58.0%) | 775 (59.5%) | *n.s.* in GSS |
| RCT Sex* | 233 (50.3%) | 352 (57.9%) | 876 (58.2%) | *mixed* *** |
| | | | | *n.s.* GSSvRCT |
| Ethnicity | | | | |
| GSS Black | 10 (5.38%) | 88 (23.4%) | 162 (12.4%) | $P < .005^{\beta}$ |
| GSS White | 176 (94.6%) | 288 (76.6%) | 1140 (87.6%) | |
| RCT Black* | 26 (5.94%) | 130 (22.5%) | 204 (14.0%) | $P < .001^{\beta}$ |
| RCT White* | 412 (94.1%) | 449 (77.6%) | 1258 (86.0%) | |
| | | | | *n.s.* GSSvRCT |

*From original publications. **N participating vs not participating vs RCT $\chi^2$ test. ***ED SR *vs* Depend and ED SR *vs* TTURC2, *P* < .05; Depend *vs* TTURC2, *n.s.*. $^{\alpha}$Age comparisons by *t* test. $^{\beta}$Ethnicity comparisons by $\chi^2$ test.

**Supplementary Table 5:** Demographics of the GSS, by Sex and by Ethnicity

| | Female | Male | Black | White |
|---|---|---|---|---|
| N | 1,090 | 774 | 260 | 1,604 |
| Age | 42.94 (11.19) | 44.07 (11.50)* | 45.22 (9.30) | 43.11 (11.60)** |
| Sex (F) | 1090 (58.48%) | 774 (41.52%)** | 174 (66.92%) | 916 (57.10%)** |
| Ethnicity | | | 260 (13.95%) | 1604 (86.05%) |
| Hispanic | 12 (1.10%) | 12 (1.55%) | 4 (1.53%) | 20 (1.25%) |
| Age of Initiation | 14.25 (3.71) | 14.16 (3.74) | 14.89 (4.27) | 14.10 (3.62) |
| Years Smoked | 28.68 (11.16) | 29.93 (11.77)* | 30.32 (10.39) | 29.02 (11.58) |

*$P < .05$, **$P < .005$. Female:Male and Black:White *P* indicator superscripts in Male and in White columns, respectively.

**Supplementary Table 6:** FTND, CPD, and TTFC, by Sex and by Ethnicity

| | All | Female | Male | Black | White |
|---|---|---|---|---|---|
| FTND N | 1843 | 1077 | 766 | 254 | 1589 |
| M (SD) | 5.41 (2.16) | 5.25 (2.11) | 5.65 (2.21)$^{\gamma}$ | 5.54 (2.04) | 5.39 (2.18) |
| CPD N | 1862 | 1090 | 772 | 260 | 1602 |
| 1-10 N (%) | 99 (5.3) | 75 (6.9) | 24 (3.1)$^{\gamma}$ | 29 (11.2) | 70 (4.4)$^{\gamma}$ |
| 11-20 N (%) | 988 (53.1) | 650 (59.6) | 338 (43.8) | 164 (63.1) | 824 (51.4) |
| 21-30 N (%) | 533 (28.6) | 272 (25.0) | 261 (33.8) | 49 (18.9) | 484 (30.2) |
| ≥ 31 N (%) | 242 (13.0) | 93 (8.5) | 149 (19.3) | 18 (6.9) | 224 (14.0) |
| TTFC N | 1861 | 1089 | 772 | 260 | 1601 |
| > 60 N (%) | 137 (7.4) | 84 (7.7) | 53 (6.9) | 10 (3.9) | 127 (7.9)$^{\gamma}$ |
| 31-60 N (%) | 285 (15.3) | 167 (15.3) | 118 (15.3) | 36 (13.9) | 249 (15.6) |
| 6-30 N (%) | 875 (47.0) | 500 (45.9) | 375 (48.6) | 108 (41.5) | 767 (47.9) |
| 0-5 N (%) | 564 (30.3) | 338 (31.0) | 226 (29.3) | 106 (40.8) | 458 (28.6) |

$^{\alpha}P < .05$, $^{\beta}P < .005$, $^{\gamma}P < .001$. Female:Male and Black:White *P* indicator superscripts in Male and in White columns, respectively.

**Supplementary Table 7:** TDS Dependence, by Sex and by Ethnicity

| | All* | Female | Male | Black | White |
|---|---|---|---|---|---|
| N | 1856 | 1086 | 770 | 260 | 1596 |
| TDS | 6.81 (1.91) | 6.95 (1.85) | 6.62 (1.98)$^{\gamma}$ | 6.63 (2.25) | 6.84 (1.85)$^{\alpha}$ |

*M (SD). $^{\alpha}P < .05$, $^{\beta}P < .005$, $^{\gamma}P < .001$. Female:Male and Black:White *P* indicator superscripts in Male and in White columns, respectively.

**Supplementary Table 8:** WISDM Primary Dependence Motives, by Sex and by Ethnicity

| | All* | Female | Male | Black | White |
|---|---|---|---|---|---|
| N | 1860 | 1089 | 771 | 260 | 1600 |
| Automaticity | 4.60 (1.68) | 4.67 (1.68) | 4.50 (1.68)$^{\alpha}$ | 4.66 (1.77) | 4.59 (1.67) |
| Loss Of Control | 5.15 (1.43) | 5.28 (1.42) | 4.98 (1.44)$^{\gamma}$ | 4.93 (1.63) | 5.19 (1.40)$^{\gamma}$ |
| Craving | 4.94 (1.31) | 4.96 (1.32) | 4.90 (1.30) | 4.96 (1.53) | 4.93 (1.27) |
| Tolerance | 4.94 (1.41) | 4.95 (1.44) | 4.92 (1.37) | 5.19 (1.43) | 4.89 (1.40)$^{\alpha}$ |
| PDM Total | 19.62 (4.75) | 19.86 (4.70) | 19.29 (4.80)$^{\gamma}$ | 19.74 (5.37) | 19.61 (4.64) |

*M (SD). $^{\alpha}P < .05$, $^{\beta}P < .005$, $^{\gamma}P < .001$. Female:Male and Black:White *P* indicator superscripts in Male and in White columns, respectively.

**Supplementary Table 9:** NDSS Scales in the GSS Participants, by Sex and by Ethnicity

| | All* | Female | Male | Black | White |
|---|---|---|---|---|---|
| N | 1809 | 1052 | 757 | 244 | 1565 |
| Drive | 0.04 (1.04) | 0.11 (1.05) | -0.05 (1.01)$^{\gamma}$ | -0.07 (1.15) | 0.06(1.02) |
| N | 1820 | 1062 | 758 | 251 | 1569 |
| Priority | 0.02 (1.06) | 0.06 (1.09) | -0.04 (1.02) | 0.29 (1.16) | -0.03 (1.04)$^{\gamma}$ |
| N | 1814 | 1055 | 759 | 244 | ???? |
| Tolerance | -0.36 (1.13) | -0.40 (1.14) | -0.31 (1.12) | -0.28 (1.05) | -0.37 (1.14) |
| N | 1815 | 1061 | 754 | 247 | 1568 |
| Continuity | -0.08 (1.02) | -0.13 (1.04) | 0.00 (1.00)$^{\alpha}$ | -0.31 (1.07) | -0.04 (1.01)$^{\gamma}$ |
| N | 1813 | 1060 | 753 | 249 | 1564 |
| Stereotypy | -0.09 (1.05) | -0.19 (1.06) | 0.04 (1.02)$^{\gamma}$ | 0.13 (1.12) | -0.13 (1.03)$^{\gamma}$ |
| N | 1800 | 1050 | 750 | 243 | 1557 |
| NDSS Total | -0.17 (0.92) | -0.18 (0.92) | -0.17 (0.92) | -0.07 (1.07) | -0.19 (0.89) |

*M (SD). $^{\alpha}P < .05$, $^{\beta}P < .005$, $^{\gamma}P < .001$. Female:Male and Black:White *P* indicator superscripts in Male and in White columns, respectively.

**Supplementary Table 10. Predicted Biomarker Interactions in Association with Nicotine Dependence**

| Measure | biomarker | interaction | N | coef | se | p | p<.05 |
|---|---|---|---|---|---|---|---|
| ftnd_1 | NMR_pred | ethnicity | 1861 | 0.1120 | 0.2506 | 0.6549 | |
| ftnd_1 | NMR_pred | sex | 1861 | -0.0342 | 0.1553 | 0.8259 | |
| ftnd_4 | NMR_pred | ethnicity | 1862 | 0.0830 | 0.2202 | 0.7064 | |
| ftnd_4 | NMR_pred | sex | 1862 | 0.0684 | 0.1364 | 0.6165 | |
| ftnd_total | NMR_pred | ethnicity | 1843 | 0.2192 | 0.6346 | 0.7298 | |
| ftnd_total | NMR_pred | sex | 1843 | 0.3025 | 0.3888 | 0.4366 | |
| tds_score | NMR_pred | ethnicity | 1856 | -0.0523 | 0.5605 | 0.9256 | |
| tds_score | NMR_pred | sex | 1856 | -0.0745 | 0.3477 | 0.8305 | |
| wisdm_auto | NMR_pred | ethnicity | 1860 | -0.0562 | 0.4861 | 0.9079 | |
| wisdm_auto | NMR_pred | sex | 1860 | -0.1746 | 0.3012 | 0.5621 | |
| wisdm_control | NMR_pred | ethnicity | 1860 | -0.1805 | 0.4102 | 0.6600 | |
| wisdm_control | NMR_pred | sex | 1860 | 0.2146 | 0.2541 | 0.3985 | |
| wisdm_craving | NMR_pred | ethnicity | 1860 | -0.3107 | 0.3864 | 0.4215 | |
| wisdm_craving | NMR_pred | sex | 1860 | -0.1586 | 0.2395 | 0.5080 | |
| wisdm_tolerance | NMR_pred | ethnicity | 1860 | 0.0636 | 0.4104 | 0.8769 | |
| wisdm_tolerance | NMR_pred | sex | 1860 | 0.2283 | 0.2543 | 0.3694 | |
| wisdm_score | NMR_pred | ethnicity | 1860 | -4.2928 | 3.8115 | 0.2602 | |
| wisdm_score | NMR_pred | sex | 1860 | -2.2567 | 2.3623 | 0.3396 | |
| ndss_continuity | NMR_pred | ethnicity | 1815 | 0.6245 | 0.3054 | 0.0410 | * |
| ndss_continuity | NMR_pred | sex | 1815 | 0.4236 | 0.1873 | 0.0239 | * |
| ndss_drive | NMR_pred | ethnicity | 1809 | 0.1634 | 0.3149 | 0.6038 | |
| ndss_drive | NMR_pred | sex | 1809 | 0.0128 | 0.1915 | 0.9467 | |
| ndss_priority | NMR_pred | ethnicity | 1820 | 0.3124 | 0.3179 | 0.3259 | |
| ndss_priority | NMR_pred | sex | 1820 | -0.0931 | 0.1934 | 0.6303 | |
| ndss_stereotypy | NMR_pred | ethnicity | 1813 | -0.6228 | 0.3100 | 0.0446 | * |
| ndss_stereotypy | NMR_pred | sex | 1813 | 0.0754 | 0.1911 | 0.6932 | |
| ndss_tolerance | NMR_pred | ethnicity | 1814 | 0.0083 | 0.3449 | 0.9809 | |
| ndss_tolerance | NMR_pred | sex | 1814 | -0.1221 | 0.2078 | 0.5569 | |
| ndss_score | NMR_pred | ethnicity | 1800 | -0.2046 | 0.2814 | 0.4672 | |
| ndss_score | NMR_pred | sex | 1800 | -0.0470 | 0.1705 | 0.7826 | |
| ftnd_1 | TNE_pred | ethnicity | 1861 | 0.0268 | 0.0549 | 0.6253 | |
| ftnd_1 | TNE_pred | sex | 1861 | -0.0214 | 0.0351 | 0.5432 | |
| ftnd_4 | TNE_pred | ethnicity | 1862 | 0.0015 | 0.0484 | 0.9755 | |
| ftnd_4 | TNE_pred | sex | 1862 | -0.0440 | 0.0309 | 0.1551 | |
| ftnd_total | TNE_pred | ethnicity | 1843 | -0.0489 | 0.1397 | 0.7264 | |
| ftnd_total | TNE_pred | sex | 1843 | -0.1018 | 0.0879 | 0.2474 | |
| tds_score | TNE_pred | ethnicity | 1856 | -0.1973 | 0.1229 | 0.1084 | |
| tds_score | TNE_pred | sex | 1856 | -0.1374 | 0.0786 | 0.0805 | |
| wisdm_auto | TNE_pred | ethnicity | 1860 | 0.0589 | 0.1068 | 0.5809 | |
| wisdm_auto | TNE_pred | sex | 1860 | 0.0879 | 0.0682 | 0.1977 | |
| wisdm_control | TNE_pred | ethnicity | 1860 | -0.0291 | 0.0900 | 0.7466 | |
| wisdm_control | TNE_pred | sex | 1860 | 0.0414 | 0.0576 | 0.4720 | |
| wisdm_craving | TNE_pred | ethnicity | 1860 | -0.0400 | 0.0848 | 0.6371 | |
| wisdm_craving | TNE_pred | sex | 1860 | -0.0033 | 0.0542 | 0.9510 | |
| wisdm_tolerance | TNE_pred | ethnicity | 1860 | 0.0177 | 0.0900 | 0.8441 | |
| wisdm_tolerance | TNE_pred | sex | 1860 | -0.0253 | 0.0575 | 0.6600 | |
| wisdm_score | TNE_pred | ethnicity | 1860 | -0.7279 | 0.8364 | 0.3842 | |
| wisdm_score | TNE_pred | sex | 1860 | 0.6810 | 0.5348 | 0.2030 | |
| ndss_continuity | TNE_pred | ethnicity | 1815 | 0.1002 | 0.0682 | 0.1418 | |
| ndss_continuity | TNE_pred | sex | 1815 | 0.0663 | 0.0427 | 0.1206 | |
| ndss_drive | TNE_pred | ethnicity | 1809 | -0.0799 | 0.0695 | 0.2504 | |
| ndss_drive | TNE_pred | sex | 1809 | -0.0177 | 0.0437 | 0.6859 | |
| ndss_priority | TNE_pred | ethnicity | 1820 | 0.0137 | 0.0690 | 0.8429 | |
| ndss_priority | TNE_pred | sex | 1820 | -0.0589 | 0.0438 | 0.1784 | |
| ndss_stereotypy | TNE_pred | ethnicity | 1813 | -0.1994 | 0.0685 | 0.0036 | * |
| ndss_stereotypy | TNE_pred | sex | 1813 | 0.0533 | 0.0435 | 0.2207 | |
| ndss_tolerance | TNE_pred | ethnicity | 1814 | 0.0395 | 0.0765 | 0.6062 | |
| ndss_tolerance | TNE_pred | sex | 1814 | -0.0408 | 0.0474 | 0.3895 | |
| ndss_score | TNE_pred | ethnicity | 1800 | -0.1194 | 0.0621 | 0.0547 | |
| ndss_score | TNE_pred | sex | 1800 | -0.0172 | 0.0388 | 0.6565 | |

**Supplementary Table 10.Predicted Biomarker Interactions in Association with Nicotine Dependence**

| **Label** | **Definition** |
|---|---|
| Measure | Nicotine Dependence item, subscale or total scale |
| Biomarker | Predicted nicotine biomarker |
| Interaction | Demographic variable tested for Interaction |
| N | N Participants with Nicotine Dependence |
| coef | Coefficient of interaction variable |
| se | Standard Error of Interaction variable |
| p | P value of interaction variable |
| p<.05 | Identifies nominally significant interaction variable |
| ftnd_1 | Time To First Cigarette item in the Fagerström Test for Nicotine Dependence (FTND) |
| ftnd_4 | Cigarettes Per Day item in the FTND |
| ftnd_total | FTND total score |
| tds_score | Tobacco Dependence Scale (TDS) |
| wisdm_auto | Automaticity subscale of the Wisconsin Inventory of Smoking Dependence Motives (WISDM) |
| wisdm_control | Loss of Control subscale, WISDM |
| wisdm_craving | Cravings subscale, WISDM |
| wisdm_tolerance | Tolerance subscale, WISDM |
| wisdm_score | Total score of the four Primary Dependence Motives of the WISDM |
| ndss_continuity | Continuity subscale of the Nicotine Dependence Syndrome Scale (NDSS) |
| ndss_drive | Drive subscale, NDSS |
| ndss_priority | Priority subscale, NDSS |
| ndss_stereotypy | Stereotypy subscale, NDSS |
| ndss_tolerance | Tolerance subscale, NDSS |
| ndss_score | Total score of the NDSS |
| NMR_pred | Predicted uNMR |
| TNE_pred | Predicted TNE |

### FIGURE LEGENDS

**Supplementary Figure 1.** Regression Tree Model, Urinary Nicotine Metabolite Ratio, Without CPD

**Supplementary Figure 2.** Regression Tree Model, Total Nicotine Equivalents, Without CPD

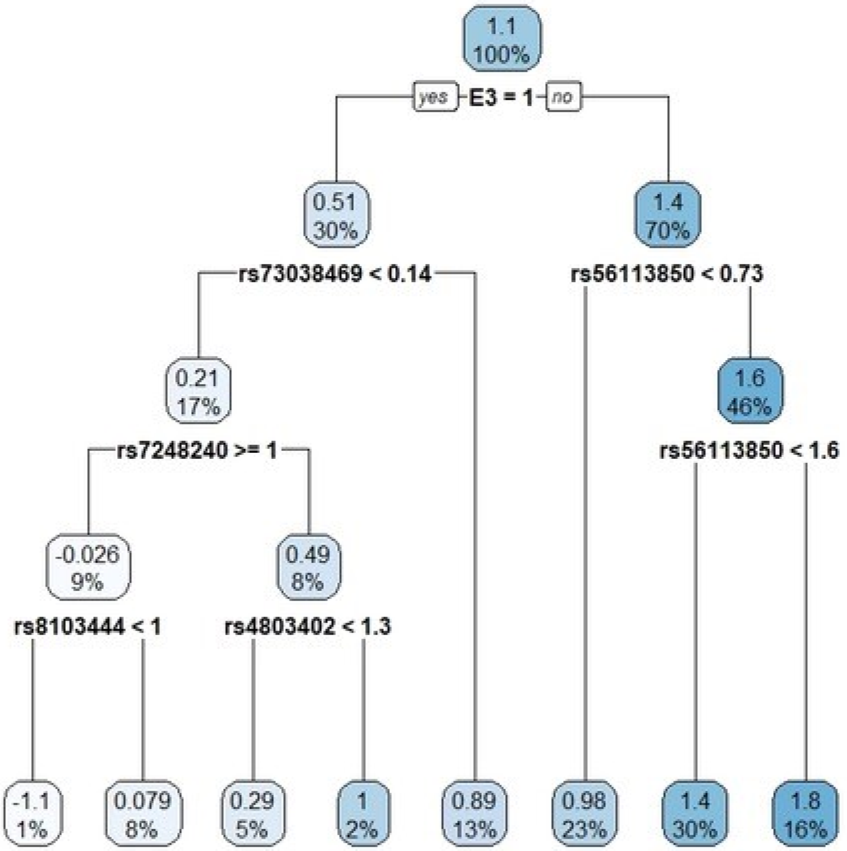
1.1
100%
yes
E3 = 1
no
0.51
30%
1.4
70%
rs73038469 < 0.14
rs56113850 < 0.73
0.21
17%
1.6
46%
rs7248240 >= 1
rs56113850 < 1.6
-0.026
9%
0.49
8%
rs8103444 < 1
rs4803402 < 1.3
-1.1
1%
0.079
8%
0.29
5%
1
2%
0.89
13%
0.98
23%
1.4
30%
1.8
16%

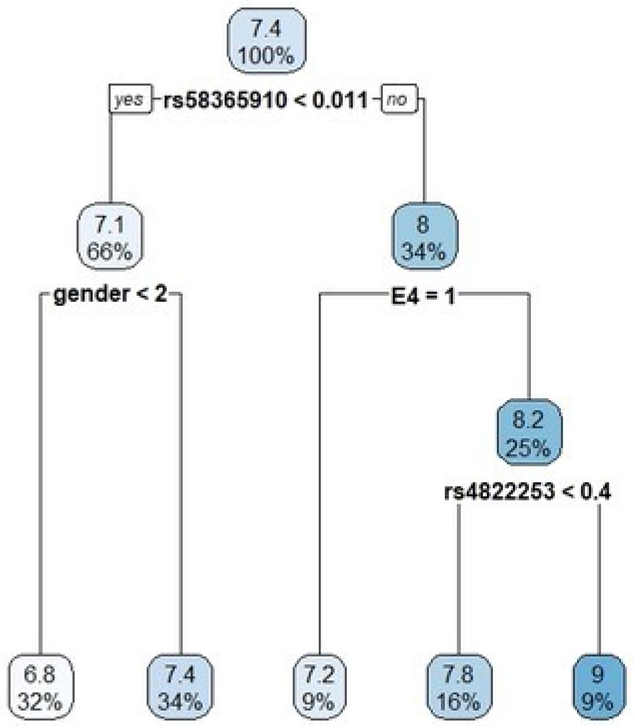
7.4
100%
yes
rs58365910 < 0.011
no
7.1
66%
8
34%
gender < 2
E4 = 1
8.2
25%
rs4822253 < 0.4
6.8
32%
7.4
34%
7.2
9%
7.8
16%
9
9%
